# Large-Scale Meta-Analysis of Potential Biomarkers for Treatment Response to Anti-PD-1/PD-L1 Immune Checkpoint Inhibitors

**DOI:** 10.1101/2020.11.25.20238865

**Authors:** Arshiya Mariam, Suneel Kamath, Kimberly Schveder, Howard L. McLeod, Daniel M. Rotroff

## Abstract

Immune checkpoint inhibitors (ICIs) blocking programmed death receptor 1 or its ligand (anti-PD-1/PD-L1) are a burgeoning class of promising cancer treatments. However, not all patients respond to these treatments. Although studies have suggested potential biomarkers to predict patients likely to respond to treatment, no consensus biomarker has been identified. Here, 95 peer-reviewed studies representing 18,978 subjects across 15 cancer types were meta-analyzed to determine biomarkers that best predicted ICI response within and across cancers. Performance was assessed using the sensitivity, specificity, area under the receiver operating characteristic curve, and others. Across all cancers, multimodal biomarkers and tumor mutational burden discriminated ICI response better than PD-L1 immunohistochemistry assays (IHC) (*P*=.04) with sensitivities of 0.57 and 0.70 and specificities of 0.76 and 0.53, respectively. Marginal improvements were also observed for nascent microbiome biomarkers compared to IHC (*P*=.06). Current ICI biomarker performances indicate that additional research is needed to develop highly accurate and precise biomarkers for widespread clinical adoption.

## Introduction

Immune checkpoint inhibitors (ICIs) have become a cornerstone of cancer therapy across multiple histologies.^1,2^ ICIs blocking programmed death protein receptor 1(PD-1) or its ligand (PD-L1) are at the forefront of clinical implementation. These therapies re-activate the immune response to tumor cells by inhibiting the interaction of PD-L1 and PD-1, and multiple studies have demonstrated their clinical benefit over standard treatments.^3–7^ Although ICIs show evidence of durable clinical benefit for individuals that respond, a systematic meta-analysis estimates that the overall response rate to anti-PD-1/anti-PD-L1 therapies is 24% (95% CI= 21%-28%).^2^ In addition, approximately 16% (95% CI: 12%-21%) of patients experience significant toxicity, including colitis, pneumonitis and endocrine organ dysfunction.^2^ For these reasons, it is critical that robust predictive biomarkers are identified that can guide clinical decision making.

Many studies have explored whether PD-L1 or PD-1 protein expression^8–11^, tumor mutational burden^12–16^, specific somatic mutations (e.g., KRAS, BRAF)^2,17–22^ and more recently, immune related adverse events^23^ can discriminate between responders and non-responders to anti-PD-1/anti-PD-L1 immunotherapies. However, results from these studies are often inconsistent or inconclusive. For example, Bellmunt et al. reported a PD-L1 expression threshold above 10% discriminated patients with urothelial bladder cancer^24^; whereas Massard et al. reported a threshold of 25% for the same cancer type.^25^ Differences in patient populations, samples collection, sample processing, technological platform, biomarker thresholds and the ICI used may all impact variability across studies. In addition to methodological differences, many ICI studies have limited sample sizes that may impact statistical power for discovering biomarkers. Although most reviews aim to qualitatively condense information across studies, the predictive accuracies of biomarkers are not always summarized in a quantitative manner ^26,27^. Relevant statistical summaries for these biomarkers can be ascertained from the literature using meta-analysis, and this approach has been gaining traction as an approach to develop consensus around important clinical questions^2,28^. An additional benefit of meta-analyses is that biomarkers can be concurrently evaluated across different treatments, threshold values and cancer types. Here we conducted the largest meta-analysis of predictive biomarkers for ICI therapy to date, including 95 peer-reviewed studies representing data from 18,978 patients. We also investigated whether the microbiome or immune mediated adverse events show evidence of biomarker potential. Furthermore, whereas most studies qualitatively assess area under the receiver operating characteristic curve (AUCs) performance, we evaluated whether the AUCs for biomarker performance were statistically different from one another. The objective of this study was to provide a comprehensive evaluation of the current state of predictive accuracies of the most commonly considered biomarkers for ICI treatment response.

## Results

After performing quality control, a total of 95 studies met the inclusion criteria for analysis, and all studies were conducted between the years 2010-2020. Objective response rate was reported in 85.27% of studies, and clinical benefit (CB) and six-months progression free survival (PFS) were reported in 8.42% and 6.30% of studies, respectively. The counts for reported studies are provided in Supplementary Figure 1. The most frequent cancer types in the dataset were non-small -cell lung cancer (27.40%) and melanoma (23.20%), and the most frequently explored biomarker for all cancer types was PD-L1 expression (Supplementary Figure 1). Below we present the meta-analysis results by cancer-type (Fig. 1 and Table 1) followed by an overall characterization across cancer types for each biomarker (Fig. 2 and Table 2). The meta-analysis results across all cancer types and other analyses are shown in Supplemental Table 5-8. All of the included studies are listed in Supplementary Table 2 and Supplementary Table 9.

**Table 1.** Meta-analyzed estimates of each biomarker for most commonly reported cancer categories

| Cancer | Biomarker | Studies (N) | Individuals (N) | gAUC<sup>1</sup> (95% CI) | Sensitivity (95% CI) | Specificity (95% CI) | PPV<sup>2</sup> (95% CI) | NPV<sup>3</sup> (95% CI) |
| --- | --- | --- | --- | --- | --- | --- | --- | --- |
| Melanoma | MB | 2 | 128 | 0.83 (0.33-0.97) | 0.58 (0.21-0.88) | 0.90 (0.50-0.99) | 0.87 (0.48-0.98) | 0.65 (0.2-0.93) |
|  | mIHC/IF | 2 | 157 | 0.82 (0.53-0.93) | 0.84 (0.62-0.95) | 0.67 (0.44-0.84) | 0.77 (0.57-0.90) | 0.78 (0.49-0.93) |
|  | Multimodal | 2 | 231 | 0.65 (0.56-0.73) | 0.64 (0.51-0.75) | 0.62 (0.54-0.70) | 0.61 (0.52-0.69) | 0.65 (0.53-0.76) |
|  | PD-L1 IHC | 14 | 2313 | 0.6 (0.53-0.66) | 0.65 (0.52-0.76) | 0.51 (0.39-0.63) | 0.50 (0.45-0.56) | 0.66 (0.56-0.74) |
|  | TMB | 4 | 227 | 0.37 (0.33-0.83) | 0.86 (0.72-0.94) | 0.36 (0.29-0.44) | 0.57 (0.50-0.63) | 0.72 (0.52-0.86) |
| Multiple cancers | Others<sup>4</sup> | 2 | 107 | 0.66 (0.56-0.72) | 0.61 (0.40-0.78) | 0.64 (0.50-0.76) | 0.40 (0.32-0.49) | 0.78 (0.72-0.84) |
|  | PD-L1 IHC | 6 | 693 | 0.64 (0.55-0.72) | 0.68 (0.54-0.79) | 0.55 (0.46-0.64) | 0.33 (0.24-0.43) | 0.85 (0.74-0.91) |
|  | TMB | 3 | 231 | 0.75 (0.58-0.87) | 0.84 (0.65-0.94) | 0.59 (0.44-0.73) | 0.63 (0.33-0.86) | 0.83 (0.47-0.96) |
| Non-small-cell lung cancer | Multimodal | 3 | 304 | 0.71 (0.43-0.9) | 0.43 (0.23-0.65) | 0.88 (0.69-0.96) | 0.61 (0.48-0.73) | 0.76 (0.70-0.81) |
|  | PD-L1 IHC | 22 | 5623 | 0.61 (0.56-0.66) | 0.67 (0.62-0.72) | 0.46 (0.39-0.54) | 0.45 (0.38-0.51) | 0.69 (0.60-0.77) |
|  | TMB | 9 | 1285 | 0.7 (0.6-0.74) | 0.58 (0.46-0.69) | 0.69 (0.63-0.75) | 0.58 (0.50-0.66) | 0.69 (0.60-0.77) |

<sup>1</sup>global area under the receiver operating characteristic curve; <sup>2</sup>positive predictive value; <sup>3</sup>negative predictive value; <sup>4</sup>listed in Supplemental Table 3.

**Table 2.** Meta-analyzed estimates of biomarkers across all studies and cancer types

| Biomarker | Studies (N) | Individuals (N) | gAUC<sup>1</sup> (95% CI) | Sensitivity (95% CI) | Specificity (95% CI) | PPV<sup>2</sup> (95% CI) | NPV<sup>3</sup> (95% CI) |
| --- | --- | --- | --- | --- | --- | --- | --- |
| AEs | 1 | 1747 |  | 0.36 | 0.66 | 0.21 | 0.81 |
| GEP | 5 | 783 | 0.56 (0.49-0.65) | 0.71 (0.55-0.84) | 0.48 (0.41-0.56) | 0.45 (0.30-0.61) | 0.73 (0.54-0.86) |
| IMDC | 2 | 547 | 0.57 (0.32-0.83) | 0.72 (0.17-0.97) | 0.43 (0.11-0.83) | 0.59 (0.51-0.66) | 0.58 (0.34-0.79) |
| MB | 3 | 228 | 0.81 (0.38-0.95) | 0.48 (0.19-0.78) | 0.90 (0.67-0.98) | 0.79 (0.53-0.92) | 0.62 (0.30-0.86) |
| mIHC/IF | 3 | 177 | 0.81 (0.58-0.91) | 0.85 (0.68-0.94) | 0.66 (0.49-0.80) | 0.78 (0.63-0.89) | 0.76 (0.56-0.89) |
| Multimodal | 5 | 559 | 0.7 (0.61-0.76) | 0.57 (0.47-0.67) | 0.76 (0.66-0.83) | 0.50 (0.41-0.59) | 0.81 (0.71-0.87) |
| Others<sup>4</sup> | 10 | 1001 | 0.67 (0.58-0.71) | 0.60 (0.50-0.69) | 0.69 (0.56-0.79) | 0.46 (0.37-0.55) | 0.81 (0.72-0.87) |
| PDL1-IHC | 76 | 13909 | 0.58 (0.56-0.6) | 0.64 (0.60-0.69) | 0.49 (0.44-0.53) | 0.40 (0.36-0.45) | 0.72 (0.67-0.76) |
| TMB | 15 | 1814 | 0.65 (0.59-0.68) | 0.70 (0.63-0.77) | 0.53 (0.46-0.60) | 0.54 (0.47-0.60) | 0.70 (0.63-0.76) |

<sup>1</sup>global area under the receiver operating characteristic curve; <sup>2</sup>positive predictive value; <sup>3</sup>negative predictive value; <sup>4</sup>listed in Supplemental Table 3.

**Fig 1.**
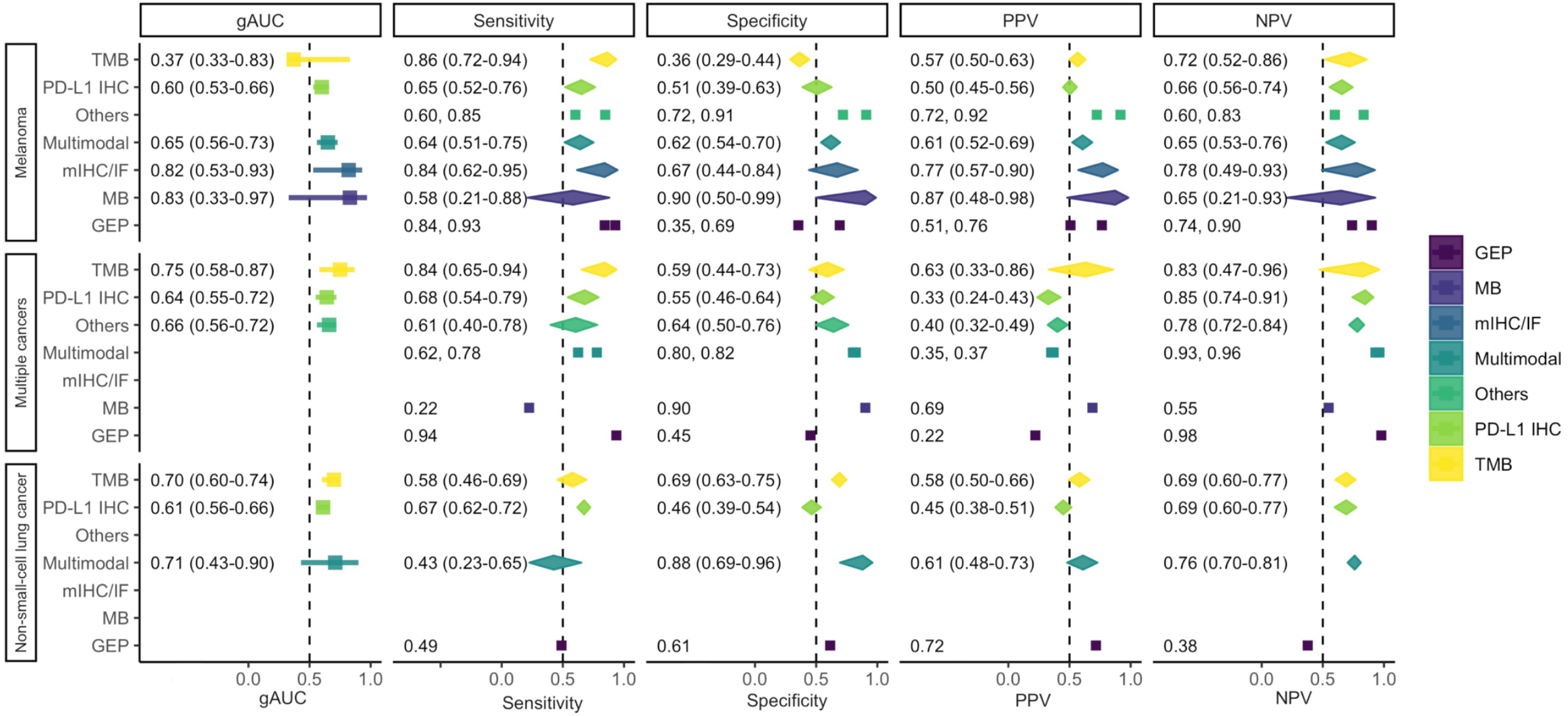
Meta-analyzed estimates for biomarkers within the three most frequently reported cancer groups. The sensitivity, specificity, positive predictive value (PPV) and negative predictive value (NPV) were meta-analyzed with false positive rate (FPR), false negative rate (FNR), false omission rate (FOR) and false discovery rate (FDR) using the linear regression approach with mixed effects implemented in R *mada* package^55^. The value for extrapolated area under the curve (gAUC) was calculated from sROC curves fitted for each biomarker within each cancer type using the Rutter-Gastonis method. The bars on gAUC facets represent 95% bootstrapped confidence intervals (95% CI).

**Fig 2.**
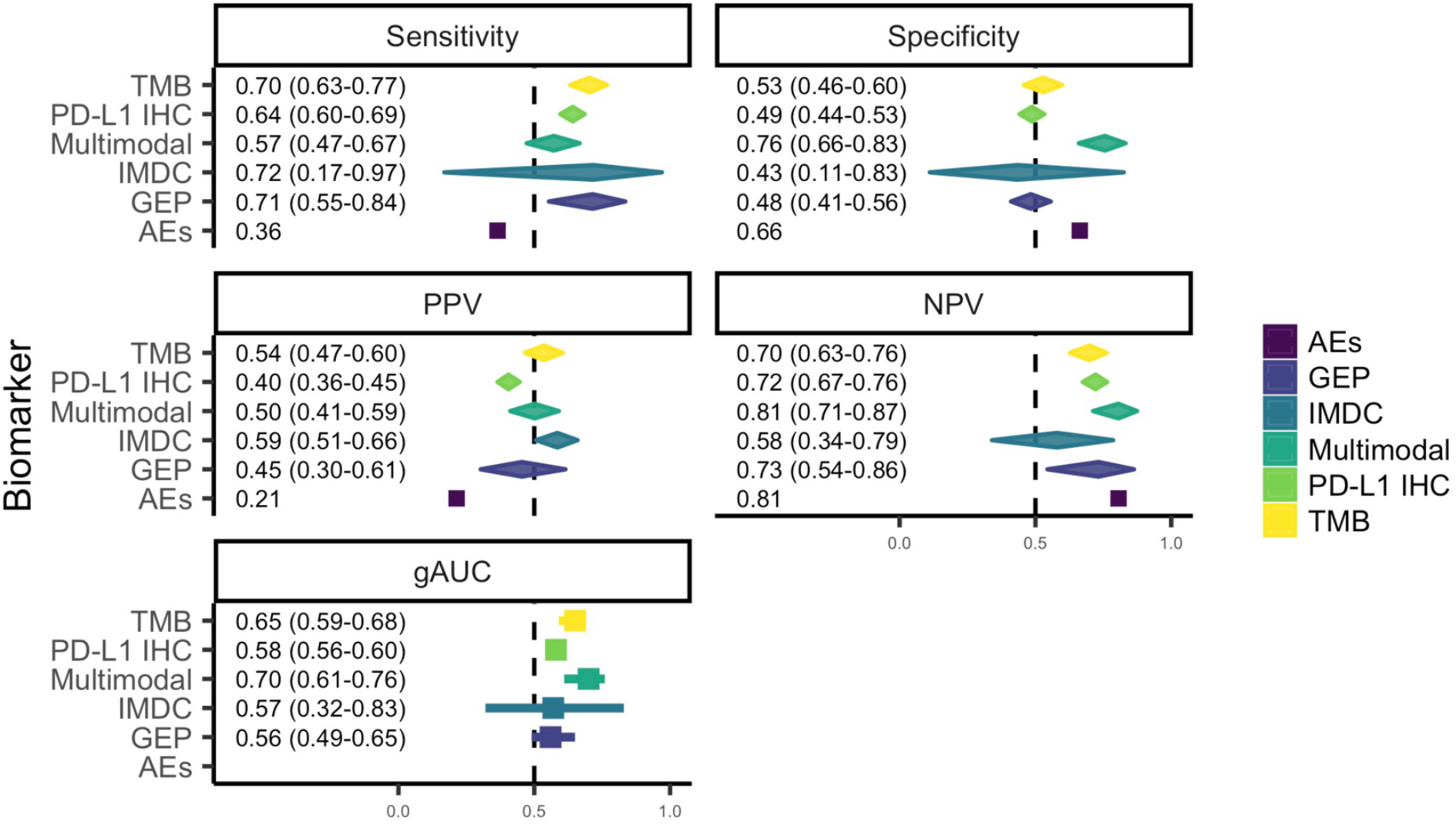
Meta-analyzed estimates of biomarkers with across cancers with pooled sample sizes greater than 500. The meta-analyzed sensitivity, specificity, positive predictive value (PPV), negative predictive value (NPV) and extrapolated area under the curve (gAUC) across cancers with a pooled sample size > 500. The sensitivity, specificity, PPV and NPV were meta-analyzed with false positive rate (FPR), false negative rate (FNR), false omission rate (FOR) and false discovery rate (FDR) using the linear regression approach with mixed effects implemented in R *mada* package^55^. The values for gAUC was calculated from sROC curves fitted for each biomarker within each cancer type using the Rutter-Gastonis method. The bars on gAUC facets represent 95% bootstrapped confidence intervals (95% CI).

### Non-small-cell lung cancer (NSCLC)

In studies of NSCLC, PD-L1 expression (N=26), tumor mutational burden (TMB) (N=9), Gene expression profiles (GEP) (N=1) and multimodal biomarkers (N=3) were investigated for prognostic biomarker potential (Supplementary Table 5). TMB and PD-L1 expression classified responders better than chance (*P*<.05 for both) (Fig. 1). The gAUC estimates for TMB, PD-L1 expression and multimodal biomarker were 0.70 (*95% CI:* 0.60-0.74), 0.61 (*95% CI:* 0.56-0.66) and 0.71 (*95% CI:* 0.43-0.90), respectively. GEP was not meta-analyzed because only a single study was available, and this study classified responders with a sensitivity and specificity of 0.49 and 0.61, respectively.

### Melanoma

In studies of melanoma, fluorescent multiplex immunohistochemistry assays (mIHC/IF) (N=2, global area under the receiver operating characteristic curve (gAUC)= 0.82, *95% CI:* 0.53-0.93), PD-L1 immunohistochemistry (IHC) (N=14, gAUC= 0.60, *95% CI:* 0.53-0.66) and multimodal biomarkers (N=2, gAUC= 0.65, *95% CI:* 0.56-0.73) were significantly more accurate than random assignment (gAUC=0.50) (*P*<.05 for all). Conversely TMB had the lowest gAUC of 0.37 (*95% CI:* 0.33-0.83, N=4) (Fig. 1). Furthermore, bootstrap hypothesis testing showed that the gAUC estimate for TMB was significantly lower than PD-L1 IHC, multimodal biomarkers, mIHC/IF and microbiome (Supplementary Fig. 2). The three most accurate biomarkers had both similar sensitivities in capturing responders and misclassification rate (Fig. 1). The greatest specificity estimate value of 0.90 was observed for microbiome (*95% CI:* 0.50-0.99). In addition to microbiome, only multimodal biomarker captured more non-responders than 0.50 (specificity=0.62, *95% CI:* 0.54-0.70).

### Biomarker efficacy across cancers

The prognostic efficacy of each biomarker was evaluated for 1) the five biomarkers with greatest cohort sizes (i.e., PD-L1 IHC, TMB, adverse events of special interest (AEs), International Metastatic RCC Database Consortium risk score (IMDC)^29,30^, GEP and multimodal biomarkers), 2) multimodal and mIHC/IF biomarkers and 3) for specific targeted somatic mutations. Because presence of a targeted somatic mutation can be representative of either an increased or reduced likelihood to benefit from ICI treatment, each mutation was meta-analyzed evaluating both possibilities (Fig. 3).

**Fig 3.**
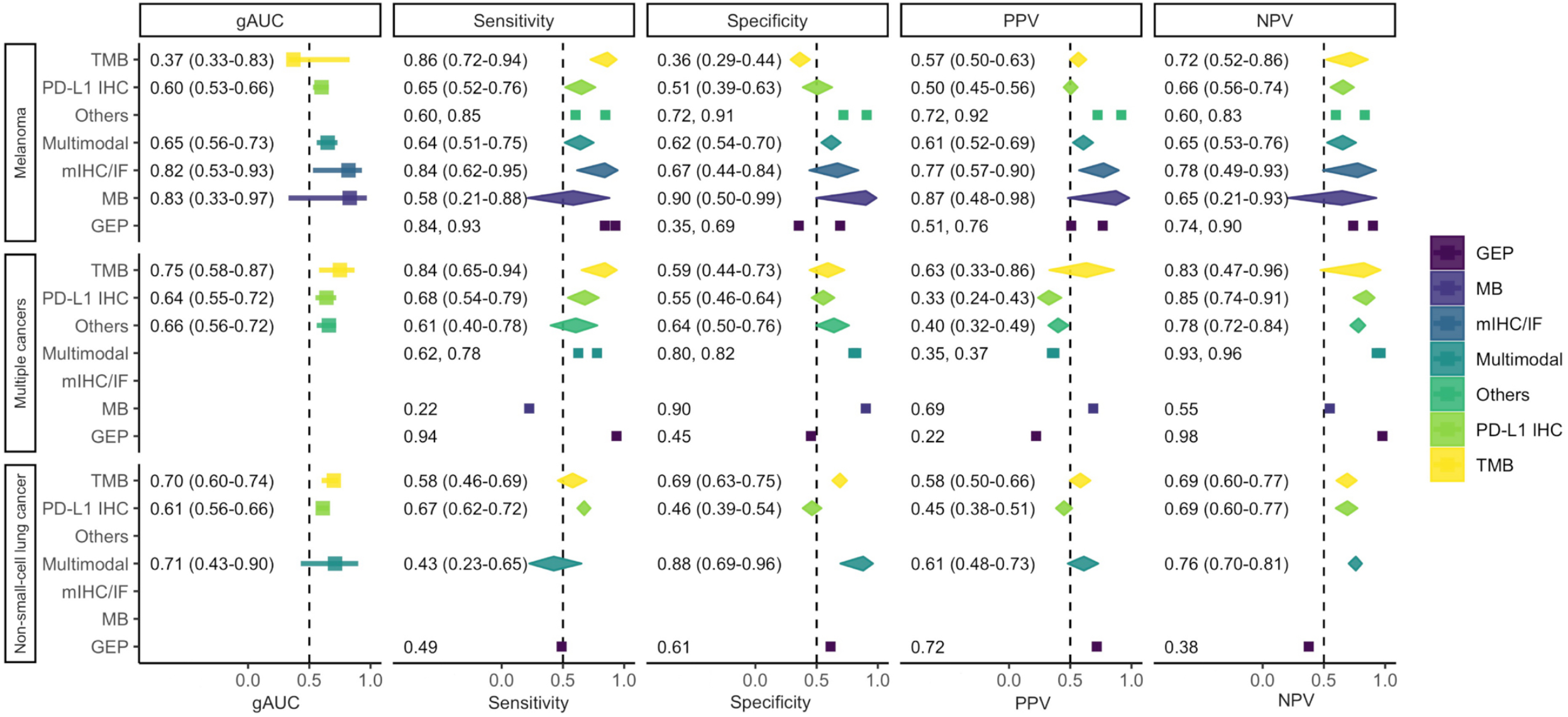
Meta-analyzed estimates for targeted somatic mutations. Meta-analyzed sensitivity, specificity, positive predictive value (PPV), negative predictive value (NPV) and extrapolated area under the curve (gAUC) values. The sensitivity, specificity, PPV and NPV were meta-analyzed with false positive rate (FPR), false negative rate (FNR), false omission rate (FOR) and false discovery rate (FDR) using the linear regression approach with mixed effects implemented in R *mada* package^55^. The values for gAUC was calculated from sROC curves fitted for each biomarker within each cancer type using the Rutter-Gastonis method. The bars on the gAUC facets represent 95% bootstrapped confidence intervals (95% CI).

### Frequently explored biomarkers

Across all evaluated cancer types, the performance for biomarkers with cohort sizes > 500 were investigated. These were PD-L1 IHC (N=13,601 subjects), AEs (N=1,747), TMB (N=1,724), GEP (N=775), multimodal biomarker (N=559) and IMDC (N=547). All biomarkers other than adverse events were meta-analyzed, because adverse events all came from a single published report^23^. The greatest and lowest gAUC values were observed for multimodal biomarker (gAUC=0.70, *95% CI:* 0.61-0.76) and GEP (gAUC=0.56, *95% CI:* 0.49-0.65), respectively. Multimodal biomarkers and TMB were significantly more accurate than PD-L1 IHC (*P*=.04) (Supplementary Fig. 6). GEP was most sensitive to responders (sensitivity=0.71, *95% CI*: 0.55-0.84) whereas AEs were the least sensitive (sensitivity=0.36). Multimodal biomarker and TMB had sensitivity estimates of 0.57 (*95% CI:* 0.47-0.67) and 0.70 (*95% CI:* 0.63-0.77), respectively. In addition to having the second greatest estimated sensitivity, accurate classification of non-responders by TMB (Specificity=0.53, *95% CI:* 0.46-0.60) was second only to multimodal biomarker (Specificity=0.76, *95% CI:* 0.66-0.83). The complete results are shown in Fig. 2.

### Multimodal and mIHC/IF biomarkers

Five studies were available that investigated multimodal biomarkers, and three studies were available that investigated mIHC/IF. The number of individuals for each biomarker was 559 and 177, respectively. The meta-analyzed gAUC estimate indicated that both biomarkers performed better than random chance (gAUC > 0.50) for discriminating treatment responders (*P*<.05). However, the gAUC estimate for multimodal biomarkers (gAUC=0.70, *95% CI:* 0.61-0.76) was not significantly different than the mIHC/IF estimate (gAUC=0.81, *95% CI:* 0.58-0.91) (*P*>.05). mIHC/IF was a more sensitive biomarker (sensitivity=0.85, *95% CI:* 0.68-0.94) compared to multimodal (sensitivity= 0.57, *95% CI:* 0.47-0.67). In addition, no significant differences were observed in the gAUC values of either of these biomarkers and TMB (Table 2) (*P*>.05). However, multimodal biomarkers classified patient response better than PD-L1 IHC (*P*=.04) while mIHC/IF failed to reach statistical significance for improvement over PD-L1 IHC (*P*=.06) (Supplementary Fig. 6).

### Targeted somatic mutations

None of the targeted mutations were significantly better than random chance at classifying responders (gAUC=0.50) (Fig. 3, Supplemental Table 8). A single study^31^ investigating alterations in DNA damage response genes as biomarker for individuals with urothelial cancer, had the greatest observed sensitivity of 0.90, but its specificity was only 0.50. Although only based on a single study^32^ of 38 individuals with melanoma, using wildtype for *BRCA2* as a biomarker resulted in a sensitivity of 0.71 and but only had a specificity of 0.06, indicating it resulted in too many false positives for predicting who is likely to respond to treatment.

### Microbiome

To our knowledge, the microbiome has been assessed for association with response to anti-PD-1/PD-L1 treatment in only three studies, two of which were in patients with melanoma. However, as described above, the microbiome was highly specific (specificity=0.90, *95% CI:* 0.50-0.99), but was poorly sensitive (sensitivity=0.58, *95% CI*: 0.21-0.88). Similarly, the specificity and sensitivity for the study^33^ in mixed cancer cases was 0.90 and 0.22, respectively. In addition to the microbiome, multimodal biomarkers were the only other biomarker that accurately captured significantly more non-responders than expected by random chance (specificity=0.60, *95% CI:* 0.54-0.70).

## Discussion

ICIs targeting PD-1/PD-L1 have resulted in breakthrough treatments for a multitude of cancers, and the impact this class of drugs has on cancer treatment cannot be overstated. Despite these successes, only 24% (95% CI= 21%-28%) of patients respond to these treatments^2^. A variety of different biomarkers have been considered; however, no consensus exists regarding which of these biomarkers is capable of or has potential to be clinically useful. This broad-based meta-analysis addresses the unmet need of characterizing commonly considered biomarkers for ICI treatment response in a variety of cancer types. To our knowledge, this is the most comprehensive study of published biomarkers for ICI response performed to date, and builds off of a recent meta-analysis performed by Lu et al.^34^. Here we extend that study in several ways by expanding the number of studies from 46 to 95, the number of patients from 8,135 to 18,978, investigating whether the microbiome or immune mediated adverse events show evidence of biomarker potential, and evaluating whether the AUCs of biomarker performance are statistically different. We also implemented bivariate linear mixed models which has been shown to provide more accurate estimates rather than estimating sensitivity and specificity separately, which can yield misleading results^34^. Lastly, we use a summary receiver operating characteristic curve (sROC) approach which addresses limitations for comparing binary tests across different studies and statistical thresholds^35^.

We found that PD-L1 IHC, multimodal biomarkers, and mIHC/IF discriminate responders significantly better than random assignment across all cancer types as well as specific cancer types—melanoma and non-small-cell lung cancer (gAUC >0.5). TMB discriminated responders better than random assignment across all cancer types and non-small-cell lung cancer but not within melanoma patients. These findings are consistent with the results previously published by Lu et al^34^. In addition, Lu et al. reported that mIHC/IF and multimodal biomarkers performed better than TMB and PD-L1 expression; however, here the performance of these biomarkers was determined to be similar. Though the methodology is similar between these two studies, some key differences may have led to discrepancies between these two studies. One of the advantages to this study is the utilization of bootstrapping to test for statistical differences in the biomarker accuracies. Similar to Lu et al^34^, we observed that the gAUC for mIHC/IF (gAUC=0.81, *95% CI*: 0.58-0.91) was higher than that of TMB (gAUC= 0.65, *95% CI*: 0.59-0.68) and PD-L1 IHC (gAUC=0.58, *95% CI*: 0.56-0.60); however, our approach did not find that these biomarkers were significantly different (*P*>.05). On the other hand, multimodal biomarkers classified patients more accurately than PD-L1 IHC (Δ gAUC= 0.12, *P*=.04). Another notable difference between the two studies is the definition of clinical response used. Here, in an effort to maintain as much consistency in reported outcomes across studies, we only considered studies that reported objective response rate (ORR), CB and PFS (6 months); whereas overall survival and different PFS time points were also considered in Lu et al.^34^ While some clinical studies have reported better efficacy of TMB than PD-L1 IHC^36^, other studies have found similar performances between the two.^34,37^ We found that TMB demonstrated better performance (gAUC= 0.65, *95% CI*: 0.59-0.68) than PD-L1 IHC (gAUC=0.58, *95% CI*: 0.56-0.60) (*P*<.05).

Because clinical treatment decisions will be made on an individual patient level, we investigated whether the weight of evidence supporting the use of a given biomarker differed across cancer types. Cancer type is one of the sources of threshold and performance heterogeneity of biomarkers between clinical studies. For example, Zhang et al. reported greater treatment response in PD-L1 positive subgroups in melanoma, non-small-cell lung cancer and renal cell carcinoma, but not in ovarian cancer, bladder cancer and multiple, combined cancer types^2^. Here, we found that, in melanoma, biomarkers mIHC/IF (gAUC= 0.82, *95% CI:* 0.53-0.93) and PD-L1 IHC (gAUC= 0.60, *95% CI:* 0.53-0.66) were not significantly different from one another (*P*>.05); whereas these were marginally significant across all cancer types (*P*=.06) with gAUCs of 0.70 (*95% CI:* 0.61-0.76) and 0.58 (*95% CI:* 0.56-0.60) for mIHC/IF and PD-L1 expression, respectively. The mIHC/IF PD-L1 expression level had better predictive value than single PD-L1 expression IHC assays with a gAUC improvement of 0.23 with marginal significance (*P*=.06) across all cancer types. Tumor type, observing pathologist, assay type and lack of broader assessment of tumor microenvironments assessment have all been reported to negatively affect the efficacy of PD-L1 IHC^34^. TMB performance in melanoma varied greatly (gAUC= 0.37, *95% CI:* 0.33-0.83), and was more homogeneous in non-small-cell lung cancer (gAUC= 0.70, *95% CI:* 0.60-0.74) and across all cancer types (gAUC=0.65, *95% CI:* 0.59-0.68). Given these findings, there are likely other factors than just cancer type that contribute to heterogeneity in performance and additional studies are needed to identify other factors that may affect biomarker performance. However, meta-analyses provide relevant summaries of results and aid in understanding relationships and heterogeneity in the context of multiple studies.^38^

Two of the more novel putative biomarkers of ICI treatment response included here are AEs and the microbiome. The AEs biomarker is distinct from the others in that it is ascertained after treatment has begun, limiting its potential use as a pre-treatment biomarker. However, if determined to be effective it could still serve as a leading indicator of response, providing opportunities to modify or enhance treatment. To our knowledge, AEs and response to ICIs have only been explored in patients with urothelial cancer.^23^ Despite being significantly associated with ORR,^23^ AEs were found to be the least sensitive for detecting responding individuals (sensitivity= 0.36). Defining this biomarker with a different criterion in different cancer types may produce different results. To this point, the microbiome had a greater meta-analyzed sensitivity estimate (sensitivity= 0.58, 95% *CI=* 0.21-0.88) for studies in melanoma patients than the study conducted in multiple cancers (sensitivity= 0.22). In addition, the microbiome was highly specific for detecting non-responders in patients with melanoma (specificity=0.90, *95% CI:* 0.50-0.99) and multiple cancers (specificity=0.90), suggesting that it may have potential use as a biomarker if used in combination with more sensitive biomarkers. The microbiome results here also justify the investigation of this biomarker in other cancer types and potentially for use with other classes of ICIs. The results of these constituent studies have not been prospectively validated in an independent cohort. However, these meta-analyses provide what could be considered an indication of the discriminatory potential for response to ICI treatment.

Multimodal and TMB outperformed PD-L1 IHC when all cancers were combined (*P*<.05), with marginal improvements also seen for TMB and the microbiome over PD-L1 IHC (*P*<.10). mIHC/IF, PD-L1 IHC and multimodal biomarkers adequately captured responders and non-responders in patients with melanoma, non-small-cell lung cancer and across combined cancer types better than random chance (*P*<.05). Although TMB discriminated responders and non-responders similar to other biomarkers for patients with non-small cell lung cancer and when cancers were combined, TMB for patients with melanoma produced mixed results.

Overall, biomarker performance greatly depends on cancer type and studies vary widely based on their methods; however, several biomarkers show promise for predicting treatment response to ICIs and additional standardization will needed to maximize their clinical utility.

## Methods

### Literature Search and Inclusion Criteria

PubMed and Google Scholar were used to search for peer-reviewed studies focused on anti-PD-1/anti-PD-L1 therapies and biomarkers. Keywords used to search for studies included: “anti-PD-1/anti-PD-L1 therapies and tumor mutational burden”, “anti-PD-1/anti-PD-L1 therapies and adverse events”, “anti-PD-1/anti-PD-L1 therapies and biomarkers” and “biomarkers for immune check point inhibitors”. Studies were selected based on accessibility and availability of summary level or patient level data based on clinical outcomes and predictive biomarkers.

### Data

For each study, the title, publication year, treatment, type of cancer, biomarker and clinical outcome details were documented. CB, ORR and PFS were considered as clinical outcomes. The clinical outcomes and biomarker thresholds were accepted as defined in each study. The following metrics for biomarker performance were calculated: sensitivity, specificity, balanced accuracy (BA), positive predictive value (PPV), negative predictive value (NPV), false omission rate (FOR), false negative rate (FNR), and false discovery rate (FDR). Each of these metrics can be calculated from a 2×2 contingency table where counts of individuals meeting the criterion for being positive or negative for the biomarker under consideration and being positive or negative for the clinical outcome (Supplemental Table 1). Only studies that provided either individual counts for each cell in the 2×2 table or the necessary individual level information to complete the 2×2 table were included. Studies that did not propose a threshold or cutoff value for the biomarker were excluded unless participant level data was available from which a 2×2 table could be developed.

#### Patient and Public Involvement

Patients were not involved in performing this research. All patient data was reported separately in the relevant peer reviewed publications which can be found in Supplemental Table 2.

#### Biomarkers

Across all included studies, the following biomarkers were investigated (Supplementary Table 3). Some studies evaluated multiple biomarker thresholds and only the results with the threshold resulting in the highest reported balanced accuracy were included for analysis. Biomarkers with less than three reported studies or a total combined patient count =< 500 were combined to form a group referred to as “other” biomarkers (Supplemental Table 4). The most frequently observed biomarkers are described below:

#### PD-L1 protein expression

PD-L1 protein expression measured on tumor cells, immune cells or both, were included. For each study, the investigators selected an expression threshold to compare observed clinical responses. Patients with PD-L1 expression greater than the threshold were expected to be more likely to respond to treatment. PD-L1 expression was further divided into assay type subsets of IHC and mIHC/IF.

#### Tumor mutational burden (TMB)

TMB refers to the number of somatic DNA mutations across the tumor genome. Since the early studies of TMB, many variations of this biomarker have been studied. Tumor mutational burden has been quantified based on non-synonymous single nucleotide variants^32,39^, frameshift mutations^40^ and circulating tumor DNA^41^. Studies calculating TMB from whole exome or whole genome sequencing were included, as well. Median TMB was a commonly reported threshold for assessing response to ICIs, although some studies investigated alternative thresholds. The TMB threshold defined by the authors of each study was accepted for the analysis here, with the exception of Hugo et al.^32^, which did not report a threshold and the median TMB was used. For all studies, TMB was evaluated to determine if being above the threshold was indicative of increased likelihood of response to treatment.

#### Targeted somatic mutations

Many studies have investigated whether somatic mutations in specific genes could serve as biomarkers for response to ICIs. We focused on several commonly reported mutations in the following genes: B-Raf proto-oncogene, serine/threonine kinase (*BRAF*)^5,42–47^, progesterone receptor (*PR*)^9,48^, estrogen receptor alpha (ERa)^9,48^, KRAS proto-oncogene, GTPase (*KRAS*)^32,44,46^, *KRAS* and *NRAS genes* (KRAS/NRAS)^45,49^ and tumor protein p53. Some of these mutations are more common in certain cancers. Therefore, we presented this information in two ways: (i) individual gene performance across all reported cancers, and (ii) we meta-analyzed each of these genes to provide an overall assessment for how well individual targeted somatic mutations discriminate patient response to ICIs.

#### Adverse events of Special Interest (AEs)

Unlike other biomarkers, which are assessed prior to treatment initiation, adverse events of special interest (AEs) were observed after the administration of ICIs, but prior to the determination of a clinical response. AEs included a variety of events including autoimmune events, rash, diarrhea, and others^23^. These data were previously reported in Maher et al. which combined data from five trials submitted to the US Food and Drug Administration (FDA)^23^ and we previously reported the discriminatory potential for these biomarkers^50^. AEs were evaluated to determine if their occurrence was indicative of increased likelihood of response to treatment.

#### Gene expression profiles (GEP)

GEPs classify responders based on arrays of inflammatory-, immune checkpoint and onco-genes. The studies focused on expression of indoleamine 2,3-dioxygenase (*IDO*) and *FOXP3* as well as angiogenesis gene signature (Angio), T-cell effector gene signature (Teff) and innate anti-PD-1 resistance gene signature (IPRES). Significant associations between high expression of *FOXP3* and *IDO* with clinical efficacy were reported^22,32,40,43^. Presence of Angio and Teff signatures along with absence of IPRES signature were also associated with clinical efficacy^22,32,40^.

#### Microbiome

Individuals with gut and oral commensal microbiomes that promote anti-tumor immunity have been shown to benefit more from ICI treatments than others^51,52^. Conversely, down-regulation of these microbiomes by antibiotics has been linked to worse treatment responses^33^. Furthermore, pre-treatment microbiome profiles have been reported to discriminate responders and non-responders^51^.

#### Multimodal biomarkers

All of the biomarkers described above capture distinct information about the tumor microenvironment (e.g. TMB, PD-L1 IHC) and the host (e.g. GEP, Microbiome). Some studies have combined TMB with PD-L1 and GEP and PD-1 IHC with PD-L1 IHC for a comprehensive assessment of the tumor and clinical efficacy.

#### Clinical responses

Either ORR, CB or PFS were used to represent clinical response to treatment. If ORR was not available, in order of preference, CB and six-months PFS were used. These responses were determined using response evaluation criteria in solid tumors (RECIST), immune-related response criteria (irRC), or modified response evaluation criteria in solid tumors (mRECIST)^53,54^ by investigator assessment or independent review. If a response was evaluated using multiple tumor criteria or by multiple assessors, the data was averaged to the nearest integer.

### Statistical Analysis

#### Meta-analysis

A PRISMA checklist detailing quality control metrics for reporting meta-analyses used in this study are presented in the Supplemental Material. Biomarker performance metrics (Supplementary Table 1) were calculated for each study and various groups were meta-analyzed for comparison using the R packages, *mada*^55^ and *meta4diag*^56^. Meta-analyses were conducted on data subsets to determine (i) discriminatory potential for each biomarker across multiple cancer types (Supplemental Table 4), and (ii) discriminatory potential for each biomarker for each cancer type. Binary test outcomes, such as sensitivity and specificity, rely on a threshold for determining the optimal test performance. However, this threshold often creates a tradeoff between certain values. In addition, simply averaging values across studies with different thresholds can confound results^57^. To address this, we implemented the sROC approach^35,55^. Bivariate analyses were performed using a linear mixed model with random effects and evaluated specificity, positive predictive value and negative predictive value with false negative rate, false omission rate and false discovery rate, respectively. A minimum size of three studies was required in order to perform each meta-analysis. Both partial AUCs (pAUC) and global AUCs (gAUCs) were calculated. While the pAUC estimates the discriminatory ability of the observed data, gAUC estimates are calculated from the extrapolated bivariate models. The confidence intervals for both AUC estimates were estimated based on a bootstrapped sample of 10,000 iterations^58^.

#### Bootstrap hypothesis testing

Biomarkers were compared pairwise to determine if the observed gAUC values were significantly different (*P*<.05). Null distributions were generated for the difference in the gAUC estimates between each pairwise biomarker by calculating the difference in the gAUC values assigned to a random mix of the studies focused on biomarkers of interest. Each distribution consisted of 10,000 such values, and the probability of differences observed between the biomarker estimates were calculated as the number of observed gAUC values greater than the gAUC from the null distribution, divided by 10,000.

## Data Availability

All available data is provided in the Supplementary Material and through the publications referenced in the Supplementary Material.

## Declarations

### Funding

D.M.R. and A.M. were supported in part by the Clinical and Translational Science Collaborative of Cleveland, (KL2TR002547) from the National Center for Advancing Translational Sciences (NCATS) component of the NIH.

### Competing Interests

D.M.R. has stock and other ownership interests in Interpares Biomedicine. He has served in a consultant and advisory role for Pharmazam and Clariifi. He has received research funding from Novo Nordisk and has intellectual property related to the detection of liver cancer. H.L.M. has stock and other ownership interests in Cancer Genetics, Interpares Biomedicine, Clariifi, and serves as a consultant or advisory role for Gentris, Cancer Genetics, Saladax Biomedical, National Institutes of Health/National Cancer Institute, Admera Health, eviCore healthcare, Pharmazam, VieCure.

### Disclaimer

The funders had no role in study design, data collection and analysis, interpretation, decision to publish, or preparation of the manuscript; or any aspect of the study.

### Contributions

A.M. performed analyses and wrote the manuscript. K.S., performed analyses and reviewed the manuscript. S.K. and H.L.M. reviewed the manuscript and provided feedback.

D.M.R. designed and supervised the study, wrote and reviewed the manuscript.

## Notes

### Author Declarations

All of the data was publically available from peer reviewed papers and no IRB approval was required.

## References

1. Hao, C. et al. Efficacy and safety of anti-PD-1 and anti-PD-1 combined with anti-CTLA-4 immunotherapy to advanced melanoma: A systematic review and meta-analysis of randomized controlled trials. Medicine (United States) vol. 96 (2017).

2. Zhang, T. et al. The efficacy and safety of anti-PD-1/PD-L1 antibodies for treatment of advanced or refractory cancers: A meta-analysis. Oncotarget 7, 73068–73079 (2016).

3. Rosenberg, J. E. et al. Atezolizumab in patients with locally advanced and metastatic urothelial carcinoma who have progressed following treatment with platinum-based chemotherapy: A single-arm, multicentre, phase 2 trial. The Lancet 387, 1909–1920 (2016).

4. Kato, K. et al. Nivolumab versus chemotherapy in patients with advanced oesophageal squamous cell carcinoma refractory or intolerant to previous chemotherapy (ATTRACTION-3): a multicentre, randomised, open-label, phase 3 trial. Lancet Oncol. 20, 1506–1517 (2019).

5. Weber, J. S. et al. Nivolumab versus chemotherapy in patients with advanced melanoma who progressed after anti-CTLA-4 treatment (CheckMate 037): A randomised, controlled, open-label, phase 3 trial. Lancet Oncol. 16, 375–384 (2015).

6. Powles, T. et al. Atezolizumab versus chemotherapy in patients with platinum-treated locally advanced or metastatic urothelial carcinoma (IMvigor211): a multicentre, open-label, phase 3 randomised controlled trial. The Lancet 391, 748–757 (2018).

7. Rittmeyer, A. et al. Atezolizumab versus docetaxel in patients with previously treated non-small-cell lung cancer (OAK): a phase 3, open-label, multicentre randomised controlled trial. The Lancet 389, 255–265 (2017).

8. Taube, J. M. et al. Association of PD-1, PD-1 ligands, and other features of the tumor immune microenvironment with response to anti-PD-1 therapy. Clin. Cancer Res. 20, 5064–5074 (2014).

9. Dirix, L. Y. et al. Avelumab, an anti-PD-L1 antibody, in patients with locally advanced or metastatic breast cancer: A phase 1b JAVELIN solid tumor study. Breast Cancer Res. Treat. 167, 671–686 (2018).

10. Kefford, R. et al. Clinical efficacy and correlation with tumor PD-L1 expression in patients (pts) with melanoma (MEL) treated with the anti-PD-1 monoclonal antibody MK-3475. J. Clin. Oncol. 32, 3005–3005 (2014).

11. Passiglia, F. et al. PD-L1 expression as predictive biomarker in patients with NSCLC: A pooled analysis. Oncotarget 7, 19738–19747 (2016).

12. Goodman, A. M. et al. Tumor mutational burden as an independent predictor of response to immunotherapy in diverse cancers. Mol. Cancer Ther. 16, 2598–2608 (2017).

13. Ready, N. et al. First-Line Nivolumab Plus Ipilimumab in Advanced Non-Small-Cell Lung Cancer (CheckMate 568): Outcomes by Programmed Death Ligand 1 and Tumor Mutational Burden as Biomarkers. J. Clin. Oncol. Off. J. Am. Soc. Clin. Oncol. 37, 992–1000 (2019).

14. Gandara, D. R. et al. Blood-based tumor mutational burden as a predictor of clinical benefit in non-small-cell lung cancer patients treated with atezolizumab. Nat. Med. 24, 1441–1448 (2018).

15. Hellmann, M. D. et al. Tumor Mutational Burden and Efficacy of Nivolumab Monotherapy and in Combination with Ipilimumab in Small-Cell Lung Cancer. Cancer Cell 33, 853-861.e4 (2018).

16. Marabelle, A. et al. Association of tumour mutational burden with outcomes in patients with advanced solid tumours treated with pembrolizumab: prospective biomarker analysis of the multicohort, open-label, phase 2 KEYNOTE-158 study. Lancet Oncol. 21, 1353–1365 (2020).

17. Rizvi, H. et al. Molecular determinants of response to anti-programmed cell death (PD)-1 and anti-programmed death-ligand 1 (PD-L1) blockade in patients with non-small-cell lung cancer profiled with targeted next-generation sequencing. J. Clin. Oncol. 36, 633–641 (2018).

18. Hellmann, M. D. et al. Genomic Features of Response to Combination Immunotherapy in Patients with Advanced Non-Small-Cell Lung Cancer. Cancer Cell 33, 843-852.e4 (2018).

19. Havel, J. J., Chowell, D. & Chan, T. A. The evolving landscape of biomarkers for checkpoint inhibitor immunotherapy. Nature Reviews Cancer vol. 19133–150 (2019).

20. Borghaei, H. et al. Nivolumab versus docetaxel in advanced nonsquamous non-small-cell lung cancer. N. Engl. J. Med. 373, 1627–1639 (2015).

21. Forde, P. M. et al. Neoadjuvant PD-1 blockade in resectable lung cancer. N. Engl. J. Med. 378, 1976–1986 (2018).

22. Socinski, M. A. et al. Atezolizumab for first-line treatment of metastatic nonsquamous NSCLC. N. Engl. J. Med. 378, 2288–2301 (2018).

23. Maher, V. E. et al. Analysis of the Association Between Adverse Events and Outcome in Patients Receiving a Programmed Death Protein 1 or Programmed Death Ligand 1 Antibody. J. Clin. Oncol. Off. J. Am. Soc. Clin. Oncol. 37, 2730–2737 (2019).

24. Bellmunt, J. et al. Pembrolizumab as Second-Line Therapy for Advanced Urothelial Carcinoma. N. Engl. J. Med. 376, 1015–1026 (2017).

25. Massard, C. et al. Safety and Efficacy of Durvalumab (MEDI4736), an Anti-Programmed Cell Death Ligand-1 Immune Checkpoint Inhibitor, in Patients With Advanced Urothelial Bladder Cancer. J. Clin. Oncol. Off. J. Am. Soc. Clin. Oncol. 34, 3119–25 (2016).

26. Kambayashi, Y., Fujimura, T., Hidaka, T. & Aiba, S. Biomarkers for Predicting Efficacies of Anti-PD1 Antibodies. Front. Med. 6, 174 (2019).

27. Topalian, S. L., Taube, J. M., Anders, R. A. & Pardoll, D. M. Mechanism-driven biomarkers to guide immune checkpoint blockade in cancer therapy. Nature Reviews Cancer vol. 16 275–287 (2016).

28. Janjigian, Y. Y. et al. 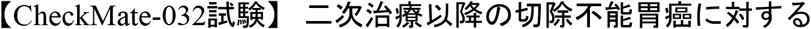 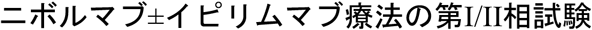. J. Clin. Oncol. 36, 2836–2844 (2018).

29. Tykodi, S. S. et al. First-line pembrolizumab (pembro) monotherapy in advanced clear cell renal cell carcinoma (ccRCC): Updated results for KEYNOTE-427 cohort A. J. Clin. Oncol. 37, 4570–4570 (2019).

30. Motzer, R. J. et al. Nivolumab plus Ipilimumab versus Sunitinib in advanced renal-cell carcinoma. N. Engl. J. Med. 378, 1277–1290 (2018).

31. Teo, M. Y. et al. Alterations in DNA damage response and repair genes as potential marker of clinical benefit from PD-1/PD-L1 blockade in advanced urothelial cancers. J. Clin. Oncol. 36, 1685–1694 (2018).

32. Hugo, W. et al. Genomic and Transcriptomic Features of Response to Anti-PD-1 Therapy in Metastatic Melanoma. Cell 165, 35–44 (2016).

33. Routy, B. et al. Gut microbiome influences efficacy of PD-1-based immunotherapy against epithelial tumors. Science 359, 91–97 (2018).

34. Lu, S. et al. Comparison of Biomarker Modalities for Predicting Response to PD-1/PD-L1 Checkpoint Blockade: A Systematic Review and Meta-analysis. JAMA Oncology vol. 5 1195–1204 (2019).

35. Holling, H., Böhning, W. & Böhning, D. Meta-analysis of diagnostic studies based upon SROC-curves: A mixed model approach using the Lehmann family. Stat. Model. 12, 347–375 (2012).

36. Boumber, Y. Tumor mutational burden (TMB) as a biomarker of response to immunotherapy in small cell lung cancer. Journal of Thoracic Disease vol. 10 4689–4693 (2018).

37. Aggen, D. H. & Drake, C. G. Biomarkers for immunotherapy in bladder cancer: A moving target. Journal for ImmunoTherapy of Cancer vol. 5 94 (2017).

38. Greco, T., Zangrillo, A., Biondi-Zoccai, G. & Landoni, G. Meta-analysis: pitfalls and hints. Heart Lung Vessels 5, 219–25 (2013).

39. Rizvi, N. A. et al. Mutational landscape determines sensitivity to PD-1 blockade in non-small cell lung cancer. Science 348, 124–128 (2015).

40. McDermott, D. F. et al. Clinical activity and molecular correlates of response to atezolizumab alone or in combination with bevacizumab versus sunitinib in renal cell carcinoma. Nat. Med. 24, 749–757 (2018).

41. Khagi, Y. et al. Hypermutated circulating tumor DNA: Correlation with response to checkpoint inhibitor–based immunotherapy. Clin. Cancer Res. 23, 5729–5736 (2017).

42. Robert, C. et al. Pembrolizumab versus ipilimumab in advanced melanoma (KEYNOTE-006): post-hoc 5-year results from an open-label, multicentre, randomised, controlled, phase 3 study. Lancet Oncol. 20, 1239–1251 (2019).

43. Hamid, O. et al. Final analysis of a randomised trial comparing pembrolizumab versus investigator-choice chemotherapy for ipilimumab-refractory advanced melanoma. Eur. J. Cancer 86, 37–45 (2017).

44. Overman, M. J. et al. 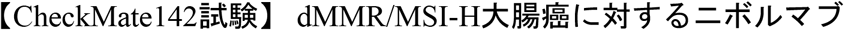 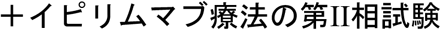. J. Clin. Oncol. 36, JCO.2017.76.990 (2018).

45. Le, D. T. et al. Phase II open-label study of pembrolizumab in treatment-refractory, microsatellite instability–high/mismatch repair–deficient metastatic colorectal cancer: KEYNOTE-164. in Journal of Clinical Oncology vol. 38 11–19 (American Society of Clinical Oncology, 2020).

46. Overman, M. J. et al. Nivolumab in patients with metastatic DNA mismatch repair-deficient or microsatellite instability-high colorectal cancer (CheckMate 142): an open-label, multicentre, phase 2 study. Lancet Oncol. 18, 1182–1191 (2017).

47. Postow, M. A. et al. Nivolumab and ipilimumab versus ipilimumab in untreated melanoma. N. Engl. J. Med. 372, 2006–2017 (2015).

48. Emens, L. A. Breast cancer immunotherapy: Facts and hopes. Clinical Cancer Research vol. 24 511–520 (2018).

49. Eng, C. et al. Atezolizumab with or without cobimetinib versus regorafenib in previously treated metastatic colorectal cancer (IMblaze370): a multicentre, open-label, phase 3, randomised, controlled trial. Lancet Oncol. 20, 849–861 (2019).

50. McLeod, H. L., Mariam, A., Schveder, K. A. & Rotroff, D. M. Assessment of adverse events and their ability to discriminate response to anti–PD-1/PD-L1 antibody immunotherapy. Journal of Clinical Oncology vol. 38 103–104 (2020).

51. Matson, V. et al. The commensal microbiome is associated with anti-PD-1 efficacy in metastatic melanoma patients. Science 359, 104–108 (2018).

52. Gopalakrishnan, V. et al. Gut microbiome modulates response to anti-PD-1 immunotherapy in melanoma patients. Science 359, 97–103 (2018).

53. El-Khoueiry, A. B. et al. Nivolumab in patients with advanced hepatocellular carcinoma (CheckMate 040): an open-label, non-comparative, phase 1/2 dose escalation and expansion trial. The Lancet 389, 2492–2502 (2017).

54. Rosenberg, J. E. et al. Atezolizumab in patients with locally advanced and metastatic urothelial carcinoma who have progressed following treatment with platinum-based chemotherapy: A single-arm, multicentre, phase 2 trial. The Lancet 387, 1909–1920 (2016).

55. Doebler, P. & Holling, H. Meta-Analysis of Diagnostic Accuracy with mada.

56. Guo, M. J. Package ‘meta4diag’ Title Meta-Analysis for Diagnostic Test Studies. (2018).

57. Gatsonis, C. et al. Meta-Analysis of Diagnostic and Screening Test Accuracy Evaluations: Methodologic Primer. 187, 271–281 (2006).

58. Riley, R. D. et al. Multivariate meta-analysis using individual participant data. Res. Synth. Methods 6, 157–174 (2015).

