## Supplemental Material for "Large-Scale Meta-Analysis of Potential Biomarkers for Treatment Response to Anti-PD-1/PD-L1 Immune Checkpoint Inhibitors"

### Supplementary Tables

**Supplementary Table 1. Formulas of accuracy metrics**

| **Metric** | **Formula** |
|---|---|
| False discovery rate (FDR) | $\frac{FP}{TP + FP}$ |
| False negative rate (FNR) | $\frac{FN}{TP + FN}$ |
| False omission rate (FOR) | $\frac{FN}{TN + FN}$ |
| False positive rate (FPR) | $\frac{FP}{TN + FP}$ |
| Negative predictive value | $\frac{TN}{TN + FN}$ |
| Positive predictive value | $\frac{TP}{TP + FP}$ |
| Sensitivity | $\frac{TP}{TP + FN}$ |
| Specificity | $\frac{TN}{TN + FP}$ |

**Supplementary Table 2. References**

| | References |
|---|---|
| 1 | Adams, S., P. Schmid, H. S. Rugo, E. P. Winer, D. Loirat, A. Awada, D. W. Cescon, H. Iwata, M. Campone, R. Nanda, R. Hui, G. Curigliano, D. Toppmeyer, J. O'Shaughnessy, S. Loi, S. Paluch-Shimon, A. R. Tan, D. Card, J. Zhao, V. Karantza, and J. Cortés. 2019. "Pembrolizumab Monotherapy for Previously Treated Metastatic Triple-Negative Breast Cancer: Cohort A of the Phase II KEYNOTE-086 Study." Annals of Oncology 30(3):397–404. |
| 2 | Antonia, Scott J., José A. López-Martin, Johanna Bendell, Patrick A. Ott, Matthew Taylor, Joseph Paul Eder, Dirk Jäger, M. Catherine Pietanza, Dung T. Le, Filippo de Braud, Michael A. Morse, Paolo A. Ascierto, Leora Horn, Asim Amin, Rathi N. Pillai, Jeffry Evans, Ian Chau, Petri Bono, Akin Atmaca, Padmanee Sharma, Christopher T. Harbison, Chen Sheng Lin, Olaf Christensen, and Emiliano Calvo. 2016. "Nivolumab Alone and Nivolumab plus Ipilimumab in Recurrent Small-Cell Lung Cancer (CheckMate 032): A Multicentre, Open-Label, Phase 1/2 Trial." The Lancet Oncology 17(7):883–95. |

| | |
|---|---|
| 3 | Armand, Philippe, Scott Rodig, Vladimir Melnichenko, Catherine Thieblemont, Kamal Bouabdallah, Gayane Tumyan, Muhit Özcan, Sergio Portino, Laura Fogliatto, Maria D. Caballero, Jan Walewski, Zafer Gulbas, Vincent Ribrag, Beth Christian, Guilherme Fleury Perini, Gilles Salles, Jakub Svoboda, Jasmine Zain, Sanjay Patel, Pei Hsuan Chen, Azra H. Ligon, Jing Ouyang, Donna Neuberg, Robert Redd, Arkendu Chatterjee, Arun Balakumaran, Robert Orlowski, Margaret Shipp, and Pier Luigi Zinzani. 2019. “Pembrolizumab in Relapsed or Refractory Primary Mediastinal Large B-Cell Lymphoma.” Pp. 3291–99 in Journal of Clinical Oncology. Vol. 37. American Society of Clinical Oncology. |
| 4 | Balar, Arjun V., Matthew D. Galsky, Jonathan E. Rosenberg, Thomas Powles, Daniel P. Petrylak, Joaquim Bellmunt, Yohann Loriot, Andrea Necchi, Jean Hoffman-Censits, Jose Luis Perez-Gracia, Nancy A. Dawson, Michiel S. van der Heijden, Robert Dreicer, Sandy Srinivas, Margitta M. Retz, Richard W. Joseph, Alexandra Drakaki, Ulka N. Vaishampayan, Srikala S. Sridhar, David I. Quinn, Ignacio Durán, David R. Shaffer, Bernhard J. Eigl, Petros D. Grivas, Evan Y. Yu, Shi Li, Edward E. Kadel, Zachary Boyd, Richard Bourgon, Priti S. Hegde, Sanjeev Mariathasan, Ann Christine Thåström, Oyewale O. Abidoye, Gregg D. Fine, and Dean F. Bajorin. 2017. “Atezolizumab as First-Line Treatment in Cisplatin-Ineligible Patients with Locally Advanced and Metastatic Urothelial Carcinoma: A Single-Arm, Multicentre, Phase 2 Trial.” The Lancet 389(10064):67–76. |
| 5 | Bauml, Joshua, Tanguy Y. Seiwert, David G. Pfister, Francis Worden, Stephen V. Liu, Jill Gilbert, Nabil F. Saba, Jared Weiss, Lori Wirth, Ammar Sukari, Hyunseok Kang, Michael K. Gibson, Erminia Massarelli, Steven Powell, Amy Meister, Xinxin Shu, Jonathan D. Cheng, and Robert Haddad. 2017. “Pembrolizumab for Platinum- and Cetuximab-Refractory Head and Neck Cancer: Results from a Single-Arm, Phase II Study.” Journal of Clinical Oncology 35(14):1542–49. |
| 6 | Bellmunt, J., R. De Wit, D. J. Vaughn, Y. Fradet, J. L. Lee, L. Fong, N. J. Vogelzang, M. A. Climent, D. P. Petrylak, T. K. Choueiri, A. Necchi, W. Gerritsen, H. Gurney, D. I. Quinn, S. Culine, C. N. Sternberg, Y. Mai, C. H. Poehlein, R. F. Perini, and D. F. Bajorin. 2017. “Pembrolizumab as Second-Line Therapy for Advanced Urothelial Carcinoma.” New England Journal of Medicine 376(11):1015–26. |
| 7 | Bonta, Ioana, John Florin Isac, Eyal Meiri, Dacian Bonta, and Patricia Rich. 2017. “Correlation between Tumor Mutation Burden and Response to Immunotherapy.” Journal of Clinical Oncology 35(15_suppl):e14579–e14579. |
| 8 | Borghaei, H., L. Paz-Ares, L. Horn, D. R. Spigel, M. Steins, N. E. Ready, L. Q. Chow, E. E. Vokes, E. Felip, E. Holgado, F. Barlesi, M. Kohlhufl, O. Arrieta, M. A. Burgio, J. Fayette, H. Lena, E. Poddubskaya, D. E. Gerber, S. N. Gettinger, C. M. Rudin, N. Rizvi, L. Crina, G. R. Blumenschein, S. J. Antonia, C. Dorange, C. T. Harbison, F. Graf Finckenstein, and J. R. Brahmer. 2015. “Nivolumab versus Docetaxel in Advanced Nonsquamous Non-Small-Cell Lung Cancer.” New England Journal of Medicine 373(17):1627–39. |
| 9 | Campesato, Luís Felipe, Romualdo Barroso-Sousa, Leandro Jimenez, Bruna R. Correa, Jorge Sabbaga, Paulo M. Hoff, Luiz F. L. Reis, Pedro Alexandre F. Galante, and Anamaria A. Camargo. 2015. “Comprehensive Cancer-Gene Panels Can Be Used to Estimate Mutational Load and Predict Clinical Benefit to PD-1 Blockade in Clinical Practice.” Oncotarget 6(33):34221–27. |
| 10 | Carbone, D. P., M. Reck, L. Paz-Ares, B. Creelan, L. Horn, M. Steins, E. Felip, M. M. Van Den Heuvel, T. E. Ciuleanu, F. Badin, N. Ready, T. J. N. Hiltermann, S. Nair, R. Juergens, S. Peters, E. Minenza, J. M. Wrangle, D. Rodriguez-Abreu, H. Borghaei, G. R. Blumenschein, L. C. Villaruz, L. Havel, J. Krejci, J. Corral Jaime, H. Chang, W. J. Geese, P. Bhagavatheeswaran, A. C. Chen, and M. A. Socinski. 2017. “First-Line Nivolumab in Stage IV or Recurrent Non-Small-Cell Lung Cancer.” New England Journal of Medicine 376(25):2415–26. |
| 11 | Chow, Laura Q. M., Robert Haddad, Shilpa Gupta, Amit Mahipal, Ranee Mehra, Makoto Tahara, Raanan Berger, Joseph Paul Eder, Barbara Burtness, Se Hoon Lee, Bhumsuk Keam, Hyunseok Kang, Kei Muro, Jared Weiss, Ravit Geva, Chia Chi Lin, Hyun Cheol |

| | |
|---|---|
| | Chung, Amy Meister, Marisa Dolled-Filhart, Kumudu Pathiraja, Jonathan D. Cheng, and Tanguy Y. Seiwert. 2016. “Antitumor Activity of Pembrolizumab in Biomarker-Unselected Patients with Recurrent and/or Metastatic Head and Neck Squamous Cell Carcinoma: Results from the Phase Ib KEYNOTE-012 Expansion Cohort.” Journal of Clinical Oncology 34(32):3838–45. |
| 12 | Chung, Hyun Cheol, Jose A. Lopez-Martin, Stephen Chuan-Hao Kao, Wilson H. Miller, Willeke Ros, Bo Gao, Aurelien Marabelle, Maya Gottfried, Alona Zer, Jean-Pierre Delord, Nicolas Penel, Shadia Ibrahim Jalal, Lei Xu, Susan Zeigenfuss, Scott K. Pruitt, and Sarina Anne Piha-Paul. 2018. “Phase 2 Study of Pembrolizumab in Advanced Small-Cell Lung Cancer (SCLC): KEYNOTE-158.” Journal of Clinical Oncology 36(15_suppl):8506–8506. |
| 13 | Chung, Hyun Cheol, Willeke Ros, Jean Pierre Delord, Ruth Perets, Antoine Italiano, Ronnie Shapira-Frommer, Lyudmila Manzuk, Sarina A. Piha-Paul, Lei Xu, Susan Zeigenfuss, Scott K. Pruitt, and Alexandra Leary. 2019. “Efficacy and Safety of Pembrolizumab in Previously Treated Advanced Cervical Cancer: Results from the Phase II KEYNOTE-158 Study.” Journal of Clinical Oncology 37(17):1470–78. |
| 14 | Cohen, Ezra E. W., Denis Soulières, Christophe Le Tourneau, José Dinis, Lisa Licitra, Myung Ju Ahn, Ainara Soria, Jean Pascal Machiels, Nicolas Mach, Ranee Mehra, Barbara Burtness, Pingye Zhang, Jonathan Cheng, Ramona F. Swaby, Kevin J. Harrington, Mirelis Acosta-Rivera, Douglas R. Adkins, Morteza Aghmesheh, Mario Airoldi, Eduardas Aleknavicius, Yousuf Al-Farhat, Alain P. Algazi, Salah Almokadem, Anna Alyasova, Jessica R. Bauman, Marco Benasso, Alfonso Berrocal, Victoria Bray, Barbara Ann Burtness, Francesco Caponigro, Ana Castro, Terrence P. Cescon, Kelvin Chan, Arvind Chaudhry, Bruno Chauffert, Ezra Cohen, Tibor Csoszi, J. P. De Boer, Jean Pierre Delord, Andreas Dietz, Jose Dinis, Charlotte Dupuis, Laurence Digue, Jozsef Erfan, Yolanda Escobar Alvarez, Mererid Evans, Mary Jo Fidler, Martin David Forster, Signe Friesland, Apar K. Ganti, Lionnel Geoffrois, Cliona Grant, Viktor Gruenwald, Kevin Harrington, Thomas Hoffmann, Geza Horvai, Arturas Inciura, Raymond Jang, Petra Jankowska, Antonio Jimeno, Mano Joseph, Alejandro Juarez Ramiro, Boguslawa Karaszewska, Andrzej Kawecki, Ulrich Keilholz, Ulrich Keller, Sung Bae Kim, Judit Kocsis, Nuria Kotecki, Mark F. Kozloff, Julio Lambea, Laszlo Landherr, Yuri Lantsukhay, Sergey Alexandrovich Lazarev, Lip Way Lee, Igor Dmitrievich Lifirenko, Danko Martincic, Oleg Vladmirovhich Matorin, Margaret McGrath, Krzysztof Misiukiewicz, John C. Morris, Fagim Fanisovich Mufazalov, Jiaxin Niu, Devraj Pamoorthy Srinivasan, Pedro Perez Segura, Daniel Rauch, Maria Leonor Ribeiro, Cristina Rodriguez, Frederic Rolland, Antonio Russo, Agnes Ruzsa, Frederico Sanches, Sang Won Shin, Mikhail Shtiveland, Denis Soulieres, Pol Specenier, Eva Szekanecz, Judit Szota, Carla M. L. van Herpen, Hector A. Velez-Cortes, William V. Walsh, Stefan Wilop, Ralph Winterhalder, Marek Wojtukiewicz, Deborah Wong, and Dan Zandberg. 2019. “Pembrolizumab versus Methotrexate, Docetaxel, or Cetuximab for Recurrent or Metastatic Head-and-Neck Squamous Cell Carcinoma (KEYNOTE-040): A Randomised, Open-Label, Phase 3 Study.” The Lancet 393(10167):156–67. |
| 15 | Cristescu, Razvan, Robin Mogg, Mark Ayers, Andrew Albright, Erin Murphy, Jennifer Yearley, Xinwei Sher, Xiao Qiao Liu, Hongchao Lu, Michael Nebozhyn, Chunsheng Zhang, Jared K. Lunceford, Andrew Joe, Jonathan Cheng, Andrea L. Webber, Nageatte Ibrahim, Elizabeth R. Plimack, Patrick A. Ott, Tanguy Y. Seiwert, Antoni Ribas, Terrill K. McClanahan, Joanne E. Tomassini, Andrey Loboda, and David Kaufman. 2018. “Pan-Tumor Genomic Biomarkers for PD-1 Checkpoint Blockade-Based Immunotherapy.” Science 362(6411). |
| 16 | Daud, Adil I., Omid Hamid, Antoni Ribas, F. Stephen Hodi, Wen-Jen Hwu, Richard Kefford, Jedd Wolchok, Peter Hersey, Jeffrey S. Weber, Richard Joseph, Tara C. Gangadhar, Roxana S. Dronca, Amita Patnaik, Hassane Zarour, Anthony M. Joshua, Kevin Gergich, Dianna Wu, Jared K. Lunceford, Kenneth Emancipator, Marisa Dolled-Filhart, Nicole Li, Scot Ebbinghaus, S. Peter Kang, and Caroline |

| | |
|---|---|
| | Robert. 2014. “Abstract CT104: Antitumor Activity of the Anti-PD-1 Monoclonal Antibody MK-3475 in Melanoma(MEL): Correlation of Tumor PD-L1 Expression with Outcome.” Pp. CT104–CT104 in Cancer Research. Vol. 74. American Association for Cancer Research (AACR). |
| 17 | Daud, Adil I., Jedd D. Wolchok, Caroline Robert, Wen Jen Hwu, Jeffrey S. Weber, Antoni Ribas, F. Stephen Hodi, Anthony M. Joshua, Richard Kefford, Peter Hersey, Richard Joseph, Tara C. Gangadhar, Roxana Dronca, Amita Patnaik, Hassane Zarour, Charlotte Roach, Grant Toland, Jared K. Lunceford, Xiaoyun Nicole Li, Kenneth Emancipator, Marisa Dolled-Filhart, S. Peter Kang, Scot Ebbinghaus, and Omid Hamid. 2016. “Programmed Death-Ligand 1 Expression and Response to the Anti-Programmed Death 1 Antibody Pembrolizumab in Melanoma.” Journal of Clinical Oncology 34(34):4102–9. |
| 18 | Dirix, Luc Y., Istvan Takacs, Guy Jerusalem, Petros Nikolinakos, Hendrik Tobias Arkenau, Andres Forero-Torres, Ralph Boccia, Marc E. Lippman, Robert Somer, Martin Smakal, Leisha A. Emens, Borys Hrinczenko, William Edenfield, Jayne Gurtler, Anja von Heydebreck, Hans Juergen Grote, Kevin Chin, and Erika P. Hamilton. 2018. “Avelumab, an Anti-PD-L1 Antibody, in Patients with Locally Advanced or Metastatic Breast Cancer: A Phase 1b JAVELIN Solid Tumor Study.” Breast Cancer Research and Treatment 167(3):671–86. |
| 19 | El-Khoueiry, Anthony B., Bruno Sangro, Thomas Yau, Todd S. Crocenzi, Masatoshi Kudo, Chiun Hsu, Tae You Kim, Su Pin Choo, Jörg Trojan, Theodore H. Welling, Tim Meyer, Yoon Koo Kang, Winnie Yeo, Akhil Chopra, Jeffrey Anderson, Christine dela Cruz, Lixin Lang, Jaclyn Neely, Hao Tang, Homa B. Dastani, and Ignacio Melero. 2017. “Nivolumab in Patients with Advanced Hepatocellular Carcinoma (CheckMate 040): An Open-Label, Non-Comparative, Phase 1/2 Dose Escalation and Expansion Trial.” The Lancet 389(10088):2492–2502. |
| 20 | Ellen Maher, V., Laura L. Fernandes, Chana Weinstock, Shenghui Tang, Sundeep Agarwal, Michael Brave, Yang Min Ning, Harpreet Singh, Daniel Suzman, James Xu, Kirsten B. Goldberg, Rajeshwari Sridhara, Amna Ibrahim, Marc Theoret, Julia A. Beaver, and Richard Pazdur. 2019. “Analysis of the Association between Adverse Events and Outcome in Patients Receiving a Programmed Death Protein 1 or Programmed Death Ligand 1 Antibody.” Journal of Clinical Oncology 37(30):2730–37. |
| 21 | Eng, Cathy, Tae Won Kim, Johanna Bendell, Guillem Argilés, Niall C. Tebbutt, Maria Di Bartolomeo, Alfredo Falcone, Marwan Fakih, Mark Kozloff, Neil H. Segal, Alberto Sobrero, Yibing Yan, Ilsung Chang, Anne Uyei, Louise Roberts, Fortunato Ciardiello, J. B. Ahn, J. Asselah, S. Badarinath, S. Baijal, S. Begbie, S. Berry, J. L. Canon, R. G. Carbone, A. Cervantes, Y. J. Cha, K. Chang, A. Chaudhry, E. Chmielowska, S. H. Cho, D. Chu, F. Couture, J. Cultrera, D. Cunningham, E. Van Cutsem, P. J. Cuyle, J. Davies, S. Dowden, M. Dvorkin, V. Ganju, R. V. Garcia, R. Kerr, T. Y. Kim, K. King, J. Kortmansky, M. Kozloff, K. O. Lam, J. Lee, A. S. Lee, B. Lesperance, G. Luppi, B. Ma, E. Maiello, R. Mandanas, J. Marshall, G. Marx, S. Mullamitha, M. Nechaeva, J. O. Park, N. Pavlakis, C. G. Ponce, P. Potemski, S. Raouf, J. Reeves, N. Segal, S. Siena, A. Smolin, J. O. Streb, A. Strickland, E. Szutowicz-Zielinska, J. M. Tabernero, B. Tan, J. S. Valera, M. Van den Eynde, P. Vergauwe, M. Vickers, M. Womack, M. Wroblewska, and R. Young. 2019. “Atezolizumab with or without Cobimetinib versus Regorafenib in Previously Treated Metastatic Colorectal Cancer (IMblaze370): A Multicentre, Open-Label, Phase 3, Randomised, Controlled Trial.” The Lancet Oncology 20(6):849–61. |
| 22 | Fehrenbacher, Louis, Alexander Spira, Marcus Ballinger, Marcin Kowanetz, Johan Vansteenkiste, Julien Mazieres, Keunchil Park, David Smith, Angel Artal-Cortes, Conrad Lewanski, Fadi Braiteh, Daniel Waterkamp, Pei He, Wei Zou, Daniel S. Chen, Jing Yi, Alan Sandler, and Achim Rittmeyer. 2016. “Atezolizumab versus Docetaxel for Patients with Previously Treated Non-Small-Cell Lung Cancer (POPLAR): A Multicentre, Open-Label, Phase 2 Randomised Controlled Trial.” The Lancet 387(10030):1837–46. |

| | |
|---|---|
| 23 | Ferris, R. L., Geroge Blumenschein, J. Fayette, J. Guigay, A. D. Colevas, L. Licitra, K. Harrington, S. Kasper, E. E. Vokes, C. Even, F. Worden, N. F. Saba, L. C. Iglesia. Docampo, R. Haddad, T. Rordorf, N. Kiyota, M. Tahara, M. Monga, M. Lynch, W. J. Geese, J. Kopit, J. W. Shaw, and M. L. Gillison. 2016. "Nivolumab for Recurrent Squamous-Cell Carcinoma of the Head and Neck." New England Journal of Medicine 375(19):1856–67. |
| 24 | Fuchs, Charles S., Toshihiko Doi, Raymond W. Jang, Kei Muro, Taroh Satoh, Manuela Machado, Weijing Sun, Shadia I. Jalal, Manish A. Shah, Jean Phillipe Metges, Marcelo Garrido, Talia Golan, Mario Mandala, Zev A. Wainberg, Daniel V. Catenacci, Atsushi Ohtsu, Kohei Shitara, Ravit Geva, Jonathan Bleeker, Andrew H. Ko, Geoffrey Ku, Philip Philip, Peter C. Enzinger, Yung Jue Bang, Diane Levitan, Jiangdian Wang, Minori Rosales, Rita P. Dalal, and Harry H. Yoon. 2018. "Safety and Efficacy of Pembrolizumab Monotherapy in Patients with Previously Treated Advanced Gastric and Gastroesophageal Junction Cancer: Phase 2 Clinical KEYNOTE-059 Trial." JAMA Oncology 4(5). |
| 25 | Gandara, David R., Sarah M. Paul, Marcin Kowanetz, Erica Schleifman, Wei Zou, Yan Li, Achim Rittmeyer, Louis Fehrenbacher, Geoff Otto, Christine Malboeuf, Daniel S. Lieber, Doron Lipson, Jacob Silterra, Lukas Amler, Todd Riehl, Craig A. Cummings, Priti S. Hegde, Alan Sandler, Marcus Ballinger, David Fabrizio, Tony Mok, and David S. Shames. 2018. "Blood-Based Tumor Mutational Burden as a Predictor of Clinical Benefit in Non-Small-Cell Lung Cancer Patients Treated with Atezolizumab." Nature Medicine 24(9):1441–48. |
| 26 | Gandhi, L., D. Rodríguez-Abreu, S. Gadgeel, E. Esteban, E. Felip, F. De Angelis, M. Domine, P. Clingan, M. J. Hochmair, S. F. Powell, S. Y. S. Cheng, H. G. Bischoff, N. Peled, F. Grossi, R. R. Jennens, M. Reck, R. Hui, E. B. Garon, M. Boyer, B. Rubio-Viqueira, S. Novello, T. Kurata, J. E. Gray, J. Vida, Z. Wei, J. Yang, H. Raftopoulos, M. C. Pietanza, and M. C. Garassino. 2018. "Pembrolizumab plus Chemotherapy in Metastatic Non-Small-Cell Lung Cancer." New England Journal of Medicine 378(22):2078–92. |
| 27 | Gandhi, Leena, Ani Balmanoukian, Rina Hui, Omid Hamid, Naiyer A. Rizvi, Natasha Leighl, Matthew Gubens, Jonathan W. Goldman, Gregory M. Lubiniecki, Kenneth Emancipator, Marisa Dolled-Filhart, Jared K. Lunceford, Michelle Niewood, Kevin Gergich, and Edward B. Garon. 2014. "Abstract CT105: MK-3475 (Anti-PD-1 Monoclonal Antibody) for Non-Small Cell Lung Cancer (NSCLC): Antitumor Activity and Association with Tumor PD-L1 Expression." Pp. CT105–CT105 in Cancer Research. Vol. 74. American Association for Cancer Research (AACR). |
| 28 | Garon, Edward B., Naiyer A. Rizvi, Rina Hui, Natasha Leighl, Ani S. Balmanoukian, Joseph Paul Eder, Amita Patnaik, Charu Aggarwal, Matthew Gubens, Leora Horn, Enric Carcereny, Myung-Ju Ahn, Enriqueta Felip, Jong-Seok Lee, Matthew D. Hellmann, Omid Hamid, Jonathan W. Goldman, Jean-Charles Soria, Marisa Dolled-Filhart, Ruth Z. Rutledge, Jin Zhang, Jared K. Lunceford, Reshma Rangwala, Gregory M. Lubiniecki, Charlotte Roach, Kenneth Emancipator, and Leena Gandhi. 2015. "Pembrolizumab for the Treatment of Non–Small-Cell Lung Cancer." New England Journal of Medicine 372(21):2018–28. |
| 29 | Gettinger, Scott N., Frances A. Shepherd, Scott Joseph Antonia, Julie R. Brahmer, Laura Quan Man Chow, Rosalyn A. Juergens, Hossein Borghaei, Yun Shen, Christopher Harbison, Suresh Alaparthy, Allen C. Chen, and Naiyer A. Rizvi. 2014. "First-Line Nivolumab (Anti-PD-1; BMS-936558, ONO-4538) Monotherapy in Advanced NSCLC: Safety, Efficacy, and Correlation of Outcomes with PD-L1 Status." Journal of Clinical Oncology 32(15_suppl):8024–8024. |
| 30 | Giraldo, Nicolas A., Peter Nguyen, Elizabeth L. Engle, Genevieve J. Kaunitz, Tricia R. Cottrell, Sneha Berry, Benjamin Green, Abha Soni, Jonathan D. Cuda, Julie E. Stein, Joel C. Sunshine, Farah Succaria, Haiying Xu, Aleksandra Ogurtsova, Ludmila Danilova, Candice D. Church, Natalie J. Miller, Steve Fling, Lisa Lundgren, Nirasha Ramchurren, Jennifer H. Yearley, Evan J. Lipson, Mac Cheever, Robert A. Anders, Paul T. Nghiem, Suzanne L. Topalian, and Janis M. Taube. 2018. "Multidimensional, Quantitative Assessment of PD-1/PD-L1 |

| | |
|---|---|
| | Expression in Patients with Merkel Cell Carcinoma and Association with Response to Pembrolizumab.” Journal for ImmunoTherapy of Cancer 6(1):99. |
| 31 | Goodman, Aaron M., Shumei Kato, Lyudmila Bazhenova, Sandip P. Patel, Garrett M. Frampton, Vincent Miller, Philip J. Stephens, Gregory A. Daniels, and Razelle Kurzrock. 2017. “Tumor Mutational Burden as an Independent Predictor of Response to Immunotherapy in Diverse Cancers.” Molecular Cancer Therapeutics 16(11):2598–2608. |
| 32 | Gopalakrishnan, V., C. N. Spencer, L. Nezi, A. Reuben, M. C. Andrews, T. V. Karpinets, P. A. Prieto, D. Vicente, K. Hoffman, S. C. Wei, A. P. Cogdill, L. Zhao, C. W. Hudgens, D. S. Hutchinson, T. Manzo, M. Petaccia De Macedo, T. Cotechini, T. Kumar, W. S. Chen, S. M. Reddy, R. Szczepaniak Sloane, J. Galloway-Pena, H. Jiang, P. L. Chen, E. J. Shpall, K. Rezvani, A. M. Alousi, R. F. Chemaly, S. Shelburne, L. M. Vence, P. C. Okhuysen, V. B. Jensen, A. G. Swennes, F. McAllister, E. Marcelo Riquelme Sanchez, Y. Zhang, E. Le Chatelier, L. Zitvogel, N. Pons, J. L. Austin-Breneman, L. E. Haydu, E. M. Burton, J. M. Gardner, E. Sirmans, J. Hu, A. J. Lazar, T. Tsujikawa, A. Diab, H. Tawbi, I. C. Glitza, W. J. Hwu, S. P. Patel, S. E. Woodman, R. N. Amaria, M. A. Davies, J. E. Gershenwald, P. Hwu, J. E. Lee, J. Zhang, L. M. Coussens, Z. A. Cooper, P. A. Futreal, C. R. Daniel, N. J. Ajami, J. F. Petrosino, M. T. Tetzlaff, P. Sharma, J. P. Allison, R. R. Jenq, and J. A. Wargo. 2018. “Gut Microbiome Modulates Response to Anti-PD-1 Immunotherapy in Melanoma Patients.” Science 359(6371):97–103. |
| 33 | Grosso, Joseph, Christine E. Horak, David Inzunza, Diana M. Cardona, Jason S. Simon, Ashok Kumar Gupta, Vindira Sankar, Jong-Soon Park, Georgia Kollia, Janis M. Taube, Robert Anders, Maria Jure-Kunkel, Jim Novotny, Clive R. Taylor, Xiaoling Zhang, Therese Phillips, Pauline Simmons, and John Cogswell. 2013. “Association of Tumor PD-L1 Expression and Immune Biomarkers with Clinical Activity in Patients (Pts) with Advanced Solid Tumors Treated with Nivolumab (Anti-PD-1; BMS-936558; ONO-4538).” Journal of Clinical Oncology 31(15_suppl):3016–3016. |
| 34 | Hellmann, M. D., L. Paz Ares, R. Bernabe Caro, B. Zurawski, S. W. Kim, E. Carcereny Costa, K. Park, A. Alexandru, L. Lupinacci, E. De La Mora Jimenez, H. Sakai, I. Albert, A. Vergnenegre, S. Peters, K. Syrigos, F. Barlesi, M. Reck, H. Borghaei, J. R. Brahmer, K. J. O fByrn, W. J. Geese, P. Bhagavatheeswaran, S. K. Rabindran, R. S. Kasinathan, F. E. Nathan, and S. S. Ramalingam. 2019. “Nivolumab plus Ipilimumab in Advanced Non-Small-Cell Lung Cancer.” New England Journal of Medicine 381(21):2020–31. |
| 35 | Hellmann, Matthew D., Margaret K. Callahan, Mark M. Awad, Emiliano Calvo, Paolo A. Ascierto, Akin Atmaca, Naiyer A. Rizvi, Fred R. Hirsch, Giovanni Selvaggi, Joseph D. Szustakowski, Ariella Sasson, Ryan Golhar, Patrik Vitazka, Han Chang, William J. Geese, and Scott J. Antonia. 2018. “Tumor Mutational Burden and Efficacy of Nivolumab Monotherapy and in Combination with Ipilimumab in Small-Cell Lung Cancer.” Cancer Cell 33(5):853-861.e4. |
| 36 | Hellmann MD, Nathanson T, Rizvi H, et al. Genomic Features of Response to Combination Immunotherapy in Patients with Advanced Non-Small-Cell Lung Cancer. *Cancer Cell*. 2018;33(5):843-852.e4. doi:10.1016/j.ccell.2018.03.018 |
| 37 | Hellmann, Matthew D., Tudor-Eliade Ciuleanu, Adam Pluzanski, Jong Seok Lee, Gregory A. Otterson, Clarisse Audigier-Valette, Elisa Minenza, Helena Linardou, Sjaak Burgers, Pamela Salman, Hossein Borghaei, Suresh S. Ramalingam, Julie Brahmer, Martin Reck, Kenneth J. O’Byrne, William J. Geese, George Green, Han Chang, Joseph Szustakowski, Prabhu Bhagavatheeswaran, Diane Healey, Yali Fu, Faith Nathan, and Luis Paz-Ares. 2018. “Nivolumab plus Ipilimumab in Lung Cancer with a High Tumor Mutational Burden.” New England Journal of Medicine 378(22):2093–2104. |
| 38 | Herbst, Roy S., Paul Baas, Dong Wan Kim, Enriqueta Felip, José L. Pérez-Gracia, Ji Youn Han, Julian Molina, Joo Hang Kim, Catherine Dubos Arvis, Myung Ju Ahn, Margarita Majem, Mary J. Fidler, Gilberto De Castro, Marcelo Garrido, Gregory M. Lubiniecki, Yue |

| | |
|---|---|
| | Shentu, Ellie Im, Marisa Dolled-Filhart, and Edward B. Garon. 2016. "Pembrolizumab versus Docetaxel for Previously Treated, PD-L1-Positive, Advanced Non-Small-Cell Lung Cancer (KEYNOTE-010): A Randomised Controlled Trial." The Lancet 387(10027):1540–50. |
| 39 | Herbst, Roy S., Jean Charles Soria, Marcin Kowanetz, Gregg D. Fine, Omid Hamid, Michael S. Gordon, Jeffery A. Sosman, David F. McDermott, John D. Powderly, Scott N. Gettinger, Holbrook E. K. Kohrt, Leora Horn, Donald P. Lawrence, Sandra Rost, Maya Leabman, Yuanyuan Xiao, Ahmad Mokatrin, Hartmut Koeppen, Priti S. Hegde, Ira Mellman, Daniel S. Chen, and F. Stephen Hodi. 2014. "Predictive Correlates of Response to the Anti-PD-L1 Antibody MPDL3280A in Cancer Patients." Nature 515(7528):563–67. |
| 40 | Horn, Leora, David R. Spigel, Everett E. Vokes, Esther Holgado, Neal Ready, Martin Steins, Elena Poddubskaya, Hossein Borghaei, Enriqueta Felip, Luis Paz-Ares, Adam Pluzanski, Karen L. Reckamp, Marco A. Burgio, Martin Kohlhäeufl, David Waterhouse, Fabrice Barlesi, Scott Antonia, Oscar Arrieta, Jérôme Fayette, Lucio Crinò, Naiyer Rizvi, Martin Reck, Matthew D. Hellmann, William J. Geese, Ang Li, Anne Blackwood-Chirchir, Diane Healey, Julie Brahmer, and Wilfried E. E. Eberhardt. 2017. "Nivolumab versus Docetaxel in Previously Treated Patients with Advanced Non-Small-Cell Lung Cancer: Two-Year Outcomes from Two Randomized, Open-Label, Phase III Trials (CheckMate 017 and CheckMate 057)." Journal of Clinical Oncology 35(35):3924–33. |
| 41 | Hugo, Willy, Jesse M. Zaretsky, Lu Sun, Chunying Song, Blanca Homet Moreno, Siwen Hu-Lieskovan, Beata Berent-Maoz, Jia Pang, Bartosz Chmielowski, Grace Cherry, Elizabeth Seja, Shirley Lomeli, Xiangju Kong, Mark C. Kelley, Jeffrey A. Sosman, Douglas B. Johnson, Antoni Ribas, and Roger S. Lo. 2016. "Genomic and Transcriptomic Features of Response to Anti-PD-1 Therapy in Metastatic Melanoma." Cell 165(1):35–44. |
| 42 | Janjigian, Yelena Y., Johanna Bendell, Emiliano Calvo, Joseph W. Kim, Paolo A. Ascierto, Padmanee Sharma, Patrick A. Ott, Katriina Peltola, Dirk Jaeger, Jeffry Evans, Filippo De Braud, Ian Chau, Christopher T. Harbison, Cecile Dorange, Marina Tschaika, and Dung T. Le. 2018. "CheckMate-032 Study: Efficacy and Safety of Nivolumab and Nivolumab plus Ipilimumab in Patients with Metastatic Esophagogastric Cancer." Pp. 2836–44 in Journal of Clinical Oncology. Vol. 36. American Society of Clinical Oncology. |
| 43 | Johnson, Douglas B., Jennifer Bordeaux, Ju Young Kim, Christine Vaupel, David L. Rimm, Thai H. Ho, Richard W. Joseph, Adil I. Daud, Robert M. Conry, Elizabeth M. Gaughan, Leonel F. Hernandez-Aya, Anastasios Dimou, Pauline Funchain, James Smithy, John S. Witte, Svetlana B. McKee, Jennifer Ko, John M. Wrangle, Bashar Dabbas, Shabnam Tangri, Jelveh Lameh, Jeffrey Hall, Joseph Markowitz, Justin M. Balko, and Naveen Dakappagari. 2018. "Quantitative Spatial Profiling of PD-1/PD-L1 Interaction and HLA-DR/IDO-1 Predicts Improved Outcomes of Anti-PD-1 Therapies in Metastatic Melanoma." Clinical Cancer Research 24(21):5250–60. |
| 44 | Johnson, Douglas B., Garrett M. Frampton, Matthew J. Rioth, Erik Yusko, Yaomin Xu, Xingyi Guo, Riley C. Ennis, David Fabrizio, Zachary R. Chalmers, Joel Greenbowe, Siraj M. Ali, Sohail Balasubramanian, James X. Sun, Yuting He, Dennie T. Frederick, Igor Puzanov, Justin M. Balko, Justin M. Cates, Jeffrey S. Ross, Catherine Sanders, Harlan Robins, Yu Shyr, Vincent A. Miller, Philip J. Stephens, Ryan J. Sullivan, Jeffrey A. Sosman, and Christine M. Lovly. 2016. "Targeted next Generation Sequencing Identifies Markers of Response to PD-1 Blockade." Cancer Immunology Research 4(11):959–67. |
| 45 | Kaufman, Howard L., Jeffery S. Russell, Omid Hamid, Shailender Bhatia, Patrick Terheyden, Sandra P. D'Angelo, Kent C. Shih, Céleste Lebbé, Michele Milella, Isaac Brownell, Karl D. Lewis, Jochen H. Lorch, Anja von Heydebreck, Meliessa Hennessy, and Paul Nghiem. 2018. "Updated Efficacy of Avelumab in Patients with Previously Treated Metastatic Merkel Cell Carcinoma after ≥1 Year of Follow-up: JAVELIN Merkel 200, a Phase 2 Clinical Trial." Journal for ImmunoTherapy of Cancer 6(1). |
| 46 | Kaufman, Howard L., Jeffery Russell, Omid Hamid, Shailender Bhatia, Patrick Terheyden, Sandra P. D'Angelo, Kent C. Shih, Céleste Lebbé, Gerald P. Linette, Michele Milella, Isaac Brownell, Karl D. Lewis, Jochen H. Lorch, Kevin Chin, Lisa Mahnke, Anja von |

| | |
|---|---|
| | Heydebreck, Jean Marie Cuillerot, and Paul Nghiem. 2016. "Avelumab in Patients with Chemotherapy-Refractory Metastatic Merkel Cell Carcinoma: A Multicentre, Single-Group, Open-Label, Phase 2 Trial." The Lancet Oncology 17(10):1374–85. |
| 47 | Kefford, Richard, Antoni Ribas, Omid Hamid, Caroline Robert, Adil Daud, Jedd D. Wolchok, Anthony M. Joshua, F. Stephen Hodi, Tara C. Gangadhar, Peter Hersey, Jeffrey S. Weber, Roxana Stefania Dronca, Amita Patnaik, Hassane M. Zarour, Marisa Dolled-Filhart, Jared Lunceford, Kenneth Emancipator, Scot Ebbinghaus, Soonmo Peter Kang, and Wen-Jen Hwu. 2014. "Clinical Efficacy and Correlation with Tumor PD-L1 Expression in Patients (Pts) with Melanoma (MEL) Treated with the Anti-PD-1 Monoclonal Antibody MK-3475." Journal of Clinical Oncology 32(15_suppl):3005–3005. |
| 48 | Khagi, Yulian, Aaron M. Goodman, Gregory A. Daniels, Sandip P. Patel, Assuntina G. Sacco, James M. Randall, Lyudmila A. Bazhenova, and Razelle Kurzrock. 2017. "Hypermutated Circulating Tumor DNA: Correlation with Response to Checkpoint Inhibitor–Based Immunotherapy." Clinical Cancer Research 23(19):5729–36. |
| 49 | Langer CJ, Gadgeel SM, Borghaei H, et al. Carboplatin and pemetrexed with or without pembrolizumab for advanced, non-squamous non-small-cell lung cancer: a randomised, phase 2 cohort of the open-label KEYNOTE-021 study. *Lancet Oncol*. 2016;17(11):1497-1508. doi:10.1016/S1470-2045(16)30498-3 |
| 50 | Larkin, J., V. Chiarion-Sileni, R. Gonzalez, J. J. Grob, C. L. Cowey, C. D. Lao, D. Schadendorf, R. Dummer, M. Smylie, P. Rutkowski, P. F. Ferrucci, A. Hill, J. Wagstaff, M. S. Carlino, J. B. Haanen, M. Maio, I. Marquez-Rodas, G. A. McArthur, P. A. Ascierto, G. V. Long, M. K. Callahan, M. A. Postow, K. Grossmann, M. Sznol, B. Dreno, L. Bastholt, A. Yang, L. M. Rollin, C. Horak, F. S. Hodi, and J. D. Wolchok. 2015. "Combined Nivolumab and Ipilimumab or Monotherapy in Untreated Melanoma." New England Journal of Medicine 373(1):23–34. |
| 51 | Larkin, James, David Minor, Sandra D'Angelo, Bart Neyns, Michael Smylie, Wilson H. Miller, Ralf Gutzmer, Gerald Linette, Bartosz Chmielowski, Christopher D. Lao, Paul Lorigan, Kenneth Grossmann, Jessica C. Hassel, Mario Sznol, Adil Daud, Jeffrey Sosman, Nikhil Khushalani, Dirk Schadendorf, Christoph Hoeller, Dana Walker, George Kong, Christine Horak, and Jeffrey Weber. 2018. "Overall Survival in Patients with Advanced Melanoma Who Received Nivolumab versus Investigator's Choice Chemotherapy in CheckMate 037: A Randomized, Controlled, Open-Label Phase III Trial." Pp. 383–90 in Journal of Clinical Oncology. Vol. 36. American Society of Clinical Oncology. |
| 52 | Loo, Kimberly, Katy K. Tsai, Kelly Mahuron, Jacqueline Liu, Mariela L. Pauli, Priscila M. Sandoval, Adi Nosrati, James Lee, Lawrence Chen, Jimmy Hwang, Lauren S. Levine, Matthew F. Krummel, Alain P. Algazi, Miguel Pampaloni, Michael D. Alvarado, Michael D. Rosenblum, and Adil I. Daud. 2017. "Partially Exhausted Tumor-Infiltrating Lymphocytes Predict Response to Combination Immunotherapy." JCI Insight 2(14). |
| 53 | Matson, Vyara, Jessica Fessler, Riyue Bao, Tara Chongsuwat, Yuanyuan Zha, Maria Luisa Alegre, Jason J. Luke, and Thomas F. Gajewski. 2018. "The Commensal Microbiome Is Associated with Anti-PD-1 Efficacy in Metastatic Melanoma Patients." Science 359(6371):104–8. |
| 54 | McDermott, David F., Mahrukh A. Huseni, Michael B. Atkins, Robert J. Motzer, Brian I. Rini, Bernard Escudier, Lawrence Fong, Richard W. Joseph, Sumanta K. Pal, James A. Reeves, Mario Sznol, John Hainsworth, W. Kimryn Rathmell, Walter M. Stadler, Thomas Hutson, Martin E. Gore, Alain Ravaud, Sergio Bracarda, Cristina Suárez, Riccardo Danielli, Viktor Gruenwald, Toni K. Choueiri, Dorothee Nickles, Suchit Jhunjhunwala, Elisabeth Piault-Louis, Alpa Thobhani, Jiaheng Qiu, Daniel S. Chen, Priti S. Hegde, Christina |

| | |
|---|---|
| | Schiff, Gregg D. Fine, and Thomas Powles. 2018. “Clinical Activity and Molecular Correlates of Response to Atezolizumab Alone or in Combination with Bevacizumab versus Sunitinib in Renal Cell Carcinoma.” Nature Medicine 24(6):749–57. |
| 55 | Mehra, Ranee, Tanguy Y. Seiwert, Shilpa Gupta, Jared Weiss, Iris Gluck, Joseph P. Eder, Barbara Burtness, Makoto Tahara, Bhumsuk Keam, Hyunseok Kang, Kei Muro, Ravit Geva, Hyun Cheol Chung, Chia Chi Lin, Deepti Aurora-Garg, Archana Ray, Kumudu Pathiraja, Jonathan Cheng, Laura Q. M. Chow, and Robert Haddad. 2018. “Efficacy and Safety of Pembrolizumab in Recurrent/Metastatic Head and Neck Squamous Cell Carcinoma: Pooled Analyses after Long-Term Follow-up in KEYNOTE-012.” British Journal of Cancer 119(2):153–59. |
| 56 | Mok, Tony S. K., Yi Long Wu, Iveta Kudaba, Dariusz M. Kowalski, Byoung Chul Cho, Hande Z. Turna, Gilberto Castro, Vichien Srimuninnimit, Konstantin K. Laktionov, Igor Bondarenko, Kaoru Kubota, Gregory M. Lubiniecki, Jin Zhang, Debra Kush, Gilberto Lopes, Gonzalo Gomez Aubin, Luis Fein, Diego Kaen, Ruben Kowalyszyn, Guillermo Lerzo, Gaston Martinengo, Matias Molina, Eduardo Richardet, Pablo Picon, Mirta Varela, Juan Jose Zarba, Sergio Jobim de Azevedo, Carlos Henrique Barrios, Carlos Beato, Carlos Alexandre Sydow Cerny, Pedro Rafael Martins De Marchi, Gustavo Fernandes, Fabio Andre Franke, Helano Freitas, Gustavo Girotto, Valeria Lopes, Lucas Santos, Marcos Andre Costa, Andrea Kazumi Shimada, Oren Smaletz, Joao Paulo Holanda Soares, Ana Paula Victorino, Carlos Ferreira, Marchela Koleva, Krassimir Koynov, Rumyana Micheva, Tsvetan Deliverski, Zhasmina Milanova, Boyan Doganov, Susanna Cheng, Flavia De Angelis, Giovanna Speranza, Rosalyn Anne Juergens, Doran Ksienski, David Fenton, Osvaldo Aren, Christian Caglevic, Hector Galindo, Felipe Rey, Jianhua Chang, Gongyan Chen, Xi Chen, Xuenong Ouyang, Ying Cheng, Zhenyu Ding, Mei Hou, Yun Fan, Jifeng Feng, Jianxing He, Yong He, Yi Hu, Wei Li, Xiaoqing Liu, Zhe Liu, Shun Lu, Shukui Qin, Qiyou Tang, Buhai Wang, Kai Wang, Li Zhang, Xin Zhang, Jun Zhao, Jie Wang, Caicun Zhou, Jianying Zhou, Qing Zhou, Yilong Wu, Andres Cardona, Ricardo Duarte, Luis Gomez Wolff, Angela Zambrano, Marcela Vallejo, Libor Havel, Vitezslav Kolek, Petr Kolman, Leona Koubkova, Lubos Petruzelka, Patrice Popelkova, Jaromir Roubec, Jaroslav Vanasek, Tomas Vlasek, Jana Jaal, Gerli Kuusk, Oscar Avendano, Hugo Castro, Karla Lopez, Mario Sandoval, Chung Man James Ho, Sing Hung Lo, Ibolya Laczo, Bela Piko, Gyula Ostoros, Keisuke Aoe, Yasuhito Fujisaka, Tomonori Hirashima, Atsushi Horiike, Yukio Hosomi, Katsuyuki Hotta, Masao Ichiki, Fumio Imamura, Yasuo Iwamoto, Kazuo Kasahara, Nobuyuki Katakami, Terufumi Kato, Shuji Murakami, Tomoya Kawaguchi, Kazuma Kishi, Kaoru Kubota, Takayasu Kurata, Yoshitaro Torii, Yasuharu Nakahara, Takashi Nishimura, Tatsuo Ohira, Hideo Saka, Toshiyuki Sawa, Nobuhiko Seki, Shunichi Sugawara, Kazuhisa Takahashi, Nagio Takigawa, Hiroshi Tanaka, Kazuhiko Yamada, Takuma Yokoyama, Toshihide Yokoyama, Hiroshige Yoshioka, Iveta Kudaba, Gunta Purkalne, Zinaida Stara, Alvydas Cesas, Saulius Cicenas, Marius Zemaitis, Soon Hin How, Chong Kin Liam, Choo Khoon Ong, Lye Mun Tho, Oscar Arrieta Rodriguez, Flor de The Bustamante Valles, Carlos Hernandez Hernandez, Luis Mas, Luis Vera, Jorge Salas, Hermes Tejada, Regina Edusma-Dy, Christina Galvez, Guia Elena Imelda Ladrera, Jerry Tan Chun Bing, Jacek Jassem, Ewa Kalinka-Warzocha, Boguslawa Karaszewska, Andrzej Kazarnowicz, Dariusz Kowalski, Krzysztof Lesniewski Kmak, Rodryg Ramlau, Antonio Araujo, Fernando Barata, Nuno Gil, Venceslau Hespanhol, Aurelia Alexandru, Mircea Dediu, Nelly Cherciu, Daniel Ciurescu, Doina Ganea, Lucian Miron, Daniela Sirbu, Maria Turdean, Sergey Emelyanov, Nina Karaseva, Lyudmila Kuzina, Sergey Lazarev, Igor Lifirenko, Larisa Bolotina, Oleg Lipatov, Elena Ovchinnikova, Marina Matrosova, Anna Alyasova, Artem Poltoratsky, Pavel Taranov, Oleg Zarubenkov, Graham Cohen, Lydia Dreosti, Freddy Seolwane, Jacqueline Hall, Gregory Hart, Christa Jordaan, Sayeuri Buddu, Michiel Botha, Gregory Landers, Bernardo Rappaport, Paul Ruff, Lucinda Shepherd, Waldemar Szpak, Myung Ju Ahn, Byoung Chul Cho, Joo Hang Kim, Per Bergstrom, Ronny Ohman, Hakan Griph, Daniel Betticher, Adrian Ochsenbein, Alfred Zippelius, Gee Chen Chan, Chao Hua Chiu, Te Chun Hsia, Wu Chou Su, Chih Hsin |

| | |
|---|---|
| | Yang, Touch Ativitavas, Pongwut Danchaivijitr, Vichien Srimuninnimit, Kasan Seetalarom, Aumkhae Sookprasert, Virote Sriuranpong, Ozden Altundag, Filiz Cay Senler, Mustafa Erman, Tuncay Goksel, Erdem Goker, Ozgur Ozyilkan, Mesut Seker, Mahmut Gumus, Hande Turna, Fulden Yumuk, Grigory Adamchuk, Igor Bondarenko, Oleksandr Ivashchuk, Olga Ponomarova, Andrii Rusyn, Sergii Shevnya, Yaroslav Shparyk, Ivan Sinielnikov, Orest Andrusenko, Dmytro Trukhyn, Grygoriy Ursol, Ihor Vynnychenko, Tien Quang Nguyen, Xuan Dung Pham, Gilberto Castro, and Kaoru Kubota. 2019. "Pembrolizumab versus Chemotherapy for Previously Untreated, PD-L1-Expressing, Locally Advanced or Metastatic Non-Small-Cell Lung Cancer (KEYNOTE-042): A Randomised, Open-Label, Controlled, Phase 3 Trial." The Lancet 393(10183):1819–30. |
| 57 | Motzer, R. J., N. M. Tannir, D. F. McDermott, O. Arén Frontera, B. Melichar, T. K. Choueiri, E. R. Plimack, P. Barthélémy, C. Porta, S. George, T. Powles, F. Donskov, V. Neiman, C. K. Kollmannsberger, P. Salman, H. Gurney, R. Hawkins, A. Ravaud, M. O. Grimm, S. Bracarda, C. H. Barrios, Y. Tomita, D. Castellano, B. I. Rini, A. C. Chen, S. Mekan, M. B. McHenry, M. Wind-Rotolo, J. Doan, P. Sharma, H. J. Hammers, and B. Escudier. 2018. "Nivolumab plus Ipilimumab versus Sunitinib in Advanced Renal-Cell Carcinoma." New England Journal of Medicine 378(14):1277–90. |
| 58 | Motzer, Robert J., Konstantin Penkov, John Haanen, Brian Rini, Laurence Albiges, Matthew T. Campbell, Balaji Venugopal, Christian Kollmannsberger, Sylvie Negrier, Motohide Uemura, Jae L. Lee, Aleksandr Vasiliev, Wilson H. Miller, Howard Gurney, Manuela Schmidinger, James Larkin, Michael B. Atkins, Jens Bedke, Boris Alekseev, Jing Wang, Mariangela Mariani, Paul B. Robbins, Aleksander Chudnovsky, Camilla Fowst, Subramanian Hariharan, Bo Huang, Alessandra Di Pietro, and Toni K. Choueiri. 2019. "Avelumab plus Axitinib versus Sunitinib for Advanced Renal-Cell Carcinoma." Pp. 1103–15 in New England Journal of Medicine. Vol. 380. Massachussetts Medical Society. |
| 59 | Muro, Kei, Hyun Cheol Chung, Veena Shankaran, Ravit Geva, Daniel Catenacci, Shilpa Gupta, Joseph Paul Eder, Talia Golan, Dung T. Le, Barbara Burtness, Autumn J. McRee, Chia Chi Lin, Kumudu Pathiraja, Jared Lunceford, Kenneth Emancipator, Jonathan Juco, Minori Koshiji, and Yung Jue Bang. 2016. "Pembrolizumab for Patients with PD-L1-Positive Advanced Gastric Cancer (KEYNOTE-012): A Multicentre, Open-Label, Phase 1b Trial." The Lancet Oncology 17(6):717–26. |
| 60 | Nghiem, Paul, Shailender Bhatia, Evan J. Lipson, William H. Sharfman, Ragini R. Kudchadkar, Andrew S. Brohl, Phillip A. Friedlander, Adil Daud, Harriet M. Kluger, Sunil A. Reddy, Brian C. Boulmay, Adam I. Riker, Melissa A. Burgess, Brent A. Hanks, Thomas Olencki, Kim Margolin, Lisa M. Lundgren, Abha Soni, Nirasha Ramchurren, Candice Church, Song Y. Park, Michi M. Shinohara, Bob Salim, Janis M. Taube, Steven R. Bird, Nageatte Ibrahim, Steven P. Fling, Blanca Homet Moreno, Elad Sharon, Martin A. Cheever, and Suzanne L. Topalian. 2019. "Durable Tumor Regression and Overall Survival in Patients with Advanced Merkel Cell Carcinoma Receiving Pembrolizumab as First-Line Therapy." Journal of Clinical Oncology 37(9):693–702. |
| 61 | Overman, Michael J., Sara Lonardi, Ka Yeung Mark Wong, Heinz Josef Lenz, Fabio Gelsomino, Massimo Aglietta, Michael A. Morse, Eric Van Cutsem, Ray McDermott, Andrew Hill, Michael B. Sawyer, Alain Hendlisz, Bart Neyns, Magali Svrcek, Rebecca A. Moss, Jean Marie Ledeine, Z. Alexander Cao, Shital Kamble, Scott Kopetz, and Thierry André. 2018. "Durable Clinical Benefit with Nivolumab plus Ipilimumab in DNA Mismatch Repair-Deficient/Microsatellite Instability-High Metastatic Colorectal Cancer." Journal of Clinical Oncology 36(8):773–79. |
| 62 | Overman, Michael J., Ray McDermott, Joseph L. Leach, Sara Lonardi, Heinz Josef Lenz, Michael A. Morse, Jayesh Desai, Andrew Hill, Michael Axelson, Rebecca A. Moss, Monica V. Goldberg, Z. Alexander Cao, Jean Marie Ledeine, Gregory A. Maglinte, Scott Kopetz, |

| | |
|---|---|
| | and Thierry André. 2017. "Nivolumab in Patients with Metastatic DNA Mismatch Repair-Deficient or Microsatellite Instability-High Colorectal Cancer (CheckMate 142): An Open-Label, Multicentre, Phase 2 Study." The Lancet Oncology 18(9):1182–91. |
| 63 | Patel, Manish R., John Ellerton, Jeffrey R. Infante, Manish Agrawal, Michael Gordon, Raid Aljumaily, Carolyn D. Britten, Luc Dirix, Keun Wook Lee, Mathew Taylor, Patrick Schöffski, Ding Wang, Alain Ravaud, Arnold B. Gelb, Junyuan Xiong, Galit Rosen, James L. Gulley, and Andrea B. Apolo. 2018. "Avelumab in Metastatic Urothelial Carcinoma after Platinum Failure (JAVELIN Solid Tumor): Pooled Results from Two Expansion Cohorts of an Open-Label, Phase 1 Trial." The Lancet Oncology 19(1):51–64. |
| 64 | Paz-Ares, L., A. Luft, D. Vicente, A. Tafreshi, M. Gümüş, J. Mazières, B. Hermes, F. Çay Şenler, T. Csőszi, A. Fülöp, J. Rodríguez-Cid, J. Wilson, S. Sugawara, T. Kato, K. H. Lee, Y. Cheng, S. Novello, B. Halmos, X. Li, G. M. Lubiniecki, B. Piperdi, and D. M. Kowalski. 2018. "Pembrolizumab plus Chemotherapy for Squamous Non-Small-Cell Lung Cancer." New England Journal of Medicine 379(21):2040–51. |
| 65 | Plimack, Elizabeth R., Joaquim Bellmunt, Shilpa Gupta, Raanan Berger, Laura Q. M. Chow, Jonathan Juco, Jared Lunceford, Sanatan Saraf, Rodolfo F. Perini, and Peter H. O'Donnell. 2017. "Safety and Activity of Pembrolizumab in Patients with Locally Advanced or Metastatic Urothelial Cancer (KEYNOTE-012): A Non-Randomised, Open-Label, Phase 1b Study." The Lancet Oncology 18(2):212–20. |
| 66 | Postow, Michael A., Jason Chesney, Anna C. Pavlick, Caroline Robert, Kenneth Grossmann, David McDermott, Gerald P. Linette, Nicolas Meyer, Jeffrey K. Giguere, Sanjiv S. Agarwala, Montaser Shaheen, Marc S. Ernstoff, David Minor, April K. Salama, Matthew Taylor, Patrick A. Ott, Linda M. Rollin, Christine Horak, Paul Gagnier, Jedd D. Wolchok, and F. Stephen Hodi. 2015. "Nivolumab and Ipilimumab versus Ipilimumab in Untreated Melanoma." New England Journal of Medicine 372(21):2006–17. |
| 67 | Powles, Thomas, Joseph Paul Eder, Gregg D. Fine, Fadi S. Braiteh, Yohann Loriot, Cristina Cruz, Joaquim Bellmunt, Howard A. Burris, Daniel P. Petrylak, Siew Leng Teng, Xiaodong Shen, Zachary Boyd, Priti S. Hegde, Daniel S. Chen, and Nicholas J. Vogelzang. 2014. "MPDL3280A (Anti-PD-L1) Treatment Leads to Clinical Activity in Metastatic Bladder Cancer." Nature 515(7528):558–62. |
| 68 | Powles, Thomas, Peter H. O'Donnell, Christophe Massard, Hendrik Tobias Arkenau, Terence W. Friedlander, Christopher J. Hoimes, Jae Lyun Lee, Michael Ong, Srikala S. Sridhar, Nicholas J. Vogelzang, Mayer N. Fishman, Jingsong Zhang, Sandy Srinivas, Jigar Parikh, Joyce Antal, Xiaoping Jin, Ashok K. Gupta, Yong Ben, and Noah M. Hahn. 2017. "Efficacy and Safety of Durvalumab in Locally Advanced or Metastatic Urothelial Carcinoma: Updated Results from a Phase 1/2 Open-Label Study." JAMA Oncology 3(9). |
| 69 | Puzanov, Igor, Reinhard Dummer, Jacob Schachter, Anna C. Pavlick, Rene Gonzalez, Paolo Antonio Ascierto, Kim Allyson Margolin, Omid Hamid, Sanjiv S. Agarwala, Matteo S. Carlino, Jochen Utikal, Michal Lotem, Antoni Ribas, Peter Mohr, Charlotte M. Roach, Marisa Dolled-Filhart, Xiaoyun Nicole Li, Scot Ebbinghaus, Soonmo Peter Kang, and Adil Daud. 2015. "Efficacy Based on Tumor PD-L1 Expression in KEYNOTE-002, a Randomized Comparison of Pembrolizumab (Pembro; MK-3475) versus Chemotherapy in Patients (Pts) with Ipilimumab-Refractory (IPI-R) Advanced Melanoma (MEL)." Journal of Clinical Oncology 33(15_suppl):3012–3012. |
| 70 | Ready, Neal, Matthew D. Hellmann, Mark M. Awad, Gregory A. Otterson, Martin Gutierrez, Justin F. Gainor, Hossein Borghaei, Jacques Jolivet, Leora Horn, Mihaela Mates, Julie Brahmer, Ian Rabinowitz, Pavan S. Reddy, Jason Chesney, James Orcutt, David R. Spigel, Martin Reck, Kenneth John O'Byrne, Luis Paz-Ares, Wenhua, Phd Hu, Kim Zerba, Xuemei Li, Brian Lestini, William J. Geese, Joseph D. Szustakowski, George Green, Han Chang, and Suresh S. Ramalingam. 2019. "First-Line Nivolumab plus Ipilimumab in Advanced Non-Small-Cell Lung Cancer (CheckMate 568): Outcomes by Programmed Death Ligand 1 and Tumor Mutational Burden as Biomarkers." Pp. 992–1000 in Journal of Clinical Oncology. Vol. 37. American Society of Clinical Oncology. |

| | |
|---|---|
| 71 | Riaz, Nadeem, Jonathan J. Havel, Vladimir Makarov, Alexis Desrichard, Walter J. Urba, Jennifer S. Sims, F. Stephen Hodi, Salvador Martín-Algarra, Rajarsi Mandal, William H. Sharfman, Shailender Bhatia, Wen Jen Hwu, Thomas F. Gajewski, Craig L. Slingluff, Diego Chowell, Sviatoslav M. Kendall, Han Chang, Rachna Shah, Fengshen Kuo, Luc G. T. Morris, John William Sidhom, Jonathan P. Schneck, Christine E. Horak, Nils Weinhold, and Timothy A. Chan. 2017. “Tumor and Microenvironment Evolution during Immunotherapy with Nivolumab.” Cell 171(4):934-949.e15. |
| 72 | Rini, Brian I., Thomas Powles, Michael B. Atkins, Bernard Escudier, David F. McDermott, Cristina Suarez, Sergio Bracarda, Walter M. Stadler, Frede Donskov, Jae Lyun Lee, Robert Hawkins, Alain Ravaud, Boris Alekseev, Michael Staehler, Motohide Uemura, Ugo De Giorgi, Begoña Mellado, Camillo Porta, Bohuslav Melichar, Howard Gurney, Jens Bedke, Toni K. Choueiri, Francis Parnis, Tarik Khaznadar, Alpa Thobhani, Shi Li, Elisabeth Piault-Louis, Gretchen Frantz, Mahrukh Huseni, Christina Schiff, Marjorie C. Green, and Robert J. Motzer. 2019. “Atezolizumab plus Bevacizumab versus Sunitinib in Patients with Previously Untreated Metastatic Renal Cell Carcinoma (IMmotion151): A Multicentre, Open-Label, Phase 3, Randomised Controlled Trial.” The Lancet 393(10189):2404–15. |
| 73 | Rittmeyer, Achim, Fabrice Barlesi, Daniel Waterkamp, Keunchil Park, Fortunato Ciardiello, Joachim von Pawel, Shirish M. Gadgeel, Toyoaki Hida, Dariusz M. Kowalski, Manuel Cobo Dols, Diego L. Cortinovis, Joseph Leach, Jonathan Polikoff, Carlos Barrios, Fairooz Kabbinavar, Osvaldo Arén Frontera, Filippo De Marinis, Hande Turna, Jong Seok Lee, Marcus Ballinger, Marcin Kowanetz, Pei He, Daniel S. Chen, Alan Sandler, and David R. Gandara. 2017. “Atezolizumab versus Docetaxel in Patients with Previously Treated Non-Small-Cell Lung Cancer (OAK): A Phase 3, Open-Label, Multicentre Randomised Controlled Trial.” The Lancet 389(10066):255–65. |
| 74 | Rizvi, Hira, Francisco Sanchez-Vega, Konnor La, Walid Chatila, Philip Jonsson, Darragh Halpenny, Andrew Plodkowski, Niamh Long, Jennifer L. Sauter, Natasha Rekhtman, Travis Hollmann, Kurt A. Schalper, Justin F. Gainor, Ronglai Shen, Ai Ni, Kathryn C. Arbour, Taha Merghoub, Jedd Wolchok, Alexandra Snyder, Jamie E. Chaft, Mark G. Kris, Charles M. Rudin, Nicholas D. Socci, Michael F. Berger, Barry S. Taylor, Ahmet Zehir, David B. Solit, Maria E. Arcila, Marc Ladanyi, Gregory J. Riely, Nikolaus Schultz, and Matthew D. Hellmann. 2018. “Molecular Determinants of Response to Anti-Programmed Cell Death (PD)-1 and Anti-Programmed Death-Ligand 1 (PD-L1) Blockade in Patients with Non-Small-Cell Lung Cancer Profiled with Targeted next-Generation Sequencing.” Journal of Clinical Oncology 36(7):633–41. |
| 75 | Rizvi, Naiyer A., Matthew D. Hellmann, Alexandra Snyder, Pia Kvistborg, Vladimir Makarov, Jonathan J. Havel, William Lee, Jianda Yuan, Phillip Wong, Teresa S. Ho, Martin L. Miller, Natasha Rekhtman, Andre L. Moreira, Fawzia Ibrahim, Cameron Bruggeman, Billel Gasmi, Roberta Zappasodi, Yuka Maeda, Chris Sander, Edward B. Garon, Taha Merghoub, Jedd D. Wolchok, Ton N. Schumacher, and Timothy A. Chan. 2015. “Mutational Landscape Determines Sensitivity to PD-1 Blockade in Non-Small Cell Lung Cancer.” Science 348(6230):124–28. |
| 76 | Robert, Caroline, Georgina V. Long, Benjamin Brady, Caroline Dutriaux, Michele Maio, Laurent Mortier, Jessica C. Hassel, Piotr Rutkowski, Catriona McNeil, Ewa Kalinka-Warzocha, Kerry J. Savage, Micaela M. Hernberg, Celeste Lebbé, Julie Charles, Catalin Mihalcioiu, Vanna Chiarion-Sileni, Cornelia Mauch, Francesco Cognetti, Ana Arance, Henrik Schmidt, Dirk Schadendorf, Helen Gogas, Lotta Lundgren-Eriksson, Christine Horak, Brian Sharkey, Ian M. Waxman, Victoria Atkinson, and Paolo A. Ascierto. 2015. “Nivolumab in Previously Untreated Melanoma without BRAF Mutation.” New England Journal of Medicine 372(4):320–30. |
| 77 | Rosenberg, Jonathan E., Jean Hoffman-Censits, Tom Powles, Michiel S. Van Der Heijden, Arjun V. Balar, Andrea Necchi, Nancy Dawson, Peter H. O’Donnell, Ani Balmanoukian, Yohann Loriot, Sandy Srinivas, Margitta M. Retz, Petros Grivas, Richard W. Joseph, Matthew D. Galsky, Mark T. Fleming, Daniel P. Petrylak, Jose Luis Perez-Gracia, Howard A. Burris, Daniel Castellano, Christina Canil, |

| | |
|---|---|
| | Joaquim Bellmunt, Dean Bajorin, Dorothee Nickles, Richard Bourgon, Garrett M. Frampton, Na Cui, Sanjeev Mariathasan, Oyewale Abidoye, Gregg D. Fine, and Robert Dreicer. 2016. “Atezolizumab in Patients with Locally Advanced and Metastatic Urothelial Carcinoma Who Have Progressed Following Treatment with Platinum-Based Chemotherapy: A Single-Arm, Multicentre, Phase 2 Trial.” The Lancet 387(10031):1909–20. |
| 78 | Routy, Bertrand, Emmanuelle Le Chatelier, Lisa Derosa, Connie P. M. Duong, Maryam Tidjani Alou, Romain Daillère, Aurélie Fluckiger, Meriem Messaoudene, Conrad Rauber, Maria P. Roberti, Marine Fidelle, Caroline Flament, Vichnou Poirier-Colame, Paule Opolon, Christophe Klein, Kristina Iribarren, Laura Mondragón, Nicolas Jacquelot, Bo Qu, Gladys Ferrere, Céline Clémenson, Laura Mezquita, Jordi Remon Masip, Charles Naltet, Solenn Brosseau, Coureche Kaderbhai, Corentin Richard, Hira Rizvi, Florence Levenez, Nathalie Galleron, Benoit Quinquis, Nicolas Pons, Bernhard Ryffel, Véronique Minard-Colin, Patrick Gonin, Jean Charles Soria, Eric Deutsch, Yohann Loriot, François Ghiringhelli, Gérard Zalcman, François Goldwasser, Bernard Escudier, Matthew D. Hellmann, Alexander Eggermont, Didier Raoult, Laurence Albiges, Guido Kroemer, and Laurence Zitvogel. 2018. “Gut Microbiome Influences Efficacy of PD-1-Based Immunotherapy against Epithelial Tumors.” Science 359(6371):91–97. |
| 79 | Schmid, Peter, Sylvia Adams, Hope S. Rugo, Andreas Schneeweiss, Carlos H. Barrios, Hiroji Iwata, Véronique Diéras, Roberto Hegg, Seock-Ah Im, Gail Shaw Wright, Volkmar Henschel, Luciana Molinero, Stephen Y. Chui, Roel Funke, Amreen Husain, Eric P. Winer, Sherene Loi, and Leisha A. Emens. 2018. “Atezolizumab and Nab-Paclitaxel in Advanced Triple-Negative Breast Cancer.” New England Journal of Medicine 379(22):2108–21. |
| 80 | Shah, Manish A., Takashi Kojima, Daniel Hochhauser, Peter Enzinger, Judith Raimbourg, Antoine Hollebecque, Florian Lordick, Sung Bae Kim, Masahiro Tajika, Heung Tae Kim, A. Craig Lockhart, Hendrik Tobias Arkenau, Farid El-Hajbi, Mukul Gupta, Per Pfeiffer, Qi Liu, Jared Lunceford, S. Peter Kang, Pooja Bhagia, and Ken Kato. 2019. “Efficacy and Safety of Pembrolizumab for Heavily Pretreated Patients with Advanced, Metastatic Adenocarcinoma or Squamous Cell Carcinoma of the Esophagus: The Phase 2 KEYNOTE-180 Study.” JAMA Oncology 5(4):546–50. |
| 81 | Sharma, Padmanee, Margitta Retz, Arlene Siefker-Radtke, Ari Baron, Andrea Necchi, Jens Bedke, Elizabeth R. Plimack, Daniel Vaena, Marc Oliver Grimm, Sergio Bracarda, José Ángel Arranz, Sumanta Pal, Chikara Ohyama, Abdel Saci, Xiaotao Qu, Alexandre Lambert, Suba Krishnan, Alex Azrilevich, and Matthew D. Galsky. 2017. “Nivolumab in Metastatic Urothelial Carcinoma after Platinum Therapy (CheckMate 275): A Multicentre, Single-Arm, Phase 2 Trial.” The Lancet Oncology 18(3):312–22. |
| 82 | Singal G, Miller PG, Agarwala V, et al. Analyzing biomarkers of cancer immunotherapy (CIT) response using a real-world clinico-genomic database. *Ann Oncol*. 2017;28:v404-v405. doi:10.1093/annonc/mdx376.005 |
| 83 | Socinski, M. A., R. M. Jotte, F. Cappuzzo, F. Orlandi, D. Stroyakovskiy, N. Nogami, D. Rodriguez-Abreu, D. Moro-Sibilot, C. A. Thomas, F. Barlesi, G. Finley, C. Kelsch, A. Lee, S. Coleman, Y. Deng, Y. Shen, M. Kowanetz, A. Lopez-Chave, A. Sandler, and M. Reck. 2018. “Atezolizumab for First-Line Treatment of Metastatic Nonsquamous NSCLC.” New England Journal of Medicine 378(24):2288–2301. |
| 84 | Spigel DR, Gettinger SN, Horn L, et al. Clinical activity, safety, and biomarkers of MPDL3280A, an engineered PD-L1 antibody in patients with locally advanced or metastatic non-small cell lung cancer (NSCLC). *J Clin Oncol*. 2013;31(15_suppl):8008-8008. doi:10.1200/jco.2013.31.15_suppl.8008 |

| | |
|---|---|
| 85 | Taube, Janis M., Alison Klein, Julie R. Brahmer, Haiying Xu, Xiaoyu Pan, Jung H. Kim, Lieping Chen, Drew M. Pardoll, Suzanne L. Topalian, and Robert A. Anders. 2014. “Association of PD-1, PD-1 Ligands, and Other Features of the Tumor Immune Microenvironment with Response to Anti-PD-1 Therapy.” Clinical Cancer Research 20(19):5064–74. |
| 86 | Topalian, Suzanne L., F. Stephen Hodi, Julie R. Brahmer, Scott N. Gettinger, David C. Smith, David F. McDermott, John D. Powderly, Richard D. Carvajal, Jeffrey A. Sosman, Michael B. Atkins, Philip D. Leming, David R. Spigel, Scott J. Antonia, Leora Horn, Charles G. Drake, Drew M. Pardoll, Lieping Chen, William H. Sharfman, Robert A. Anders, Janis M. Taube, Tracee L. McMiller, Haiying Xu, Alan J. Korman, Maria Jure-Kunkel, Shruti Agrawal, Daniel McDonald, Georgia D. Kollia, Ashok Gupta, Jon M. Wigginton, and Mario Sznol. 2012. “Safety, Activity, and Immune Correlates of Anti-PD-1 Antibody in Cancer.” New England Journal of Medicine 366(26):2443–54. |
| 87 | Tumeh, Paul C., Christina L. Harview, Jennifer H. Yearley, I. Peter Shintaku, Emma J. M. Taylor, Lidia Robert, Bartosz Chmielowski, Marko Spasic, Gina Henry, Voicu Ciobanu, Alisha N. West, Manuel Carmona, Christine Kivork, Elizabeth Seja, Grace Cherry, Antonio J. Gutierrez, Tristan R. Grogan, Christine Mateus, Gorana Tomasic, John A. Glaspy, Ryan O. Emerson, Harlan Robins, Robert H. Pierce, David A. Elashoff, Caroline Robert, and Antoni Ribas. 2014. “PD-1 Blockade Induces Responses by Inhibiting Adaptive Immune Resistance.” Nature 515(7528):568–71. |
| 88 | Tykodi, Scott S., Frede Donskov, Jae-Lyun Lee, Cezary Szczylik, Jahangeer Malik, Boris Yakovlevich Alekseev, James M. G. Larkin, Vsevolod Borisovich Matveev, Rustem Gafanov, Piotr Tomczak, Poul F. Geertsen, Pawel J. Wiechno, Sang Joon Shin, Frederic Pouliot, Teresa Alonso-Gordoa, Rachel Kloss Silverman, Rodolfo F. Perini, Charles Schloss, David F. McDermott, and Michael B. Atkins. 2019. “First-Line Pembrolizumab (Pembro) Monotherapy in Advanced Clear Cell Renal Cell Carcinoma (CcRCC): Updated Results for KEYNOTE-427 Cohort A.” Journal of Clinical Oncology 37(15_suppl):4570–4570. |
| 89 | Wang, Li, Abdel Saci, Peter M. Szabo, Scott D. Chasalow, Mireia Castillo-Martin, Josep Domingo-Domenech, Arlene Siefker-Radtke, Padmanee Sharma, John P. Sfakianos, Yixuan Gong, Ana Dominguez-Andres, William K. Oh, David Mulholland, Alex Azrilevich, Liangyuan Hu, Carlos Cordon-Cardo, Hélène Salmon, Nina Bhardwaj, Jun Zhu, and Matthew D. Galsky. 2018. “EMT- and Stroma-Related Gene Expression and Resistance to PD-1 Blockade in Urothelial Cancer.” Nature Communications 9(1). |
| 90 | Weber, Jeffrey S., Sandra P. D’Angelo, David Minor, F. Stephen Hodi, Ralf Gutzmer, Bart Neyns, Christoph Hoeller, Nikhil I. Khushalani, Wilson H. Miller, Christopher D. Lao, Gerald P. Linette, Luc Thomas, Paul Lorigan, Kenneth F. Grossmann, Jessica C. Hassel, Michele Maio, Mario Sznol, Paolo A. Ascierto, Peter Mohr, Bartosz Chmielowski, Alan Bryce, Inge M. Svane, Jean Jacques Grob, Angela M. Krackhardt, Christine Horak, Alexandre Lambert, Arvin S. Yang, and James Larkin. 2015. “Nivolumab versus Chemotherapy in Patients with Advanced Melanoma Who Progressed after Anti-CTLA-4 Treatment (CheckMate 037): A Randomised, Controlled, Open-Label, Phase 3 Trial.” The Lancet Oncology 16(4):375–84. |
| 91 | Weber, Jeffrey S., Ragini Reiney Kudchadkar, Bin Yu, Donna Gallenstein, Christine E. Horak, H. David Inzunza, Xiuhua Zhao, Alberto J. Martinez, Wenshi Wang, Geoffrey Gibney, Jodi Kroeger, Cabell Eysmans, Amod A. Sarnaik, and Y. Ann Chen. 2013. “Safety, Efficacy, and Biomarkers of Nivolumab with Vaccine in Ipilimumab-Refractory or -Naive Melanoma.” Journal of Clinical Oncology 31(34):4311–18. |
| 92 | Wolchok JD, Kluger H, Callahan MK, et al. Nivolumab plus Ipilimumab in Advanced Melanoma. *N Engl J Med*. 2013;369(2):122-133. doi:10.1056/nejmoa1302369 |
| 93 | Wolchok, J. D., V. Chiarion-Sileni, R. Gonzalez, P. Rutkowski, J. J. Grob, C. L. Cowey, C. D. Lao, J. Wagstaff, D. Schadendorf, P. F. Ferrucci, M. Smylie, R. Dummer, A. Hill, D. Hogg, J. Haanen, M. S. Carlino, O. Bechter, M. Maio, I. Marquez-Rodas, M. Guidoboni, G. |

| | |
|---|---|
| | McArthur, C. Lebbé, P. A. Ascierto, G. V. Long, J. Cebon, J. Sosman, M. A. Postow, M. K. Callahan, D. Walker, L. Rollin, R. Bhore, F. S. Hodi, and J. Larkin. 2017. "Overall Survival with Combined Nivolumab and Ipilimumab in Advanced Melanoma." New England Journal of Medicine 377(14):1345–56. |
| 94 | Younes, Anas, Armando Santoro, Margaret Shipp, Pier Luigi Zinzani, John M. Timmerman, Stephen Ansell, Philippe Armand, Michelle Fanale, Voravit Ratanatharathorn, John Kuruvilla, Jonathon B. Cohen, Graham Collins, Kerry J. Savage, Marek Trneny, Kazunobu Kato, Benedetto Farsaci, Susan M. Parker, Scott Rodig, Margaretha G. M. Roemer, Azra H. Ligon, and Andreas Engert. 2016. "Nivolumab for Classical Hodgkin's Lymphoma after Failure of Both Autologous Stem-Cell Transplantation and Brentuximab Vedotin: A Multicentre, Multicohort, Single-Arm Phase 2 Trial." The Lancet Oncology 17(9):1283–94. |
| 95 | Zhu, Andrew X., Richard S. Finn, Julien Edeline, Stephane Cattan, Sadahisa Ogasawara, Daniel Palmer, Chris Verslype, Vittorina Zagonel, Laetitia Fartoux, Arndt Vogel, Debashis Sarker, Gontran Verset, Stephen L. Chan, Jennifer Knox, Bruno Daniele, Andrea L. Webber, Scot W. Ebbinghaus, Junshui Ma, Abby B. Siegel, Ann Lii Cheng, Masatoshi Kudo, Angela Alistar, Jamil Asselah, Jean Frederic Blanc, Ivan Borbath, Timothy Cannon, Ki Chung, Allen Cohn, David P. Cosgrove, Nevena Damjanov, Mukul Gupta, Yoshivasu Karino, Mark Karwal, Andreas Kaubisch, Robin Kelley, Jena Luc Van Laethem, Timothy Larson, James Lee, Daneng Li, Atisha Manhas, Gulam Abbas Manji, Kazushi Numata, Benjamin Parsons, Andrew S. Paulson, Carmine Pinto, Robert Ramirez, Suresh Ratnam, Magnus Rizell, Olivier Rosmorduc, Yvonne Sada, Yutaka Sasaki, Per I. Stal, Simone Strasser, Joerg Trojan, Gina Vaccaro, Hans Van Vlierberghe, Alan Weiss, Karl Heinz Weiss, and Tatsuya Yamashita. 2018. "Pembrolizumab in Patients with Advanced Hepatocellular Carcinoma Previously Treated with Sorafenib (KEYNOTE-224): A Non-Randomised, Open-Label Phase 2 Trial." The Lancet Oncology 19(7):940–52. |

**Supplementary Table 3. Breakdown of biomarkers in the dataset**

| Biomarker Acronym | Biomarkers | Studies N | N |
|---|---|---|---|
| PD-L1 IHC | PD-L1 protein expression immunohistochemistry assay | 76 | 13909 |
| TMB | Tumor mutational burden | 15 | 1814 |
| Others | | 10 | 1001 |
| GEP | Gene expression profiles | 5 | 783 |
| Multimodal | Multimodal biomarkers | 5 | 559 |
| MB | Microbiome | 3 | 228 |
| mIHC/IF | PD-L1 protein expression fluorescent multiplex immunohistochemistry assay | 3 | 177 |
| IMDC | International Metastatic RCC Database Consortium Risk Score | 2 | 547 |
| AEs | Adverse events | 1 | 1747 |
| Others[1] | CD20+ tumor infiltrating level score, CD4<CD8 tumor infiltrating cells, CD8+ cell density, circulating tumor mutational DNA, IDO-1 or HLA-DR expression, Immune infiltrate score, somatic insertions/deletions (indels), lymphocyte counts, lymphoid aggregates, microsatellite instability, necrosis, PD-L2 expression, PD-1 protein expression, partially exhausted cytotoxic T lymphocytes | 10 | 1001 |

[1]Biomarkers were placed into the other category if the number of studies investigating the relevant biomarker were < 3 and the total number of subjects across all studies was < 500.

**Supplementary Table 4. Breakdown of cancer types in the dataset**

| Cancer Type | Biomarkers Explored | Studies (N) | Percentage (%) |
|---|---|---|---|
| Non-Small-Cell Lung Cancer | PD-L1 IHC, TMB, Multimodal, GEP | 26 | 27.40 |
| Melanoma | TMB, GEP, PD-L1 IHC, Multimodal, MB, Others, Mutation, mIHC/IF, AEs | 22 | 23.20 |
| Multiple Cancers | Mutation, TMB, GEP, PD-L1 IHC, Multimodal, Others, MB | 10 | 10.50 |
| Urothelial Cancer | PD-L1 IHC, AEs, Mutation, Others, GEP | 9 | 9.47 |
| Head and Neck Cancer | PD-L1e, GEP, TMB, Multimodal, Others | 8 | 8.42 |
| Colorectal Cancer | PD-L1 IHC, Mutation, Others | 5 | 5.26 |
| Renal Cell Carcinoma | TMB, Mutation, GEP, Others, PD-L1 IHC, IMDC | 3 | 4.21 |
| Merkel cell | Others, mIHC/IF, PD-L1 IHC | 3 | 3.16 |
| Breast | PD-L1 IHC, Mutation | 3 | 3.16 |
| Gastric and Gastroesophageal Junction | PD-L1 IHC, Others | 3 | 3.16 |
| Small-Cell Lung Cancer | PD-L1 IHC, TMB | 3 | 3.16 |
| Hepatocellular Carcinoma | PD-L1 IHC | 2 | 2.11 |
| B-Cell Lymphoma | PD-L1 IHC | 1 | 1.05 |
| Cervical cancer | PD-L1 IHC | 1 | 1.05 |
| Hodgkin's Lymphoma | PD-L1 IHC | 1 | 1.05 |

#### Supplementary Table 5. Meta-analyzed estimates of biomarkers by cancer types

| Cancer | Biomarker[1] | Reference | Studies N[2] | N | pAUC[3] | gAUC[3] | Sensitivity | False positive rate | Specificity | False negative rate | Positive predictive value | False omission rate | Negative predictive value | False discovery rate |
|---|---|---|---|---|---|---|---|---|---|---|---|---|---|---|
| Breast | PD-L1 IHC | 1,18,79 | 3 | 756 | 0.44 (0.25-0.65) | 0.52 (0.35-0.69) | 0.43 (0.24-0.65) | 0.30 (0.07-0.71) | 0.70 (0.29-0.93) | 0.57 (0.35-0.76) | 0.25 (0.08-0.57) | 0.19 (0.06-0.47) | 0.81 (0.53-0.94) | 0.75 (0.43-0.92) |
| Colon | PD-L1 IHC | 21,62,61 | 3 | 342 | 0.53 (0.27-0.66) | 0.56 (0.41-0.65) | 0.46 (0.27-0.66) | 0.40 (0.29-0.52) | 0.60 (0.48-0.71) | 0.54 (0.34-0.73) | 0.17 (0.05-0.47) | 0.13 (0.03-0.43) | 0.87 (0.57-0.97) | 0.83 (0.53-0.95) |
| Gastric and gastroesophageal junction | PD-L1 IHC | 24,42,59 | 3 | 325 | 0.47 (0.25-0.69) | 0.61 (0.41-0.74) | 0.46 (0.24-0.69) | 0.28 (0.12-0.53) | 0.72 (0.47-0.88) | 0.54 (0.31-0.76) | 0.34 (0.25-0.43) | 0.20 (0.13-0.28) | 0.80 (0.72-0.87) | 0.66 (0.57-0.75) |
| HCC | PD-L1 IHC | 19,95 | 2 | 272 | 0.33 (0.16-0.76) | 0.6 (0.38-0.72) | 0.43 (0.15-0.75) | 0.36 (0.23-0.52) | 0.64 (0.48-0.77) | 0.57 (0.25-0.85) | 0.41 (0.20-0.65) | 0.33 (0.08-0.74) | 0.67 (0.26-0.92) | 0.59 (0.35-0.80) |
| Head and neck | Others | 52,55 | 2 | 236 | 0.71 (0.33-0.93) | 0.71 (0.45-0.89) | 0.79 (0.39-0.96) | 0.43 (0.21-0.69) | 0.57 (0.31-0.79) | 0.21 (0.04-0.61) | 0.53 (0.21-0.82) | 0.19 (0.08-0.39) | 0.81 (0.61-0.92) | 0.47 (0.18-0.79) |
| Head and neck | PD-L1 IHC | 5,11,14,15,23,55,81 | 7 | 1111 | 0.73 (0.63-0.85) | 0.55 (0.45-0.68) | 0.81 (0.70-0.88) | 0.70 (0.61-0.78) | 0.30 (0.22-0.39) | 0.19 (0.12-0.30) | 0.23 (0.18-0.30) | 0.15 (0.12-0.20) | 0.84 (0.80-0.88) | 0.77 (0.70-0.82) |
| Head and neck | GEP | 15 | 1 | 105 | NA | NA | 0.95 | 0.65 | 0.35 | 0.05 | 0.27 | 0.03 | 0.97 | 0.73 |
| Head and neck | TMB | 15 | 1 | 105 | NA | NA | 0.71 | 0.45 | 0.55 | 0.29 | 0.28 | 0.12 | 0.88 | 0.72 |
| Head and neck | Multimodal (TMB and GEP) | 15 | 1 | 105 | NA | NA | 0.67 | 0.29 | 0.71 | 0.33 | 0.37 | 0.1 | 0.9 | 0.63 |
| Head and neck | Multimodal (TMB and PD-L1 IHC) | 15 | 1 | 107 | NA | NA | 0.67 | 0.38 | 0.62 | 0.33 | 0.3 | 0.12 | 0.88 | 0.7 |
| Melanoma | MB | 32,53 | 2 | 128 | 0.64 (0.14-0.83) | 0.83 (0.33-0.97) | 0.58 (0.21-0.88) | 0.10 (0.01-0.50) | 0.90 (0.50-0.99) | 0.42 (0.12-0.79) | 0.87 (0.48-0.98) | 0.35 (0.07-0.79) | 0.65 (0.21-0.93) | 0.13 (0.02-0.52) |
| Melanoma | mIHC/IF | 43,87 | 2 | 157 | 0.82 (0.31-0.93) | 0.82 (0.53-0.93) | 0.84 (0.62-0.95) | 0.33 (0.16-0.56) | 0.67 (0.44-0.84) | 0.16 (0.05-0.38) | 0.77 (0.57-0.90) | 0.22 (0.07-0.51) | 0.78 (0.49-0.93) | 0.23 (0.10-0.43) |
| Melanoma | Multimodal | 15,43 | 2 | 231 | 0.38 (0.25-0.83) | 0.65 (0.56-0.73) | 0.64 (0.51-0.75) | 0.38 (0.30-0.46) | 0.62 (0.54-0.70) | 0.36 (0.25-0.49) | 0.61 (0.52-0.69) | 0.35 (0.24-0.47) | 0.65 (0.53-0.76) | 0.39 (0.31-0.48) |
| Melanoma | PD-L1 IHC | 15,16,17,33,47,50,51,66,69,76,91,90,93 | 14 | 2313 | 0.63 (0.49-0.72) | 0.6 (0.53-0.66) | 0.65 (0.52-0.76) | 0.49 (0.37-0.61) | 0.51 (0.39-0.63) | 0.35 (0.24-0.48) | 0.50 (0.45-0.56) | 0.34 (0.26-0.44) | 0.66 (0.56-0.74) | 0.50 (0.44-0.55) |
| Melanoma | TMB | 15,31,44,71 | 4 | 227 | 0.38 (0.24-0.93) | 0.37 (0.33-0.83) | 0.86 (0.72-0.94) | 0.64 (0.56-0.71) | 0.36 (0.29-0.44) | 0.14 (0.06-0.28) | 0.57 (0.50-0.63) | 0.28 (0.14-0.48) | 0.72 (0.52-0.86) | 0.43 (0.37-0.50) |
| Merkel cell | PD-L1 IHC | 46,45,60 | 3 | 194 | 0.73 (0.58-0.9) | 0.51 (0.3-0.8) | 0.83 (0.66-0.92) | 0.73 (0.59-0.84) | 0.27 (0.16-0.41) | 0.17 (0.08-0.34) | 0.50 (0.37-0.63) | 0.38 (0.20-0.60) | 0.62 (0.40-0.80) | 0.50 (0.37-0.63) |
| Multiple cancers | Others | 48,85 | 2 | 107 | 0.65 (0.39-0.74) | 0.66 (0.56-0.72) | 0.61 (0.40-0.78) | 0.36 (0.24-0.50) | 0.64 (0.50-0.76) | 0.39 (0.22-0.60) | 0.40 (0.32-0.49) | 0.22 (0.16-0.28) | 0.78 (0.72-0.84) | 0.60 (0.51-0.68) |
| Multiple cancers | PD-L1 IHC | 15,39,80,81,85,86 | 6 | 693 | 0.61 (0.49-0.78) | 0.64 (0.55-0.72) | 0.68 (0.54-0.79) | 0.45 (0.36-0.54) | 0.55 (0.46-0.64) | 0.32 (0.21-0.46) | 0.33 (0.24-0.43) | 0.15 (0.09-0.26) | 0.85 (0.74-0.91) | 0.67 (0.57-0.76) |
| Multiple cancers | TMB | 7,15,31 | 3 | 231 | 0.81 (0.48-0.92) | 0.75 (0.58-0.87) | 0.84 (0.65-0.94) | 0.41 (0.27-0.56) | 0.59 (0.44-0.73) | 0.16 (0.06-0.35) | 0.63 (0.33-0.86) | 0.17 (0.04-0.53) | 0.83 (0.47-0.96) | 0.37 (0.14-0.67) |
| Non-small-cell lung cancer | Multimodal | 10,37,74 | 3 | 304 | 0.4 (0.21-0.67) | 0.71 (0.43-0.9) | 0.43 (0.23-0.65) | 0.12 (0.04-0.31) | 0.88 (0.69-0.96) | 0.57 (0.35-0.77) | 0.61 (0.48-0.73) | 0.24 (0.19-0.30) | 0.76 (0.70-0.81) | 0.39 (0.27-0.52) |

| | | | | | | | | | | | | | | |
|---|---|---|---|---|---|---|---|---|---|---|---|---|---|---|
| Non-small-cell lung cancer | PD-L1 IHC | 8,10,22,25,27,26,28,29,37,35,34,39,38,40,49,56,64,70,73,74,83,84 | 22 | 5623 | 0.56 (0.56-0.71) | 0.61 (0.56-0.66) | 0.67 (0.62-0.72) | 0.54 (0.46-0.61) | 0.46 (0.39-0.54) | 0.33 (0.28-0.38) | 0.45 (0.38-0.51) | 0.31 (0.23-0.40) | 0.69 (0.60-0.77) | 0.55 (0.49-0.62) |
| Non-small-cell lung cancer | TMB | 9,10,25,31,37,70,75,74,82 | 9 | 1285 | 0.36 (0.3-0.75) | 0.7 (0.6-0.74) | 0.58 (0.46-0.69) | 0.31 (0.25-0.37) | 0.69 (0.63-0.75) | 0.42 (0.31-0.54) | 0.58 (0.50-0.66) | 0.31 (0.23-0.40) | 0.69 (0.60-0.77) | 0.42 (0.34-0.50) |
| Renal cell carcinoma | PD-L1 IHC | 57,58,72,88 | 4 | 1160 | 0.53 (0.37-0.63) | 0.51 (0.45-0.57) | 0.50 (0.40-0.61) | 0.48 (0.38-0.58) | 0.52 (0.42-0.62) | 0.50 (0.39-0.60) | 0.68 (0.48-0.83) | 0.66 (0.40-0.85) | 0.34 (0.15-0.60) | 0.32 (0.17-0.52) |
| Renal cell carcinoma | TMB | 54 | 1 | 71 | 0.66 (0.52-0.8) | 0.53 (0.36-0.68) | 0.70 (0.57-0.81) | 0.66 (0.52-0.77) | 0.34 (0.23-0.48) | 0.30 (0.19-0.43) | 0.50 (0.37-0.62) | 0.45 (0.29-0.62) | 0.55 (0.38-0.71) | 0.50 (0.38-0.63) |
| Small-cell lung cancer | PD-L1 IHC | 2,12,35 | 3 | 280 | 0.61 (0.21-0.87) | 0.59 (0.27-0.84) | 0.59 (0.13-0.93) | 0.45 (0.10-0.85) | 0.55 (0.15-0.90) | 0.41 (0.07-0.87) | 0.26 (0.16-0.39) | 0.13 (0.06-0.29) | 0.87 (0.71-0.94) | 0.74 (0.61-0.84) |
| Urothelial cancer | PD-L1 IHC | 4,6,63,65,67,68,77 | 7 | 1041 | 0.56 (0.39-0.71) | 0.56 (0.49-0.62) | 0.57 (0.39-0.74) | 0.48 (0.33-0.63) | 0.52 (0.37-0.67) | 0.43 (0.26-0.61) | 0.37 (0.23-0.53) | 0.29 (0.17-0.44) | 0.71 (0.56-0.83) | 0.63 (0.47-0.77) |
| B-Cell Lymphoma | PD-L1 IHC | 3 | 1 | 42 | NA | NA | 0.89 | 0.74 | 0.26 | 0.11 | 0.5 | 0.25 | 0.75 | 0.5 |
| Cervical cancer | PD-L1 IHC | 13 | 1 | 84 | NA | NA | 0.8 | 0.86 | 0.14 | 0.2 | 0.17 | 0.23 | 0.77 | 0.83 |
| Melanoma | GEP | 15 | 1 | 86 | NA | NA | 0.84 | 0.65 | 0.35 | 0.16 | 0.51 | 0.26 | 0.74 | 0.49 |
| Multiple cancers | GEP | 15 | 1 | 113 | NA | NA | 0.94 | 0.55 | 0.45 | 0.06 | 0.22 | 0.02 | 0.98 | 0.78 |
| Multiple cancers | Multimodal (TMB and GEP) | 15 | 1 | 113 | NA | NA | 0.62 | 0.18 | 0.82 | 0.38 | 0.37 | 0.07 | 0.93 | 0.63 |
| Multiple cancers | Multimodal (TMB and PD-L1 IHC) | 15 | 1 | 74 | NA | NA | 0.78 | 0.2 | 0.8 | 0.22 | 0.35 | 0.04 | 0.96 | 0.65 |
| Colon | Others (anti-PD-1 and MEK treatments) | 21 | 1 | 90 | NA | NA | 0.5 | 0.02 | 0.98 | 0.5 | 0.33 | 0.01 | 0.99 | 0.67 |
| Colon | Others (anti_PD-1 treatment) | 21 | 1 | 183 | NA | NA | 0.4 | 0.01 | 0.99 | 0.6 | 0.67 | 0.02 | 0.98 | 0.33 |
| Merkel cell | Others | 30 | 1 | 23 | NA | NA | 0.81 | 0.57 | 0.43 | 0.19 | 0.76 | 0.5 | 0.5 | 0.24 |
| Merkel cell | mIHC/IF | 30 | 1 | 20 | NA | NA | 0.93 | 0.33 | 0.67 | 0.07 | 0.87 | 0.2 | 0.8 | 0.13 |
| Small-cell lung cancer | TMB (anti-PD-1 and anti-CTLA-4 treatments) | 35 | 1 | 133 | NA | NA | 0.87 | 0.66 | 0.34 | 0.13 | 0.14 | 0.05 | 0.95 | 0.86 |
| Small-cell lung cancer | TMB (anti-PD-1 treatment) | 35 | 1 | 78 | NA | NA | 0.75 | 0.62 | 0.38 | 0.25 | 0.29 | 0.19 | 0.81 | 0.71 |
| Melanoma | GEP | 41 | 1 | 27 | NA | NA | 0.93 | 0.31 | 0.69 | 0.07 | 0.76 | 0.1 | 0.9 | 0.24 |

| | | | | | | | | | | | | | | |
|---|---|---|---|---|---|---|---|---|---|---|---|---|---|---|
| Melanoma | Others (discovery) | 43 | 1 | 24 | NA | NA | 0.85 | 0.09 | 0.91 | 0.15 | 0.92 | 0.17 | 0.83 | 0.08 |
| Melanoma | Others (validation) | 43 | 1 | 142 | NA | NA | 0.6 | 0.28 | 0.72 | 0.4 | 0.72 | 0.4 | 0.6 | 0.28 |
| Urothelial cancer | AEs | 20 | 1 | 1747 | NA | NA | 0.36 | 0.34 | 0.66 | 0.64 | 0.21 | 0.19 | 0.81 | 0.79 |
| Renal cell carcinoma | GEP (angiogenesis gene signature) | 54 | 1 | 86 | NA | NA | 0.56 | 0.52 | 0.48 | 0.44 | 0.41 | 0.38 | 0.62 | 0.59 |
| Renal cell carcinoma | Others (anti-PD-1 treatment) | 54 | 1 | 71 | NA | NA | 0.71 | 0.64 | 0.36 | 0.29 | 0.36 | 0.29 | 0.71 | 0.64 |
| Renal cell carcinoma | GEP (T-cell effector gene signature) | 54 | 1 | 88 | NA | NA | 0.59 | 0.35 | 0.65 | 0.41 | 0.7 | 0.47 | 0.53 | 0.3 |
| Renal cell carcinoma | Others (anti-PD-1 treatment and chemotherapy) | 54 | 1 | 65 | NA | NA | 0.49 | 0.62 | 0.38 | 0.51 | 0.57 | 0.7 | 0.3 | 0.43 |
| Renal cell carcinoma | IMDC | 58 | 1 | 437 | NA | NA | 0.9 | 0.76 | 0.24 | 0.1 | 0.56 | 0.31 | 0.69 | 0.44 |
| Multiple cancers | MB | 78 | 1 | 100 | NA | NA | 0.22 | 0.1 | 0.9 | 0.78 | 0.69 | 0.45 | 0.55 | 0.31 |
| Non-small-cell lung cancer | GEP | 15 | 1 | 341 | NA | NA | 0.49 | 0.39 | 0.61 | 0.51 | 0.72 | 0.62 | 0.38 | 0.28 |
| Renal cell carcinoma | IMDC | 88 | 1 | 110 | NA | NA | 0.41 | 0.34 | 0.66 | 0.59 | 0.62 | 0.54 | 0.46 | 0.38 |
| Urothelial cancer | Others | 89 | 1 | 214 | NA | NA | 0.67 | 0.45 | 0.55 | 0.33 | 0.31 | 0.15 | 0.85 | 0.69 |
| Urothelial cancer | GEP | 89 | 1 | 214 | NA | NA | 0.51 | 0.5 | 0.5 | 0.49 | 0.23 | 0.22 | 0.78 | 0.77 |
| Gastric and gastroesophageal junction | Others (anti-PD-1nand anti-CTLA-4 treatments) | 42 | 1 | 25 | NA | NA | 0.5 | 0.24 | 0.76 | 0.5 | 0.29 | 0.11 | 0.89 | 0.71 |
| Gastric and gastroesophageal junction | Others (anti-PD-1 treatment) | 42 | 1 | 23 | NA | NA | 0.2 | 0.06 | 0.94 | 0.8 | 0.5 | 0.19 | 0.81 | 0.5 |
| Hodgkin's lymphoma | PD-L1 IHC | 94 | 1 | 40 | NA | NA | 0.78 | 0.33 | 0.67 | 0.22 | 0.97 | 0.8 | 0.2 | 0.03 |

[1]differences between cohorts are described.

[2]number of unique studies. If multiple cohorts were described in a study, those were meta-analyzed. This is how meta-analyzed results can be shown for a single study.

[3]estimates were not meta-analyzed if pAUC or gAUC are listed as NA, due to less than three cohorts available.

#### Supplementary Table 6. Meta-analyzed estimates of biomarkers for all cancer types (with N>500)

| Biomarker | Studies N[1] | N | pAUC[2] | gAUC[2] | Sensitivity | False positive rate | Specificity | False negative rate | Positive predictive value | False omission rate | Negative predictive value | False discovery rate |
|---|---|---|---|---|---|---|---|---|---|---|---|---|
| AEs | 1 | 1747 | #N/A | #N/A | 0.36 | 0.34 | 0.66 | 0.64 | 0.21 | 0.19 | 0.81 | 0.79 |
| GEP | 5 | 783 | 0.6 (0.42-0.83) | 0.56 (0.49-0.65) | 0.71 (0.55-0.84) | 0.52 (0.44-0.59) | 0.48 (0.41-0.56) | 0.29 (0.16-0.45) | 0.45 (0.30-0.61) | 0.27 (0.14-0.46) | 0.73 (0.54-0.86) | 0.55 (0.39-0.70) |
| IMDC | 2 | 547 | 0.68 (0.2-0.95) | 0.57 (0.32-0.83) | 0.72 (0.17-0.97) | 0.57 (0.17-0.89) | 0.43 (0.11-0.83) | 0.28 (0.03-0.83) | 0.59 (0.51-0.66) | 0.42 (0.21-0.66) | 0.58 (0.34-0.79) | 0.41 (0.34-0.49) |
| Multimodal | 5 | 559 | 0.52 (0.45-0.69) | 0.7 (0.61-0.76) | 0.57 (0.47-0.67) | 0.24 (0.17-0.34) | 0.76 (0.66-0.83) | 0.43 (0.33-0.53) | 0.50 (0.41-0.59) | 0.19 (0.13-0.29) | 0.81 (0.71-0.87) | 0.50 (0.41-0.59) |
| Others | 10 | 1001 | 0.59 (0.52-0.69) | 0.67 (0.58-0.71) | 0.60 (0.50-0.69) | 0.31 (0.21-0.44) | 0.69 (0.56-0.79) | 0.40 (0.31-0.50) | 0.46 (0.37-0.55) | 0.19 (0.13-0.28) | 0.81 (0.72-0.87) | 0.54 (0.45-0.63) |
| PD-L1 IHC | 76 | 13909 | 0.53 (0.53-0.65) | 0.58 (0.56-0.6) | 0.64 (0.60-0.69) | 0.51 (0.47-0.56) | 0.49 (0.44-0.53) | 0.36 (0.31-0.40) | 0.40 (0.36-0.45) | 0.28 (0.24-0.33) | 0.72 (0.67-0.76) | 0.60 (0.55-0.64) |
| TMB | 15 | 1814 | 0.57 (0.52-0.75) | 0.65 (0.59-0.68) | 0.70 (0.63-0.77) | 0.47 (0.40-0.54) | 0.53 (0.46-0.60) | 0.30 (0.23-0.37) | 0.54 (0.47-0.60) | 0.30 (0.24-0.37) | 0.70 (0.63-0.76) | 0.46 (0.40-0.53) |

[1]number of unique studies. If multiple cohorts were described in a study, those were meta-analyzed. This is how meta-analyzed results can be shown for a single study.

[2]estimates were not meta-analyzed if pAUC or gAUC are listed as NA, due to only a single cohort from a single study.

#### Supplementary Table 7. Meta-analyzed estimates of all biomarkers for all cancer types

| Biomarker | Studies N[1] | N | pAUC[2] | gAUC[2] | Sensitivity | False positive rate | Specificity | False negative rate | Positive predictive value | False omission rate | Negative predictive value | False discovery rate |
|---|---|---|---|---|---|---|---|---|---|---|---|---|
| AEs | 1 | 1747 | #N/A | #N/A | 0.36 | 0.34 | 0.66 | 0.64 | 0.21 | 0.19 | 0.81 | 0.79 |
| GEP | 5 | 783 | 0.6 (0.42-0.83) | 0.56 (0.49-0.65) | 0.71 (0.55-0.84) | 0.52 (0.44-0.59) | 0.48 (0.41-0.56) | 0.29 (0.16-0.45) | 0.45 (0.30-0.61) | 0.27 (0.14-0.46) | 0.73 (0.54-0.86) | 0.55 (0.39-0.70) |
| IMDC | 2 | 547 | 0.68 (0.2-0.95) | 0.57 (0.32-0.83) | 0.72 (0.17-0.97) | 0.57 (0.17-0.89) | 0.43 (0.11-0.83) | 0.28 (0.03-0.83) | 0.59 (0.51-0.66) | 0.42 (0.21-0.66) | 0.58 (0.34-0.79) | 0.41 (0.34-0.49) |
| MB | 3 | 228 | 0.56 (0.16-0.77) | 0.81 (0.38-0.95) | 0.48 (0.19-0.78) | 0.10 (0.02-0.33) | 0.90 (0.67-0.98) | 0.52 (0.22-0.81) | 0.79 (0.53-0.92) | 0.38 (0.14-0.70) | 0.62 (0.30-0.86) | 0.21 (0.08-0.47) |
| mIHC/IF | 3 | 177 | 0.81 (0.34-0.91) | 0.81 (0.58-0.91) | 0.85 (0.68-0.94) | 0.34 (0.20-0.51) | 0.66 (0.49-0.80) | 0.15 (0.06-0.32) | 0.78 (0.63-0.89) | 0.24 (0.11-0.44) | 0.76 (0.56-0.89) | 0.22 (0.11-0.37) |
| Multimodal | 5 | 559 | 0.52 (0.45-0.69) | 0.7 (0.61-0.76) | 0.57 (0.47-0.67) | 0.24 (0.17-0.34) | 0.76 (0.66-0.83) | 0.43 (0.33-0.53) | 0.50 (0.41-0.59) | 0.19 (0.13-0.29) | 0.81 (0.71-0.87) | 0.50 (0.41-0.59) |
| Others | 10 | 1001 | 0.59 (0.52-0.69) | 0.67 (0.58-0.71) | 0.60 (0.50-0.69) | 0.31 (0.21-0.44) | 0.69 (0.56-0.79) | 0.40 (0.31-0.50) | 0.46 (0.37-0.55) | 0.19 (0.13-0.28) | 0.81 (0.72-0.87) | 0.54 (0.45-0.63) |
| PD-L1 IHC | 76 | 13909 | 0.53 (0.53-0.65) | 0.58 (0.56-0.6) | 0.64 (0.60-0.69) | 0.51 (0.47-0.56) | 0.49 (0.44-0.53) | 0.36 (0.31-0.40) | 0.40 (0.36-0.45) | 0.28 (0.24-0.33) | 0.72 (0.67-0.76) | 0.60 (0.55-0.64) |
| TMB | 15 | 1814 | 0.57 (0.52-0.75) | 0.65 (0.59-0.68) | 0.70 (0.63-0.77) | 0.47 (0.40-0.54) | 0.53 (0.46-0.60) | 0.30 (0.23-0.37) | 0.54 (0.47-0.60) | 0.30 (0.24-0.37) | 0.70 (0.63-0.76) | 0.46 (0.40-0.53) |

[1]number of unique studies. If multiple cohorts were described in a study, those were meta-analyzed. This is how meta-analyzed results can be shown for a single study.

[2]estimates were not meta-analyzed if pAUC or gAUC are listed as NA, due to only a single cohort from a single study.

#### Supplementary Table 8. Meta-analyzed estimates of targeted mutations for all cancer types

| Biomarker | Mutation Status | Cancer/s | Studies N[1] | N | pAUC[2] | gAUC[2] | Sensitivity | False positive rate | Specificity | False negative rate | Positive predictive value | False omission rate | Negative predictive value | False discovery rate |
|---|---|---|---|---|---|---|---|---|---|---|---|---|---|---|
| BRAF | Mutant | melanoma, colon | 7 | 1022 | 0.54 (0.34-0.62) | 0.48 (0.44-0.53) | 0.49 (0.35-0.63) | 0.51 (0.39-0.63) | 0.49 (0.37-0.61) | 0.51 (0.37-0.65) | 0.42 (0.35-0.50) | 0.47 (0.37-0.57) | 0.53 (0.43-0.63) | 0.58 (0.50-0.65) |
| BRAF | Wildtype | melanoma, colon | 7 | 1022 | 0.45 (0.35-0.64) | 0.51 (0.46-0.55) | 0.51 (0.37-0.65) | 0.49 (0.37-0.61) | 0.51 (0.39-0.63) | 0.49 (0.35-0.63) | 0.47 (0.37-0.57) | 0.42 (0.35-0.50) | 0.58 (0.50-0.65) | 0.53 (0.43-0.63) |
| ER,HER2+/- or PR | Mutant | Breast | 1 | 156 | 0.29 (0.14-0.62) | 0.39 (0.2-0.61) | 0.28 (0.08-0.65) | 0.41 (0.17-0.70) | 0.59 (0.30-0.83) | 0.72 (0.35-0.92) | 0.03 (0.01-0.06) | 0.05 (0.03-0.08) | 0.95 (0.92-0.97) | 0.97 (0.94-0.99) |
| ER,HER2+/- or PR | Wildtype | Breast | 1 | 156 | 0.7 (0.4-0.86) | 0.6 (0.34-0.79) | 0.72 (0.35-0.92) | 0.59 (0.30-0.83) | 0.41 (0.17-0.70) | 0.28 (0.08-0.65) | 0.05 (0.03-0.08) | 0.03 (0.01-0.06) | 0.97 (0.94-0.99) | 0.95 (0.92-0.97) |
| PBRM1 | Mutant | renal cell carcinoma | 1 | 71 | 0.45 (0.3-0.65) | 0.55 (0.32-0.75) | 0.46 (0.29-0.64) | 0.38 (0.22-0.56) | 0.62 (0.44-0.78) | 0.54 (0.36-0.71) | 0.54 (0.14-0.89) | 0.45 (0.27-0.63) | 0.55 (0.37-0.73) | 0.46 (0.11-0.86) |
| PBRM1 | Wildtype | renal cell carcinoma | 1 | 71 | 0.54 (0.36-0.7) | 0.44 (0.24-0.64) | 0.54 (0.36-0.71) | 0.62 (0.44-0.78) | 0.38 (0.22-0.56) | 0.46 (0.29-0.64) | 0.45 (0.27-0.63) | 0.54 (0.14-0.89) | 0.46 (0.11-0.86) | 0.55 (0.37-0.73) |
| RAS | Mutant | melanoma, colon | 3 | 286 | 0.25 (0.15-0.43) | 0.29 (0.19-0.59) | 0.26 (0.15-0.42) | 0.40 (0.23-0.60) | 0.60 (0.40-0.77) | 0.74 (0.58-0.85) | 0.10 (0.01-0.54) | 0.16 (0.04-0.49) | 0.84 (0.51-0.96) | 0.90 (0.46-0.99) |
| RAS | Wildtype | melanoma, colon | 3 | 286 | 0.74 (0.52-0.82) | 0.7 (0.37-0.8) | 0.74 (0.58-0.85) | 0.60 (0.40-0.77) | 0.40 (0.23-0.60) | 0.26 (0.15-0.42) | 0.16 (0.04-0.49) | 0.10 (0.01-0.54) | 0.90 (0.46-0.99) | 0.84 (0.51-0.96) |
| VHL | Mutant | renal cell carcinoma | 1 | 71 | 0.59 (0.42-0.75) | 0.41 (0.33-0.7) | 0.61 (0.43-0.76) | 0.59 (0.47-0.70) | 0.41 (0.30-0.53) | 0.39 (0.24-0.57) | 0.49 (0.26-0.73) | 0.46 (0.14-0.81) | 0.54 (0.19-0.86) | 0.51 (0.27-0.74) |
| VHL | Wildtype | renal cell carcinoma | 1 | 71 | 0.4 (0.23-0.56) | 0.58 (0.29-0.67) | 0.39 (0.24-0.57) | 0.41 (0.30-0.53) | 0.59 (0.47-0.70) | 0.61 (0.43-0.76) | 0.46 (0.14-0.81) | 0.49 (0.26-0.73) | 0.51 (0.27-0.74) | 0.54 (0.19-0.86) |
| BRCA2 | Wildtype | melanoma | 1 | 38 | NA | NA | 0.71 | 0.94 | 0.06 | 0.29 | 0.48 | 0.86 | 0.14 | 0.52 |
| BRCA2 | Mutant | melanoma | 1 | 38 | NA | NA | 0.29 | 0.06 | 0.94 | 0.71 | 0.86 | 0.48 | 0.52 | 0.14 |
| DDR | Wildtype | urothelial cancer | 1 | 60 | NA | NA | 0.09 | 0.5 | 0.5 | 0.91 | 0.18 | 0.67 | 0.33 | 0.82 |
| DDR | Mutant | urothelial cancer | 1 | 60 | NA | NA | 0.91 | 0.5 | 0.5 | 0.09 | 0.67 | 0.18 | 0.82 | 0.33 |
| TP53 | Mutant | multiple cancers | 1 | 42 | NA | NA | 0.45 | 0.32 | 0.68 | 0.55 | 0.56 | 0.42 | 0.58 | 0.44 |
| TP53 | Wildtype | multiple cancers | 1 | 42 | NA | NA | 0.55 | 0.68 | 0.32 | 0.45 | 0.42 | 0.56 | 0.44 | 0.58 |
| TP53 and KRAS | Wildtype | melanoma | 1 | 38 | NA | NA | 0.48 | 0.53 | 0.47 | 0.52 | 0.53 | 0.58 | 0.42 | 0.47 |
| TP53 and KRAS | Mutant | melanoma | 1 | 38 | NA | NA | 0.52 | 0.47 | 0.53 | 0.48 | 0.58 | 0.53 | 0.47 | 0.42 |

[1]number of unique studies. If multiple cohorts were described in a study, those were meta-analyzed. This is how meta-analyzed results can be shown for a single study.

[2]estimates were not meta-analyzed if pAUC or gAUC are listed as NA, due to only a single cohort from a single study.

#### Supplementary Table 9. Study-level data*

| Authors | Year | Cancer | Treatment | Biomarker | Maximum Balance Accuracy | N | Sensitivity | Specificity | Positive predictive value | Negative predictive value | False positive rate | False discovery rate | False omission rate | False negative rate |
|---|---|---|---|---|---|---|---|---|---|---|---|---|---|---|
| Adams et al | 2019 | Breast | anti-PD-1 therapy | PD-L1 IHC | 0.60 | 169 | 0.60 | 0.38 | 0.14 | 0.84 | 0.63 | 0.86 | 0.16 | 0.40 |
| Antonia et al | 2016 | Small-cell lung cancer | anti-PD-1 therapy | PD-L1 IHC | 0.79 | 113 | 0.79 | 0.16 | 0.20 | 0.74 | 0.84 | 0.80 | 0.26 | 0.21 |
| Armand et al | 2019 | B-Cell Lymphoma | anti-PD-1 therapy | PD-L1 IHC | 0.89 | 42 | 0.89 | 0.26 | 0.50 | 0.75 | 0.74 | 0.50 | 0.25 | 0.11 |
| Balar et al | 2017 | Urothelial cancer | anti-PD-1 therapy | PD-L1 IHC | 0.72 | 119 | 0.72 | 0.35 | 0.33 | 0.74 | 0.65 | 0.68 | 0.26 | 0.28 |
| Baumi et al | 2017 | Head and neck | anti-PD-1 therapy | PD-L1 IHC | 0.85 | 166 | 0.85 | 0.16 | 0.20 | 0.81 | 0.84 | 0.80 | 0.19 | 0.15 |
| Bellmunt et al | 2017 | Urothelial cancer | anti-PD-1 therapy | PD-L1 IHC | 0.25 | 270 | 0.25 | 0.72 | 0.24 | 0.72 | 0.28 | 0.76 | 0.28 | 0.75 |
| Bonta et al | 2017 | Multiple cancers | anti-PD-1 therapy+anti-CTLA4 | TMB | 0.94 | 30 | 0.94 | 0.58 | 0.77 | 0.88 | 0.42 | 0.23 | 0.13 | 0.06 |
| Borghaei et al | 2015 | Non-small-cell lung cancer | anti-PD-1 therapy | PD-L1 IHC | 0.69 | 231 | 0.69 | 0.53 | 0.38 | 0.81 | 0.47 | 0.62 | 0.19 | 0.31 |
| Campesato et al | 2015 | Non-small-cell lung cancer | anti-PD-1 therapy | TMB | 0.79 | 31 | 0.79 | 0.71 | 0.69 | 0.80 | 0.29 | 0.31 | 0.20 | 0.21 |
| Carbone DP et al | 2017 | Non-small-cell lung cancer | anti-PD-1 therapy | PD-L1 IHC | 0.55 | 158 | 0.55 | 0.72 | 0.46 | 0.79 | 0.28 | 0.54 | 0.21 | 0.45 |
| Carbone DP et al | 2017 | Non-small-cell lung cancer | anti-PD-1 therapy | TMB | 0.46 | 158 | 0.46 | 0.79 | 0.55 | 0.73 | 0.21 | 0.45 | 0.27 | 0.54 |
| Carbone DP et al | 2017 | Non-small-cell lung cancer | anti-PD-1 therapy | Multimodal | 0.26 | 158 | 0.26 | 0.96 | 0.75 | 0.75 | 0.04 | 0.25 | 0.25 | 0.74 |
| Chow et al | 2016 | Head and neck | anti-PD-1 therapy | PD-L1 IHC | 0.71 | 132 | 0.71 | 0.33 | 0.19 | 0.84 | 0.67 | 0.81 | 0.16 | 0.29 |
| Chow et al | 2016 | Head and neck | anti-PD-1 therapy | PD-L1 IHC | 0.96 | 132 | 0.96 | 0.22 | 0.21 | 0.96 | 0.78 | 0.79 | 0.04 | 0.04 |
| Chung et al | 2018 | Small-cell lung cancer | anti-PD-1 therapy | PD-L1 IHC | 0.83 | 92 | 0.83 | 0.64 | 0.36 | 0.94 | 0.36 | 0.64 | 0.06 | 0.17 |
| Chung et al | 2019 | Cervical cancer | anti-PD-1 therapy | PD-L1 IHC | 0.80 | 84 | 0.80 | 0.14 | 0.17 | 0.77 | 0.86 | 0.83 | 0.23 | 0.20 |
| Cohen et al | 2019 | Head and neck | anti-PD-1 therapy | PD-L1 IHC | 0.88 | 199 | 0.88 | 0.29 | 0.51 | 0.74 | 0.71 | 0.49 | 0.26 | 0.12 |
| Cristescu et al | 2018 | HNSCC | anti-PD-1 therapy | GEP | 0.95 | 105 | 0.95 | 0.35 | 0.27 | 0.97 | 0.65 | 0.73 | 0.03 | 0.05 |
| Cristescu et al | 2018 | HNSCC | anti-PD-1 therapy | PD-L1 IHC | 0.90 | 107 | 0.90 | 0.19 | 0.21 | 0.89 | 0.81 | 0.79 | 0.11 | 0.10 |
| Cristescu et al | 2018 | HNSCC | anti-PD-1 therapy | TMB | 0.71 | 105 | 0.71 | 0.55 | 0.28 | 0.88 | 0.45 | 0.72 | 0.12 | 0.29 |
| Cristescu et al | 2018 | HNSCC | anti-PD-1 therapy | Multimodal | 0.67 | 105 | 0.67 | 0.71 | 0.37 | 0.90 | 0.29 | 0.63 | 0.10 | 0.33 |
| Cristescu et al | 2018 | HNSCC | anti-PD-1 therapy | Multimodal | 0.67 | 107 | 0.67 | 0.62 | 0.30 | 0.88 | 0.38 | 0.70 | 0.12 | 0.33 |
| Cristescu et al | 2018 | Melanoma | anti-PD-1 therapy | GEP | 0.84 | 86 | 0.84 | 0.35 | 0.51 | 0.74 | 0.65 | 0.49 | 0.26 | 0.16 |
| Cristescu et al | 2018 | Melanoma | anti-PD-1 therapy | PD-L1 IHC | 0.92 | 89 | 0.92 | 0.16 | 0.45 | 0.73 | 0.84 | 0.55 | 0.27 | 0.08 |
| Cristescu et al | 2018 | Melanoma | anti-PD-1 therapy | TMB | 0.89 | 86 | 0.89 | 0.31 | 0.51 | 0.79 | 0.69 | 0.49 | 0.21 | 0.11 |
| Cristescu et al | 2018 | Melanoma | anti-PD-1 therapy | Multimodal | 0.68 | 86 | 0.68 | 0.58 | 0.57 | 0.70 | 0.42 | 0.43 | 0.30 | 0.32 |

| | | | | | | | | | | | | | | |
|---|---|---|---|---|---|---|---|---|---|---|---|---|---|---|
| Cristescu et al | 2018 | Melanoma | anti-PD-1 therapy | Multimodal | 0.74 | 89 | 0.74 | 0.59 | 0.57 | 0.75 | 0.41 | 0.43 | 0.25 | 0.26 |
| Cristescu et al | 2018 | Multiple cancers | anti-PD-1 therapy | GEP | 0.94 | 113 | 0.94 | 0.45 | 0.22 | 0.98 | 0.55 | 0.78 | 0.02 | 0.06 |
| Cristescu et al | 2018 | Multiple cancers | anti-PD-1 therapy | PD-L1 IHC | 1.00 | 74 | 1.00 | 0.48 | 0.21 | 1.00 | 0.52 | 0.79 | 0.00 | 0.00 |
| Cristescu et al | 2018 | Multiple cancers | anti-PD-1 therapy | TMB | 0.94 | 113 | 0.94 | 0.45 | 0.22 | 0.98 | 0.55 | 0.78 | 0.02 | 0.06 |
| Cristescu et al | 2018 | Multiple cancers | anti-PD-1 therapy | Multimodal | 0.63 | 113 | 0.63 | 0.82 | 0.37 | 0.93 | 0.18 | 0.63 | 0.07 | 0.38 |
| Cristescu et al | 2018 | Multiple cancers | anti-PD-1 therapy | Multimodal | 0.78 | 74 | 0.78 | 0.80 | 0.35 | 0.96 | 0.20 | 0.65 | 0.04 | 0.22 |
| Daud et al | 2014 | Melanoma | anti-PD-1 therapy | PD-L1 IHC | 0.86 | 71 | 0.86 | 0.32 | 0.58 | 0.69 | 0.68 | 0.42 | 0.31 | 0.14 |
| Daud et al | 2016 | Melanoma | anti-PD-1 therapy | PD-L1 IHC | 0.88 | 451 | 0.88 | 0.31 | 0.45 | 0.79 | 0.69 | 0.55 | 0.21 | 0.13 |
| Dirix et al | 2018 | Breast | anti-PD-1 therapy | PD-L1 IHC | 0.25 | 136 | 0.25 | 0.93 | 0.25 | 0.93 | 0.07 | 0.75 | 0.07 | 0.75 |
| El-Khoueiry et al | 2017 | HCC | anti-PD-1 therapy | PD-L1 IHC | 0.12 | 41 | 0.12 | 0.53 | 0.30 | 0.26 | 0.47 | 0.70 | 0.74 | 0.88 |
| El-Khoueiry et al | 2017 | HCC | anti-PD-1 therapy | PD-L1 IHC | 0.22 | 168 | 0.22 | 0.84 | 0.74 | 0.34 | 0.16 | 0.26 | 0.66 | 0.78 |
| Eng et al | 2019 | Colon | anti-PD-1 therapy | PD-L1 IHC | 0.50 | 90 | 0.50 | 0.61 | 0.03 | 0.98 | 0.39 | 0.97 | 0.02 | 0.50 |
| Eng et al | 2019 | Colon | anti-PD-1 therapy | Others | 0.50 | 90 | 0.50 | 0.98 | 0.33 | 0.99 | 0.02 | 0.67 | 0.01 | 0.50 |
| Eng et al | 2019 | Colon | anti-PD-1 therapy+MEK | PD-L1 IHC | 0.60 | 183 | 0.60 | 0.57 | 0.04 | 0.98 | 0.43 | 0.96 | 0.02 | 0.40 |
| Eng et al | 2019 | Colon | anti-PD-1 therapy+MEK | Others | 0.40 | 183 | 0.40 | 0.99 | 0.67 | 0.98 | 0.01 | 0.33 | 0.02 | 0.60 |
| Fehrenbacher et al | 2016 | Non-small-cell lung cancer | anti-PD-1 therapy | PD-L1 IHC | 0.81 | 144 | 0.81 | 0.38 | 0.18 | 0.92 | 0.62 | 0.82 | 0.08 | 0.19 |
| Ferris et al | 2016 | Head and neck | anti-PD-1 therapy | PD-L1 IHC | 0.63 | 161 | 0.63 | 0.47 | 0.17 | 0.88 | 0.53 | 0.83 | 0.12 | 0.38 |
| Fuchs et al | 2018 | Gastric and gastroesophageal junction | anti-PD-1 therapy | PD-L1 IHC | 0.68 | 216 | 0.68 | 0.45 | 0.38 | 0.74 | 0.55 | 0.62 | 0.26 | 0.32 |
| Gandara et al | 2018 | Non-small-cell lung cancer | anti-PD-1 therapy | TMB | 0.26 | 583 | 0.26 | 0.72 | 0.49 | 0.48 | 0.28 | 0.51 | 0.52 | 0.74 |
| Gandara et al | 2018 | 8Non-small-cell lung cancer | anti-PD-1 therapy | PD-L1 IHC | 0.58 | 578 | 0.58 | 0.45 | 0.52 | 0.51 | 0.55 | 0.48 | 0.49 | 0.42 |
| Gandhi et al | 2014 | Non-small-cell lung cancer | anti-PD-1 therapy | PD-L1 IHC | 0.73 | 29 | 0.73 | 0.88 | 0.69 | 0.90 | 0.12 | 0.31 | 0.10 | 0.27 |
| Gandhi et al | 2018 | Non-small-cell lung cancer | anti-PD-1 therapy | PD-L1 IHC | 0.75 | 387 | 0.75 | 0.45 | 0.69 | 0.53 | 0.55 | 0.31 | 0.47 | 0.25 |
| Garon et al | 2015 | Non-small-cell lung cancer | anti-PD-1 therapy | PD-L1 IHC | 0.81 | 121 | 0.81 | 0.36 | 0.21 | 0.90 | 0.64 | 0.79 | 0.10 | 0.19 |
| Garon et al | 2015 | Non-small-cell lung cancer | anti-PD-1 therapy | PD-L1 IHC | 0.89 | 356 | 0.89 | 0.29 | 0.49 | 0.78 | 0.71 | 0.51 | 0.22 | 0.11 |
| Garon et al | 2015 | Non-small-cell lung cancer | anti-PD-1 therapy | PD-L1 IHC | 0.94 | 204 | 0.94 | 0.17 | 0.28 | 0.89 | 0.83 | 0.72 | 0.11 | 0.06 |
| Gettinger et al | 2014 | Non-small-cell lung cancer | anti-PD-1 therapy | PD-L1 IHC | 1.00 | 15 | 1.00 | 0.67 | 0.67 | 1.00 | 0.33 | 0.33 | 0.00 | 0.00 |
| Giraldo et al | 2018 | Merkel cell | anti-PD-1 therapy | Others | 0.81 | 23 | 0.81 | 0.43 | 0.76 | 0.50 | 0.57 | 0.24 | 0.50 | 0.19 |
| Giraldo et al | 2018 | Merkel cell | anti-PD-1 therapy | mIHC/IF | 0.93 | 20 | 0.93 | 0.67 | 0.87 | 0.80 | 0.33 | 0.13 | 0.20 | 0.07 |
| Goodman et al | 2017 | Melanoma | anti-PD-1 therapy | TMB | 1.00 | 11 | 1.00 | 0.67 | 0.89 | 1.00 | 0.33 | 0.11 | 0.00 | 0.00 |

| | | | | | | | | | | | | | | |
|---|---|---|---|---|---|---|---|---|---|---|---|---|---|---|
| Goodman et al | 2017 | Multiple cancers | anti-PD-1 therapy | TMB | 0.79 | 47 | 0.79 | 0.74 | 0.76 | 0.77 | 0.26 | 0.24 | 0.23 | 0.21 |
| Goodman et al | 2017 | Multiple cancers | anti-PD-1 therapy+anti-CTLA4 | TMB | 0.68 | 88 | 0.68 | 0.68 | 0.80 | 0.54 | 0.32 | 0.20 | 0.46 | 0.32 |
| Goodman et al | 2017 | Non-small-cell lung cancer | anti-PD-1 therapy | TMB | 0.69 | 36 | 0.69 | 0.75 | 0.69 | 0.75 | 0.25 | 0.31 | 0.25 | 0.31 |
| Gopalakrishnan | 2018 | Melanoma | anti-PD-1 therapy | MB | 0.37 | 43 | 0.37 | 1.00 | 1.00 | 0.41 | 0.00 | 0.00 | 0.59 | 0.63 |
| Gopalakrishnan | 2018 | Melanoma | anti-PD-1 therapy | MB | 0.40 | 86 | 0.40 | 0.68 | 0.66 | 0.43 | 0.32 | 0.34 | 0.57 | 0.60 |
| Grosso et al | 2013 | Melanoma | anti-PD-1 therapy | PD-L1 IHC | 0.70 | 34 | 0.70 | 0.63 | 0.44 | 0.83 | 0.38 | 0.56 | 0.17 | 0.30 |
| Hellmann et al. | 2018 | Non-small-cell lung cancer | anti-PD-1 therapy+anti-CTLA4 | PD-L1 IHC | 0.82 | 70 | 0.82 | 0.48 | 0.51 | 0.80 | 0.52 | 0.49 | 0.20 | 0.18 |
| Hellmann et al. | 2018 | Non-small-cell lung cancer | anti-PD-1 therapy+anti-CTLA4 | Multimodal | 0.65 | 70 | 0.65 | 0.81 | 0.63 | 0.83 | 0.19 | 0.38 | 0.17 | 0.35 |
| Hellmann et al. | 2018 | Non-small-cell lung cancer | anti-PD-1 therapy+anti-CTLA4 | TMB | 0.65 | 75 | 0.65 | 0.66 | 0.65 | 0.66 | 0.34 | 0.35 | 0.34 | 0.35 |
| Hellmann et al. | 2018 | Non-small-cell lung cancer | anti-PD-1 therapy+anti-CTLA4 | PD-L1 IHC | 0.76 | 139 | 0.76 | 0.30 | 0.50 | 0.58 | 0.70 | 0.50 | 0.42 | 0.24 |
| Hellmann et al. | 2018 | Small-cell lung cancer | anti-PD-1 therapy | PD-L1 IHC | 0.10 | 75 | 0.10 | 0.85 | 0.09 | 0.86 | 0.15 | 0.91 | 0.14 | 0.90 |
| Hellmann et al. | 2018 | Small-cell lung cancer | anti-PD-1 therapy | TMB | 0.87 | 133 | 0.87 | 0.34 | 0.14 | 0.95 | 0.66 | 0.86 | 0.05 | 0.13 |
| Hellmann et al. | 2018 | Small-cell lung cancer | anti-PD-1 therapy+anti-CTLA4 | TMB | 0.75 | 78 | 0.75 | 0.38 | 0.29 | 0.81 | 0.62 | 0.71 | 0.19 | 0.25 |
| Helmann et al | 2019 | Non-small-cell lung cancer | anti-PD-1 therapy + chemo | PD-L1 IHC | 0.62 | 508 | 0.62 | 0.15 | 0.69 | 0.12 | 0.85 | 0.31 | 0.88 | 0.38 |
| Helmann et al | 2019 | Non-small-cell lung cancer | anti-PD-1 therapy+anti-CTLA4 | PD-L1 IHC | 0.65 | 573 | 0.65 | 0.29 | 0.34 | 0.59 | 0.71 | 0.66 | 0.41 | 0.35 |
| Herbst et al | 2014 | Multiple cancers | anti-PD-1 therapy | PD-L1 IHC | 0.69 | 150 | 0.69 | 0.47 | 0.49 | 0.67 | 0.53 | 0.51 | 0.33 | 0.31 |
| Herbst et al | 2014 | Non-small-cell lung cancer | anti-PD-1 therapy | PD-L1 IHC | 0.50 | 46 | 0.50 | 0.39 | 0.35 | 0.55 | 0.61 | 0.65 | 0.45 | 0.50 |
| Herbst et al | 2016 | Non-small-cell lung cancer | anti-PD-1 therapy | PD-L1 IHC | 0.68 | 690 | 0.68 | 0.64 | 0.30 | 0.90 | 0.36 | 0.70 | 0.10 | 0.32 |
| Horn et al | 2017 | Non-small-cell lung cancer | anti-PD-1 therapy | PD-L1 IHC | 0.53 | 104 | 0.53 | 0.49 | 0.54 | 0.48 | 0.51 | 0.46 | 0.52 | 0.47 |
| Hugo et al. | 2016 | Melanoma | anti-PD-1 therapy | GEP | 0.93 | 27 | 0.93 | 0.69 | 0.76 | 0.90 | 0.31 | 0.24 | 0.10 | 0.07 |
| Janjigian et al | 2018 | Gastric and gastroesophageal junction | anti-PD-1 therapy | PD-L1 IHC | 0.50 | 36 | 0.50 | 0.67 | 0.23 | 0.87 | 0.33 | 0.77 | 0.13 | 0.50 |
| Janjigian et al | 2018 | Gastric and gastroesophageal junction | anti-PD-1 therapy | Others | 0.50 | 25 | 0.50 | 0.76 | 0.29 | 0.89 | 0.24 | 0.71 | 0.11 | 0.50 |
| Janjigian et al | 2018 | Gastric and gastroesophageal junction | anti-PD-1 therapy+anti-CTLA4 | PD-L1 IHC | 0.50 | 74 | 0.50 | 0.75 | 0.32 | 0.87 | 0.25 | 0.68 | 0.13 | 0.50 |
| Janjigian et al | 2018 | Gastric and gastroesophageal junction | anti-PD-1 therapy+anti-CTLA4 | Others | 0.20 | 23 | 0.20 | 0.94 | 0.50 | 0.81 | 0.06 | 0.50 | 0.19 | 0.80 |
| Johnson et al | 2018 | Melanoma | anti-PD-1 therapy | Others | 0.85 | 24 | 0.85 | 0.91 | 0.92 | 0.83 | 0.09 | 0.08 | 0.17 | 0.15 |

| | | | | | | | | | | | | | | |
|---|---|---|---|---|---|---|---|---|---|---|---|---|---|---|
| Johnson et al | 2018 | Melanoma | anti-PD-1 therapy | mIHC/IF | 0.92 | 24 | 0.92 | 0.82 | 0.86 | 0.90 | 0.18 | 0.14 | 0.10 | 0.08 |
| Johnson et al | 2018 | Melanoma | anti-PD-1 therapy | Multimodal | 0.69 | 24 | 0.69 | 0.82 | 0.82 | 0.69 | 0.18 | 0.18 | 0.31 | 0.31 |
| Johnson et al | 2018 | Melanoma | anti-PD-1 therapy | Others | 0.60 | 142 | 0.60 | 0.72 | 0.72 | 0.60 | 0.28 | 0.28 | 0.40 | 0.40 |
| Johnson et al | 2018 | Melanoma | anti-PD-1 therapy | mIHC/IF | 0.71 | 142 | 0.71 | 0.52 | 0.64 | 0.59 | 0.48 | 0.36 | 0.41 | 0.29 |
| Johnson et al | 2018 | Melanoma | anti-PD-1 therapy | Multimodal | 0.51 | 142 | 0.51 | 0.66 | 0.65 | 0.53 | 0.34 | 0.35 | 0.48 | 0.49 |
| Kaufman et al | 2016 | Merkel cell | anti-PD-1 therapy | PD-L1 IHC | 0.87 | 74 | 0.87 | 0.25 | 0.34 | 0.81 | 0.75 | 0.66 | 0.19 | 0.13 |
| Kaufman et al | 2018 | Merkel cell | anti-PD-1 therapy | PD-L1 IHC | 0.88 | 74 | 0.88 | 0.26 | 0.36 | 0.81 | 0.74 | 0.64 | 0.19 | 0.13 |
| Kefford et al | 2014 | Melanoma | anti-PD-1 therapy | PD-L1 IHC | 0.97 | 71 | 0.97 | 0.36 | 0.51 | 0.94 | 0.64 | 0.49 | 0.06 | 0.03 |
| Khagi et al | 2017 | Multiple cancers | anti-PD-1 therapy | Others | 0.56 | 66 | 0.56 | 0.78 | 0.45 | 0.85 | 0.22 | 0.55 | 0.15 | 0.44 |
| Langer et al | 2016 | Non-small-cell lung cancer | anti-PD-1 therapy + chemo | PD-L1 IHC | 0.64 | 60 | 0.64 | 0.33 | 0.54 | 0.43 | 0.67 | 0.46 | 0.57 | 0.36 |
| Larkin et al | 2018 | Melanoma | anti-PD-1 therapy | PD-L1 IHC | 0.87 | 248 | 0.87 | 0.34 | 0.34 | 0.87 | 0.66 | 0.66 | 0.13 | 0.13 |
| Larkin et al | 2015 | Melanoma | anti-PD-1 therapy | PD-L1 IHC | 0.35 | 288 | 0.35 | 0.78 | 0.58 | 0.59 | 0.22 | 0.43 | 0.41 | 0.65 |
| Larkin et al | 2015 | Melanoma | anti-PD-1 therapy+anti-CTLA4 | PD-L1 IHC | 0.30 | 278 | 0.30 | 0.83 | 0.72 | 0.45 | 0.17 | 0.28 | 0.55 | 0.70 |
| Loo et al | 2017 | Head and neck | anti-PD-1 therapy | Others | 0.97 | 64 | 0.97 | 0.53 | 0.67 | 0.94 | 0.47 | 0.33 | 0.06 | 0.03 |
| Loo et al | 2017 | Head and neck | anti-PD-1 therapy+anti-CTLA4 | Others | 0.46 | 29 | 0.46 | 0.81 | 0.67 | 0.65 | 0.19 | 0.33 | 0.35 | 0.54 |
| Maher et al | 2019 | Urothelial cancer | anti-PD-1 therapy | AEs | 0.36 | 1747 | 0.36 | 0.66 | 0.21 | 0.81 | 0.34 | 0.79 | 0.19 | 0.64 |
| Matson et al | 2018 | Melanoma | anti-PD-1 therapy | MB | 0.94 | 42 | 0.94 | 1.00 | 1.00 | 0.96 | 0.00 | 0.00 | 0.04 | 0.06 |
| McDermott et al | 2018 | Renal cell carcinoma | anti-PD-1 therapy | TMB | 0.71 | 71 | 0.71 | 0.53 | 0.44 | 0.78 | 0.47 | 0.56 | 0.22 | 0.29 |
| McDermott et al | 2018 | Renal cell carcinoma | anti-PD-1 therapy | TMB | 0.79 | 71 | 0.79 | 0.21 | 0.34 | 0.67 | 0.79 | 0.66 | 0.33 | 0.21 |
| McDermott et al | 2018 | Renal cell carcinoma | anti-PD-1 therapy | GEP | 0.56 | 86 | 0.56 | 0.48 | 0.41 | 0.63 | 0.52 | 0.59 | 0.38 | 0.44 |
| McDermott et al | 2018 | Renal cell carcinoma | anti-PD-1 therapy | Others | 0.71 | 71 | 0.71 | 0.36 | 0.36 | 0.71 | 0.64 | 0.64 | 0.29 | 0.29 |
| McDermott et al | 2018 | Renal cell carcinoma | anti-PD-1 therapy | TMB | 0.79 | 71 | 0.79 | 0.21 | 0.34 | 0.67 | 0.79 | 0.66 | 0.33 | 0.21 |
| McDermott et al | 2018 | Renal cell carcinoma | anti-PD-1 therapy+bev | TMB | 0.44 | 65 | 0.44 | 0.46 | 0.58 | 0.32 | 0.54 | 0.42 | 0.68 | 0.56 |
| McDermott et al | 2018 | Renal cell carcinoma | anti-PD-1 therapy+bev | TMB | 0.66 | 65 | 0.66 | 0.42 | 0.66 | 0.42 | 0.58 | 0.34 | 0.58 | 0.34 |
| McDermott et al | 2018 | Renal cell carcinoma | anti-PD-1 therapy+bev | TMB | 0.80 | 65 | 0.80 | 0.25 | 0.65 | 0.43 | 0.75 | 0.35 | 0.57 | 0.20 |
| McDermott et al | 2018 | Renal cell carcinoma | anti-PD-1 therapy+bev | GEP | 0.59 | 88 | 0.59 | 0.65 | 0.70 | 0.53 | 0.35 | 0.30 | 0.47 | 0.41 |
| McDermott et al | 2018 | Renal cell carcinoma | anti-PD-1 therapy+bev | Others | 0.49 | 65 | 0.49 | 0.38 | 0.57 | 0.30 | 0.63 | 0.43 | 0.70 | 0.51 |
| Mehra et al | 2018 | Head and neck | anti-PD-1 therapy | PD-L1 IHC | 0.65 | 188 | 0.65 | 0.34 | 0.18 | 0.82 | 0.66 | 0.82 | 0.18 | 0.35 |
| Mehra et al | 2018 | Head and neck | anti-PD-1 therapy | Others | 0.81 | 172 | 0.81 | 0.39 | 0.23 | 0.90 | 0.61 | 0.77 | 0.10 | 0.19 |

| | | | | | | | | | | | | | | |
|---|---|---|---|---|---|---|---|---|---|---|---|---|---|---|
| Mehra et al | 2018 | Head and neck | anti-PD-1 therapy | PD-L1 IHC | 0.94 | 188 | 0.94 | 0.22 | 0.21 | 0.94 | 0.78 | 0.79 | 0.06 | 0.06 |
| Mok et al | 2019 | Non-small-cell lung cancer | anti-PD-1 therapy | PD-L1 IHC | 0.72 | 637 | 0.72 | 0.41 | 0.51 | 0.63 | 0.59 | 0.49 | 0.37 | 0.28 |
| Motzer et al | 2018 | Renal cell carcinoma | anti-PD-1 therapy | PD-L1 IHC | 0.51 | 184 | 0.51 | 0.59 | 0.85 | 0.22 | 0.41 | 0.15 | 0.78 | 0.49 |
| Motzer et al | 2019 | Renal cell carcinoma | anti-PD-1 therapy | PD-L1 IHC | 0.61 | 442 | 0.61 | 0.40 | 0.61 | 0.40 | 0.60 | 0.39 | 0.60 | 0.39 |
| Motzer et al | 2019 | Renal cell carcinoma | anti-PD-1 therapy | IMDC | 0.90 | 437 | 0.90 | 0.24 | 0.56 | 0.69 | 0.76 | 0.44 | 0.31 | 0.10 |
| Muro et al | 2016 | Gastric and gastroesophageal junction | anti-PD-1 therapy | PD-L1 IHC | 0.13 | 35 | 0.13 | 0.93 | 0.33 | 0.78 | 0.07 | 0.67 | 0.22 | 0.88 |
| Nghiem et al | 2019 | Merkel cell | anti-PD-1 therapy | PD-L1 IHC | 0.89 | 46 | 0.89 | 0.16 | 0.60 | 0.50 | 0.84 | 0.40 | 0.50 | 0.11 |
| Nghiem et al | 2019 | Merkel cell | anti-PD-1 therapy | PD-L1 IHC | 0.52 | 46 | 0.52 | 0.53 | 0.61 | 0.43 | 0.47 | 0.39 | 0.57 | 0.48 |
| Nghiem et al | 2019 | Merkel cell | anti-PD-1 therapy | PD-L1 IHC | 0.89 | 46 | 0.89 | 0.16 | 0.60 | 0.50 | 0.84 | 0.40 | 0.50 | 0.11 |
| Overman et al | 2017 | Colon | anti-PD-1 therapy | PD-L1 IHC | 0.74 | 68 | 0.74 | 0.39 | 0.32 | 0.79 | 0.61 | 0.68 | 0.21 | 0.26 |
| Overman et al | 2017 | Colon | anti-PD-1 therapy | PD-L1 IHC | 0.32 | 68 | 0.32 | 0.69 | 0.29 | 0.72 | 0.31 | 0.71 | 0.28 | 0.68 |
| Overman et al | 2018 | Colon | anti-PD-1 therapy | PD-L1 IHC | 0.29 | 91 | 0.29 | 0.72 | 0.54 | 0.48 | 0.28 | 0.46 | 0.52 | 0.71 |
| Patel et al | 2018 | Urothelial cancer | anti-PD-1 therapy | PD-L1 IHC | 0.57 | 114 | 0.57 | 0.68 | 0.65 | 0.60 | 0.32 | 0.35 | 0.40 | 0.43 |
| Paz_Ares et al | 2018 | Non-small-cell lung cancer | anti-PD-1 therapy + chemo | PD-L1 IHC | 0.63 | 271 | 0.63 | 0.33 | 0.53 | 0.42 | 0.67 | 0.47 | 0.58 | 0.37 |
| Plimack et al | 2017 | Urothelial cancer | anti-PD-1 therapy | PD-L1 IHC | 0.60 | 25 | 0.60 | 0.60 | 0.27 | 0.86 | 0.40 | 0.73 | 0.14 | 0.40 |
| Postow et al | 2015 | Melanoma | anti-PD-1 therapy+anti-CTLA4 | PD-L1 IHC | 0.31 | 80 | 0.31 | 0.71 | 0.58 | 0.45 | 0.29 | 0.42 | 0.55 | 0.69 |
| Powles et al | 2014 | Urothelial cancer | anti-PD-1 therapy | PD-L1 IHC | 0.84 | 59 | 0.84 | 0.24 | 0.67 | 0.45 | 0.76 | 0.33 | 0.55 | 0.16 |
| Powles et al | 2017 | Urothelial cancer | anti-PD-1 therapy | PD-L1 IHC | 0.88 | 144 | 0.88 | 0.43 | 0.17 | 0.96 | 0.57 | 0.83 | 0.04 | 0.12 |
| Puzanov et al | 2015 | Melanoma | anti-PD-1 therapy | PD-L1 IHC | 0.53 | 286 | 0.53 | 0.17 | 0.40 | 0.26 | 0.83 | 0.60 | 0.74 | 0.47 |
| Ready et al | 2019 | Non-small-cell lung cancer | anti-PD-1 therapy | PD-L1 IHC | 0.67 | 252 | 0.67 | 0.52 | 0.43 | 0.75 | 0.48 | 0.57 | 0.25 | 0.33 |
| Ready et al | 2019 | Non-small-cell lung cancer | anti-PD-1 therapy | TMB | 0.69 | 98 | 0.69 | 0.63 | 0.52 | 0.78 | 0.37 | 0.48 | 0.22 | 0.31 |
| Riaz et al | 2017 | Melanoma | anti-PD-1 therapy | TMB | 0.61 | 65 | 0.61 | 0.37 | 0.58 | 0.40 | 0.63 | 0.43 | 0.60 | 0.39 |
| Rini et al | 2019 | Renal cell carcinoma | anti-PD-1 therapy | PD-L1 IHC | 0.38 | 424 | 0.38 | 0.58 | 0.80 | 0.18 | 0.43 | 0.20 | 0.82 | 0.62 |
| Rittmeyer et al | 2017 | Non-small-cell lung cancer | anti-PD-1 therapy | PD-L1 IHC | 0.75 | 421 | 0.75 | 0.46 | 0.18 | 0.92 | 0.54 | 0.82 | 0.08 | 0.25 |
| Rizvi et al | 2015 | Non-small-cell lung cancer | anti-PD-1 therapy | TMB | 0.86 | 16 | 0.86 | 0.78 | 0.75 | 0.88 | 0.22 | 0.25 | 0.13 | 0.14 |
| Rizvi et al | 2015 | Non-small-cell lung cancer | anti-PD-1 therapy | TMB | 0.79 | 31 | 0.79 | 0.82 | 0.79 | 0.82 | 0.18 | 0.21 | 0.18 | 0.21 |
| Rizvi et al | 2015 | Non-small-cell lung cancer | anti-PD-1 therapy | TMB | 0.64 | 31 | 0.64 | 0.71 | 0.64 | 0.71 | 0.29 | 0.36 | 0.29 | 0.36 |
| Rizvi et al | 2015 | Non-small-cell lung cancer | anti-PD-1 therapy | TMB | 0.71 | 15 | 0.71 | 0.88 | 0.83 | 0.78 | 0.13 | 0.17 | 0.22 | 0.29 |
| Rizvi et al | 2018 | Non-small-cell lung cancer | anti-PD-1 therapy | PD-L1 IHC | 0.64 | 76 | 0.64 | 0.59 | 0.43 | 0.77 | 0.41 | 0.57 | 0.23 | 0.36 |

| | | | | | | | | | | | | | | |
|---|---|---|---|---|---|---|---|---|---|---|---|---|---|---|
| Rizvi et al | 2018 | Non-small-cell lung cancer | anti-PD-1 therapy | Multimodal | 0.40 | 76 | 0.40 | 0.80 | 0.50 | 0.73 | 0.20 | 0.50 | 0.27 | 0.60 |
| Rizvi et al | 2018 | Non-small-cell lung cancer | anti-PD-1 therapy | TMB | 0.64 | 240 | 0.64 | 0.56 | 0.37 | 0.79 | 0.44 | 0.63 | 0.21 | 0.36 |
| Robert et al | 2015 | Melanoma | anti-PD-1 therapy | PD-L1 IHC | 0.46 | 209 | 0.46 | 0.73 | 0.53 | 0.67 | 0.27 | 0.47 | 0.33 | 0.54 |
| Rosenberg et al | 2016 | Urothelial cancer | anti-PD-1 therapy | PD-L1 IHC | 0.69 | 260 | 0.69 | 0.33 | 0.58 | 0.43 | 0.67 | 0.42 | 0.57 | 0.31 |
| Rosenberg et al | 2016 | Urothelial cancer | anti-PD-1 therapy | PD-L1 IHC | 0.22 | 310 | 0.22 | 0.80 | 0.19 | 0.84 | 0.20 | 0.81 | 0.16 | 0.78 |
| Routy et al | 2018 | Multiple cancers | anti-PD-1 therapy | MB | 0.22 | 100 | 0.22 | 0.90 | 0.69 | 0.55 | 0.10 | 0.31 | 0.45 | 0.78 |
| Schmid et al | 2018 | Breast | anti-PD-1 therapy + chemo | PD-L1 IHC | 0.46 | 451 | 0.46 | 0.64 | 0.56 | 0.54 | 0.36 | 0.44 | 0.46 | 0.54 |
| Shah et al | 2019 | Multiple cancers | anti-PD-1 therapy | PD-L1 IHC | 0.68 | 121 | 0.68 | 0.56 | 0.22 | 0.90 | 0.44 | 0.78 | 0.10 | 0.32 |
| Sharma et al | 2017 | Head and neck | anti-PD-1 therapy | PD-L1 IHC | 0.56 | 265 | 0.56 | 0.56 | 0.24 | 0.84 | 0.44 | 0.76 | 0.16 | 0.44 |
| Sharma et al | 2017 | Multiple cancers | anti-PD-1 therapy | PD-L1 IHC | 0.44 | 265 | 0.44 | 0.73 | 0.28 | 0.84 | 0.27 | 0.72 | 0.16 | 0.56 |
| Singal et al | 2017 | Non-small-cell lung cancer | anti-PD-1 therapy+anti-CTLA4 | TMB | 0.23 | 33 | 0.23 | 1.00 | 1.00 | 0.39 | 0.00 | 0.00 | 0.61 | 0.77 |
| Socinski et al | 2018 | Non-small-cell lung cancer | anti-PD-1 therapy + chemo | PD-L1 IHC | 0.60 | 356 | 0.60 | 0.59 | 0.75 | 0.42 | 0.41 | 0.25 | 0.58 | 0.40 |
| Socinski et al | 2018 | Non-small-cell lung cancer | anti-PD-1 therapy + chemo | GEP | 0.49 | 341 | 0.49 | 0.61 | 0.72 | 0.38 | 0.39 | 0.28 | 0.62 | 0.51 |
| Soria et al | 2013 | Non-small-cell lung cancer | anti-PD-1 therapy | PD-L1 IHC | 0.27 | 30 | 0.27 | 1.00 | 1.00 | 0.58 | 0.00 | 0.00 | 0.42 | 0.73 |
| Soria et al | 2013 | Non-small-cell lung cancer | anti-PD-1 therapy | PD-L1 IHC | 0.50 | 30 | 0.50 | 1.00 | 1.00 | 0.85 | 0.00 | 0.00 | 0.15 | 0.50 |
| Taube et al | 2014 | Multiple cancers | anti-PD-1 therapy | Others | 0.83 | 41 | 0.83 | 0.55 | 0.43 | 0.89 | 0.45 | 0.57 | 0.11 | 0.17 |
| Taube et al | 2014 | Multiple cancers | anti-PD-1 therapy | PD-L1 IHC | 0.92 | 41 | 0.92 | 0.59 | 0.48 | 0.94 | 0.41 | 0.52 | 0.06 | 0.08 |
| Taube et al | 2014 | Multiple cancers | anti-PD-1 therapy | Others | 0.67 | 37 | 0.67 | 0.68 | 0.50 | 0.81 | 0.32 | 0.50 | 0.19 | 0.33 |
| Taube et al | 2014 | Multiple cancers | anti-PD-1 therapy | Others | 0.57 | 30 | 0.57 | 0.70 | 0.36 | 0.84 | 0.30 | 0.64 | 0.16 | 0.43 |
| Taube et al | 2014 | Multiple cancers | anti-PD-1 therapy | Others | 0.64 | 29 | 0.64 | 0.50 | 0.44 | 0.69 | 0.50 | 0.56 | 0.31 | 0.36 |
| Taube et al | 2014 | Multiple cancers | anti-PD-1 therapy | Others | 0.92 | 41 | 0.92 | 0.28 | 0.34 | 0.89 | 0.72 | 0.66 | 0.11 | 0.08 |
| Taube et al | 2014 | Multiple cancers | anti-PD-1 therapy | Others | 0.17 | 41 | 0.17 | 0.86 | 0.33 | 0.71 | 0.14 | 0.67 | 0.29 | 0.83 |
| Taube et al | 2014 | Multiple cancers | anti-PD-1 therapy | Others | 0.33 | 41 | 0.33 | 0.66 | 0.29 | 0.70 | 0.34 | 0.71 | 0.30 | 0.67 |
| Topalian et al | 2012 | Multiple cancers | anti-PD-1 therapy | PD-L1 IHC | 1.00 | 42 | 1.00 | 0.52 | 0.36 | 1.00 | 0.48 | 0.64 | 0.00 | 0.00 |
| Tumeh et al | 2014 | Melanoma | anti-PD-1 therapy | mIHC/IF | 1.00 | 15 | 1.00 | 0.80 | 0.91 | 1.00 | 0.20 | 0.09 | 0.00 | 0.00 |
| Tykodi et al | 2019 | Renal cell carcinoma | anti-PD-1 therapy | IMDC | 0.41 | 110 | 0.41 | 0.66 | 0.62 | 0.46 | 0.34 | 0.38 | 0.54 | 0.59 |
| Tykodi et al | 2019 | Renal cell carcinoma | anti-PD-1 therapy | PD-L1 IHC | 0.58 | 110 | 0.58 | 0.59 | 0.44 | 0.71 | 0.41 | 0.56 | 0.29 | 0.43 |
| Wang et al | 2018 | Urothelial cancer | anti-PD-1 therapy | Others | 0.67 | 214 | 0.67 | 0.55 | 0.31 | 0.85 | 0.45 | 0.69 | 0.15 | 0.33 |

| | | | | | | | | | | | | | | |
|---|---|---|---|---|---|---|---|---|---|---|---|---|---|---|
| Wang et al | 2018 | Urothelial cancer | anti-PD-1 therapy | GEP | 0.51 | 214 | 0.51 | 0.50 | 0.23 | 0.78 | 0.50 | 0.77 | 0.22 | 0.49 |
| Weber et al | 2013 | Melanoma | anti-PD-1 therapy | PD-L1 IHC | 0.64 | 44 | 0.64 | 0.53 | 0.39 | 0.76 | 0.47 | 0.61 | 0.24 | 0.36 |
| Weber et al | 2015 | Melanoma | anti-PD-1 therapy | PD-L1 IHC | 0.65 | 119 | 0.65 | 0.62 | 0.44 | 0.80 | 0.38 | 0.56 | 0.20 | 0.35 |
| Wolchok et al | 2013 | Melanoma | anti-PD-1 therapy+anti-CTLA4 | PD-L1 IHC | 0.40 | 35 | 0.40 | 0.65 | 0.46 | 0.59 | 0.35 | 0.54 | 0.41 | 0.60 |
| Wolchok et al | 2017 | Melanoma | anti-PD-1 therapy | PD-L1 IHC | 0.69 | 288 | 0.69 | 0.49 | 0.54 | 0.65 | 0.51 | 0.46 | 0.35 | 0.31 |
| Wolchok et al | 2017 | Melanoma | anti-PD-1 therapy+anti-CTLA4 | PD-L1 IHC | 0.60 | 278 | 0.60 | 0.51 | 0.65 | 0.46 | 0.49 | 0.35 | 0.54 | 0.40 |
| Younes et al | 2016 | Hodgkin's lymphoma | anti-PD-1 therapy | PD-L1 IHC | 0.78 | 40 | 0.78 | 0.67 | 0.97 | 0.20 | 0.33 | 0.03 | 0.80 | 0.22 |
| Zhu et al | 2018 | Hepatocellular Carcinoma | anti-PD-1 therapy | PD-L1 IHC | 0.72 | 103 | 0.72 | 0.54 | 0.25 | 0.90 | 0.46 | 0.75 | 0.10 | 0.28 |
| Zhu et al | 2018 | Hepatocellular Carcinoma | anti-PD-1 therapy | PD-L1 IHC | 0.72 | 104 | 0.72 | 0.55 | 0.25 | 0.90 | 0.45 | 0.75 | 0.10 | 0.28 |

*Study-level metrics are only shown for biomarker criteria with maximum balance accuracy

### Supplementary Figures

Supplementary Figure 1. Literary review description

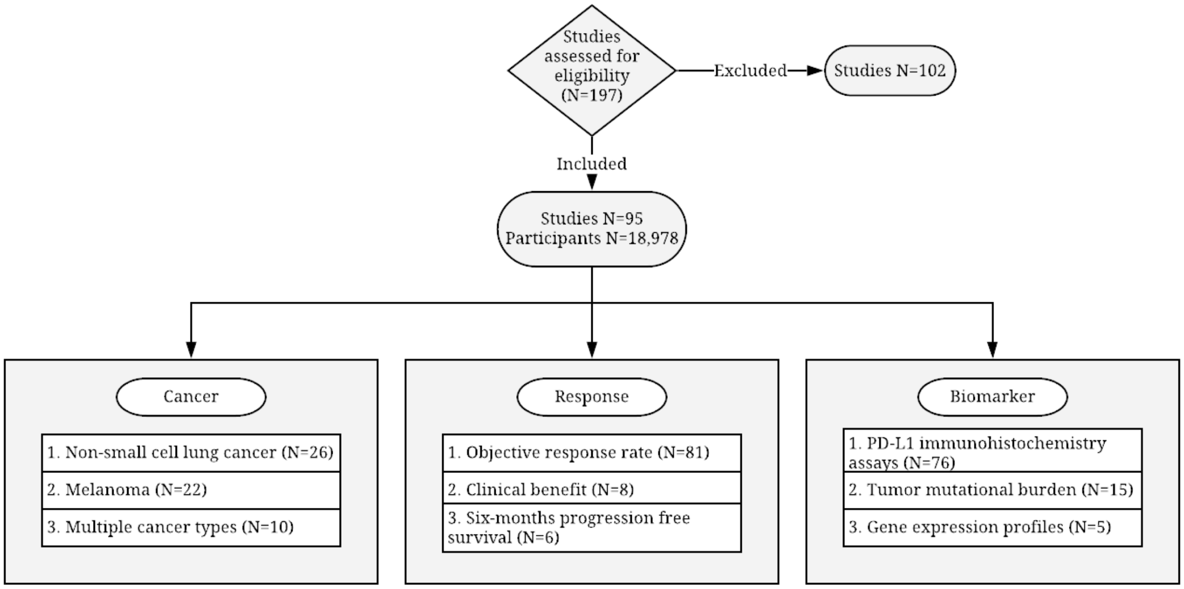

**Supplementary Figure 2. Results of bootstrap hypothesis tests within melanoma subset**

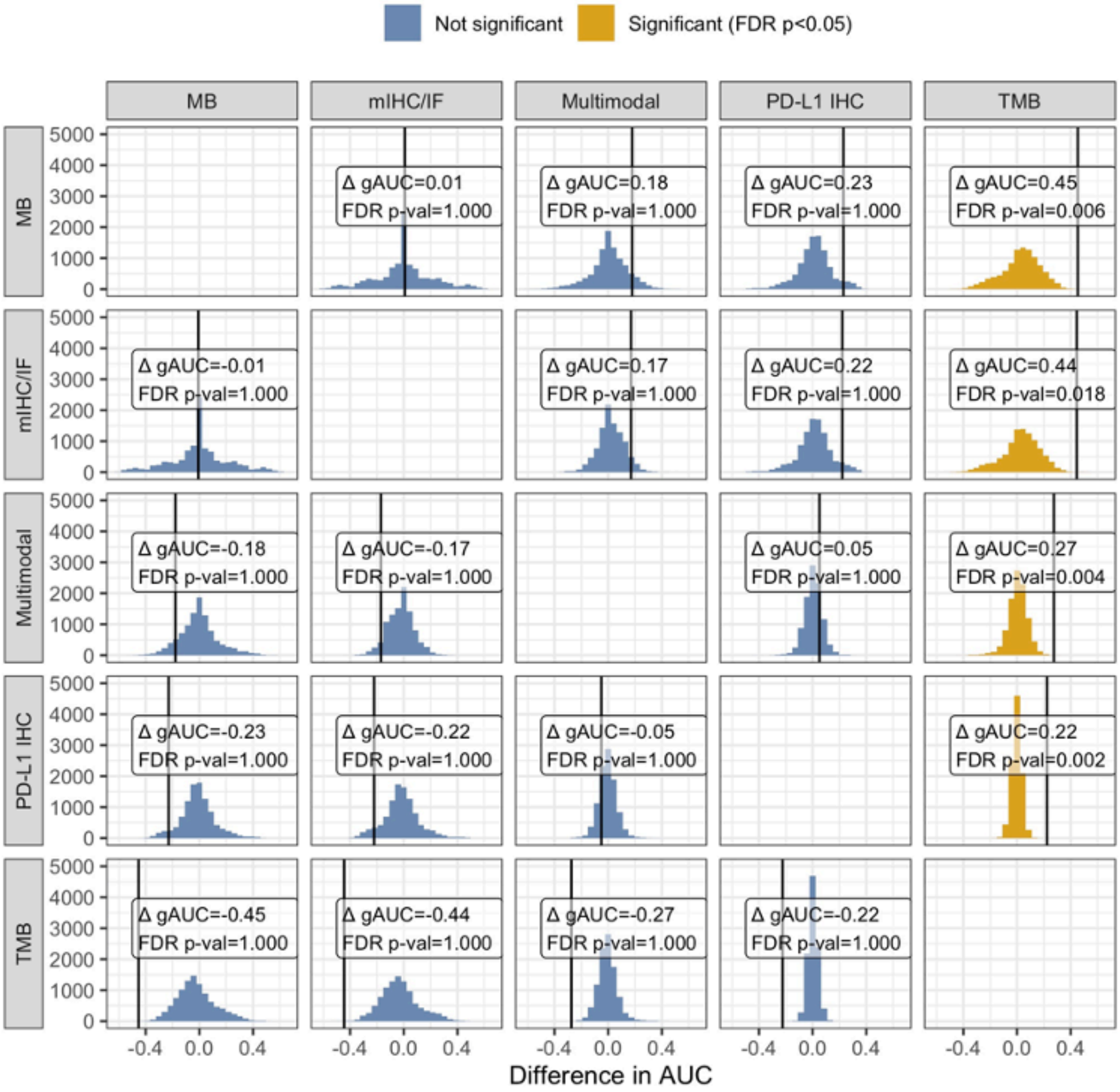

**Supplementary Figure 3. Results of bootstrap hypothesis tests within multiple cancer subset**

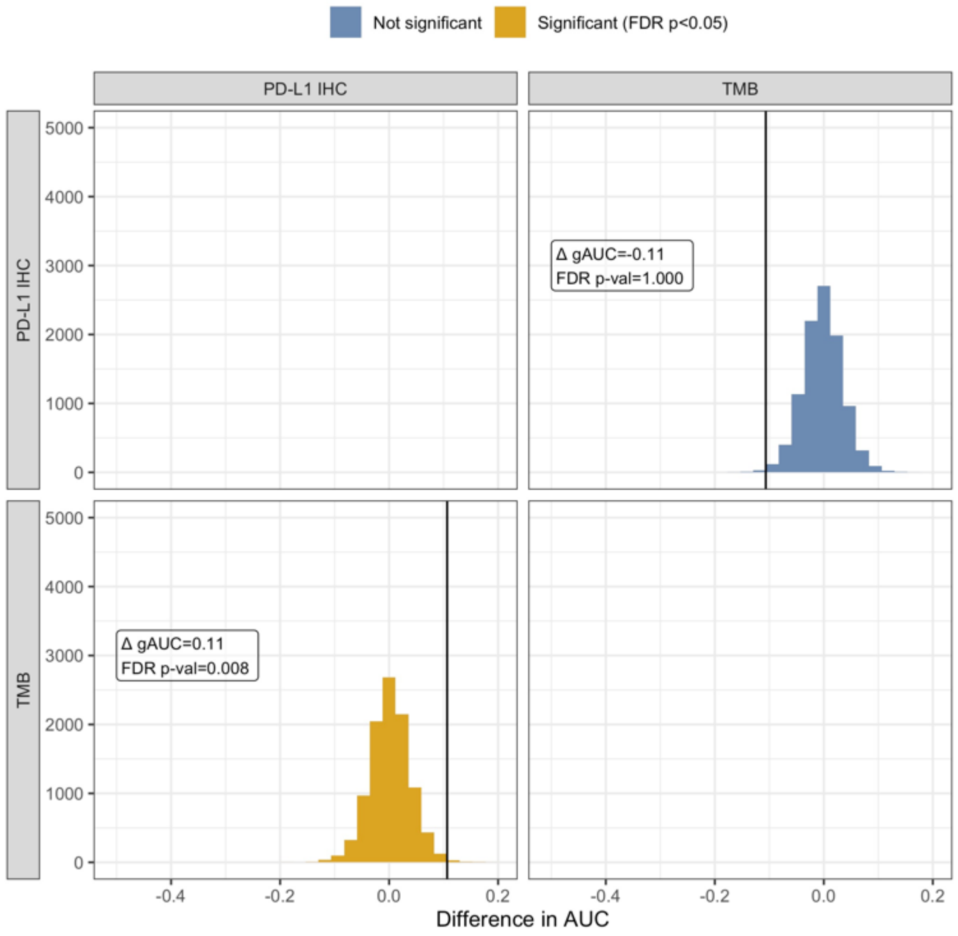

**Supplementary Figure 4. Results of bootstrap hypothesis tests within non-small-cell lung cancer subset**

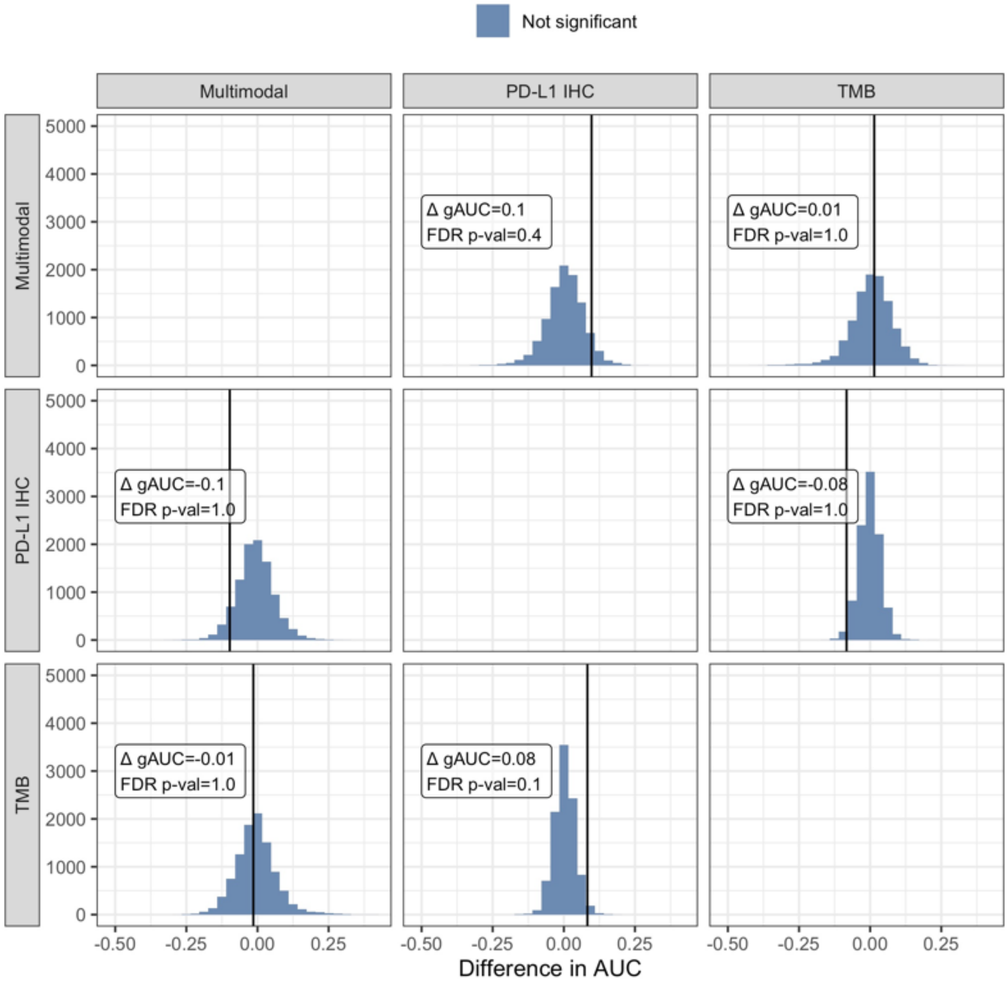

**Supplementary Figure 5. Results of bootstrap hypothesis tests across all cancer types**

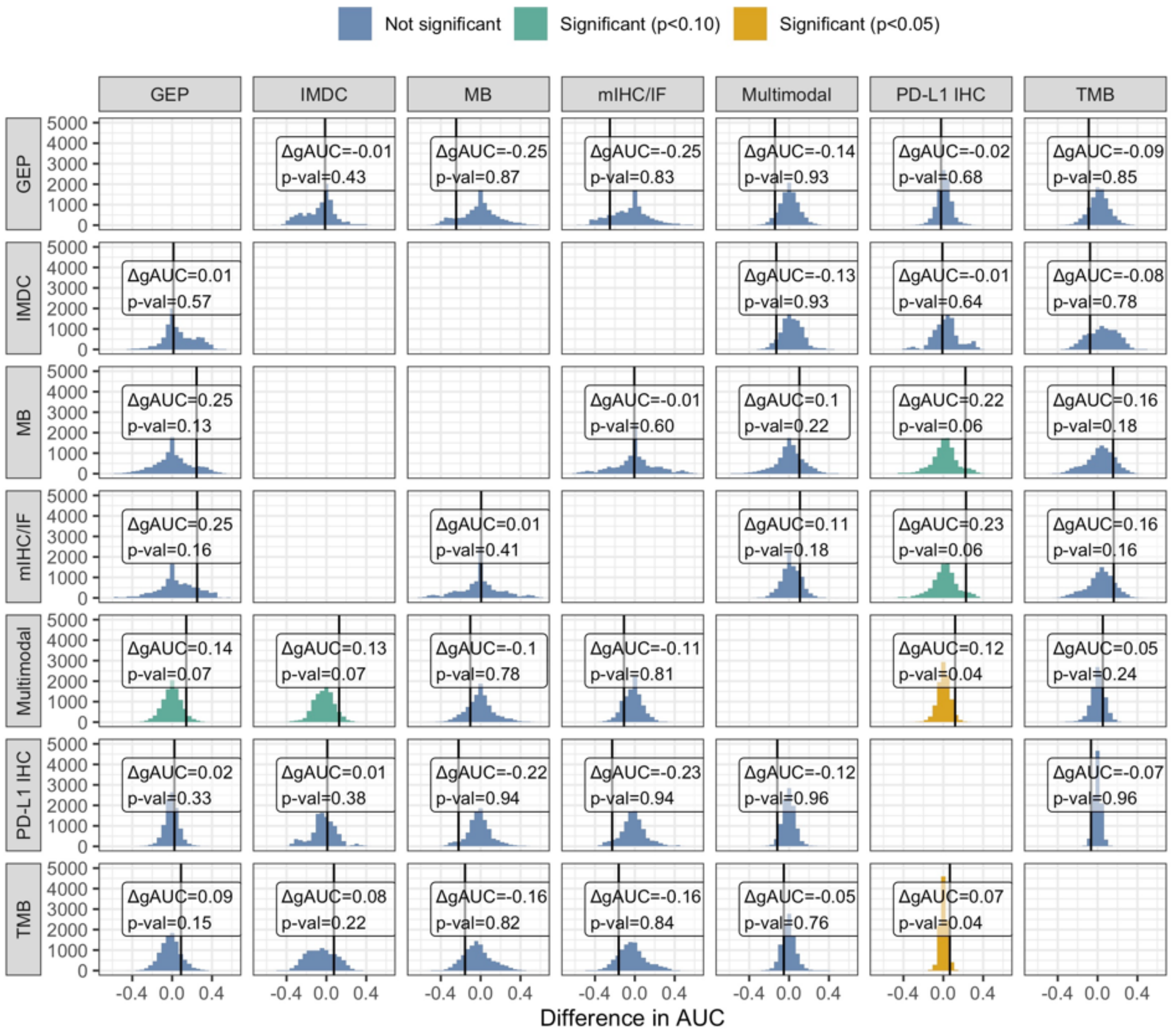

**Supplementary Figure 6. sROC curves of biomarkers for all cancer types**

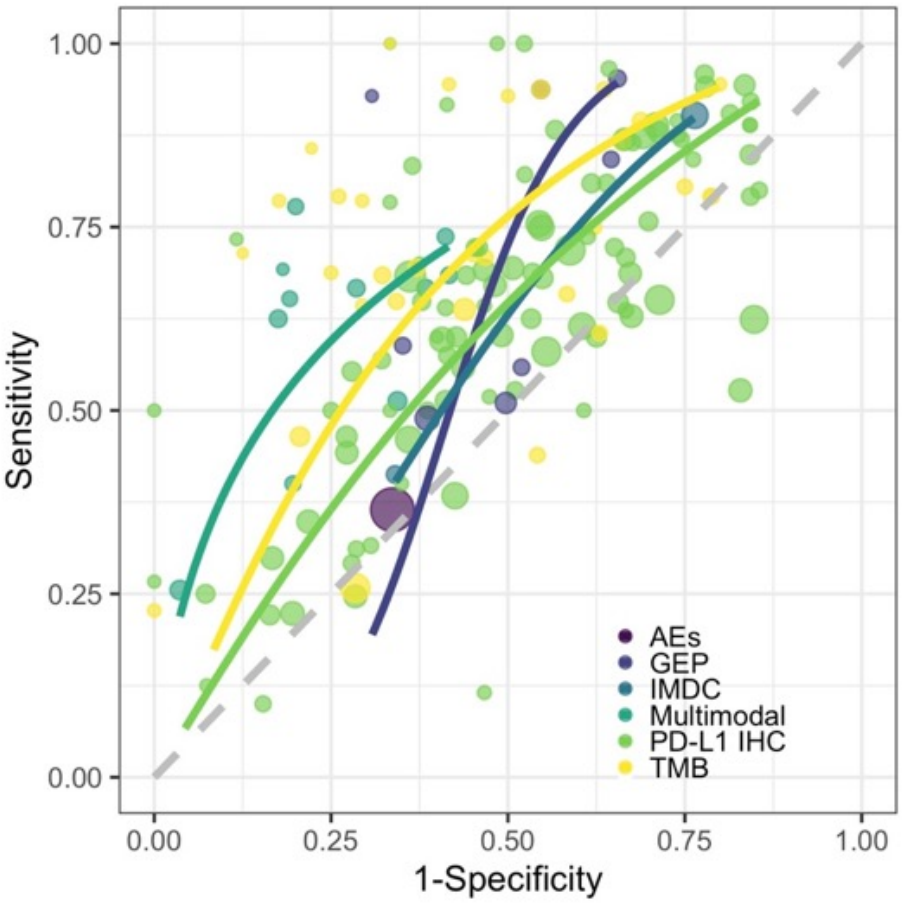

**Supplementary Figure 7. sROC curves of multimodal and multiplex immunohistochemistry assays (mIHC/IF) for all cancer types**

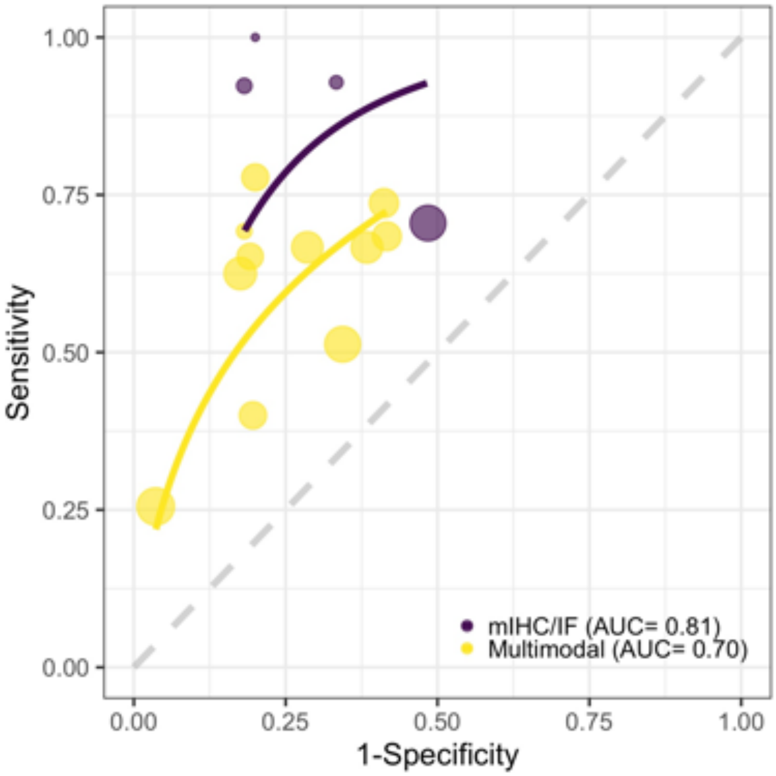

### Prisma Checklist

#### PRISMA 2009 Checklist

| Section/topic | # | Checklist item | Reported on page # |
|---|---|---|---|
| **TITLE** | | | |
| Title | 1 | Identify the report as a systematic review, meta-analysis, or both. | 1 |
| **ABSTRACT** | | | |
| Structured summary | 2 | Provide a structured summary including, as applicable: background; objectives; data sources; study eligibility criteria, participants, and interventions; study appraisal and synthesis methods; results; limitations; conclusions and implications of key findings; systematic review registration number. | 2 |
| **INTRODUCTION** | | | |
| Rationale | 3 | Describe the rationale for the review in the context of what is already known. | 3 |
| Objectives | 4 | Provide an explicit statement of questions being addressed with reference to participants, interventions, comparisons, outcomes, and study design (PICOS). | 4 |
| **METHODS** | | | |
| Protocol and registration | 5 | Indicate if a review protocol exists, if and where it can be accessed (e.g., Web address), and, if available, provide registration information including registration number. | |
| Eligibility criteria | 6 | Specify study characteristics (e.g., PICOS, length of follow-up) and report characteristics (e.g., years considered, language, publication status) used as criteria for eligibility, giving rationale. | 4 |
| Information sources | 7 | Describe all information sources (e.g., databases with dates of coverage, contact with study authors to identify additional studies) in the search and date last searched. | 4 |
| Search | 8 | Present full electronic search strategy for at least one database, including any limits used, such that it could be repeated. | 4 |
| Study selection | 9 | State the process for selecting studies (i.e., screening, eligibility, included in systematic review, and, if applicable, included in the meta-analysis). | 4 |
| Data collection process | 10 | Describe method of data extraction from reports (e.g., piloted forms, independently, in duplicate) and any processes for obtaining and confirming data from investigators. | 5 |
| Data items | 11 | List and define all variables for which data were sought (e.g., PICOS, funding sources) and any assumptions and simplifications made. | 5 |
| Risk of bias in individual studies | 12 | Describe methods used for assessing risk of bias of individual studies (including specification of whether this was done at the study or outcome level), and how this information is to be used in any data synthesis. | 8 |

| | | | |
|---|---|---|---|
| Summary measures | 13 | State the principal summary measures (e.g., risk ratio, difference in means). | 5-8 |
| Synthesis of results | 14 | Describe the methods of handling data and combining results of studies, if done, including measures of consistency (e.g., $I^2$) for each meta-analysis. | 8 |

| Section/topic | # | Checklist item | Reported on page # |
|---|---|---|---|
| Risk of bias across studies | 15 | Specify any assessment of risk of bias that may affect the cumulative evidence (e.g., publication bias, selective reporting within studies). | |
| Additional analyses | 16 | Describe methods of additional analyses (e.g., sensitivity or subgroup analyses, meta-regression), if done, indicating which were pre-specified. | |
| **RESULTS** | | | |
| Study selection | 17 | Give numbers of studies screened, assessed for eligibility, and included in the review, with reasons for exclusions at each stage, ideally with a flow diagram. | 9 |
| Study characteristics | 18 | For each study, present characteristics for which data were extracted (e.g., study size, PICOS, follow-up period) and provide the citations. | 5 and SI |
| Risk of bias within studies | 19 | Present data on risk of bias of each study and, if available, any outcome level assessment (see item 12). | |
| Results of individual studies | 20 | For all outcomes considered (benefits or harms), present, for each study: (a) simple summary data for each intervention group (b) effect estimates and confidence intervals, ideally with a forest plot. | SI |
| Synthesis of results | 21 | Present results of each meta-analysis done, including confidence intervals and measures of consistency. | 9 |
| Risk of bias across studies | 22 | Present results of any assessment of risk of bias across studies (see Item 15). | |
| Additional analysis | 23 | Give results of additional analyses, if done (e.g., sensitivity or subgroup analyses, meta-regression [see Item 16]). | |
| **DISCUSSION** | | | |
| Summary of evidence | 24 | Summarize the main findings including the strength of evidence for each main outcome; consider their relevance to key groups (e.g., healthcare providers, users, and policy makers). | 12 |
| Limitations | 25 | Discuss limitations at study and outcome level (e.g., risk of bias), and at review-level (e.g., incomplete retrieval of identified research, reporting bias). | 15 |
| Conclusions | 26 | Provide a general interpretation of the results in the context of other evidence, and implications for future research. | 16 |
| **FUNDING** | | | |
| Funding | 27 | Describe sources of funding for the systematic review and other support (e.g., supply of data); role of funders for the systematic review. | 17 |

*From:* Moher D, Liberati A, Tetzlaff J, Altman DG, The PRISMA Group (2009). Preferred Reporting Items for Systematic Reviews and Meta-Analyses: The PRISMA Statement. PLoS Med 6(7): e1000097. doi:10.1371/journal.pmed1000097

For more information, visit: **www.prisma-statement.org**.
